# Studying the course of Covid-19 by a recursive delay approach

**DOI:** 10.1101/2021.01.18.21250012

**Authors:** Matthias Kreck, Erhard Scholz

## Abstract

In an earlier paper we proposed a recursive model for epidemics; in the present paper we generalize this model to include the asymptomatic or unrecorded symptomatic people, which we call *dark people* (dark sector). We call this the SEPAR_*d*_-model. A delay differential equation version of the model is added; it allows a better comparison to other models. We carry this out by a comparison with the classical SIR model and indicate why we believe that the SEPAR_*d*_ model may work better for Covid-19 than other approaches.

In the second part of the paper we explain how to deal with the data provided by the JHU, in particular we explain how to derive central model parameters from the data. Other parameters, like the size of the dark sector, are less accessible and have to be estimated more roughly, at best by results of representative serological studies which are accessible, however, only for a few countries. We start our country studies with Switzerland where such data are available. Then we apply the model to a collection of other countries, three European ones (Germany, France, Sweden), the three most stricken countries from three other continents (USA, Brazil, India). Finally we show that even the aggregated world data can be well represented by our approach.

At the end of the paper we discuss the use of the model. Perhaps the most striking application is that it allows a quantitative analysis of the influence of the time until people are sent to quarantine or hospital. This suggests that imposing means to shorten this time is a powerful tool to flatten the curves.

## Introduction

There is a flood of papers using the standard S(E)IR models for describing the outspread of Covid-19 and for forecasts. Part of them is discussed in [19]. We propose alternative delay models and explain the differences.

In [10] we have proposed a discrete delay model for an epidemic which we call SEPAR-model (in our paper we called it *SEPIR* model). In this paper we explained why and under which conditions the model is adequate for an epidemic. In the present note we add two new compartments reflecting asymptomatic or symptomatic, but not counted, infected which we call the *dark sector*. We call this model the *generalized SEPAR-model*, abbreviated *SEPAR*_*d*_, where *d* stands for dark. This is our main new contribution. We will discuss the role of the dark sector in a theoretical comparison of the SEPAR_*d*_-model with the SEPAR model. We will see, that – as expected – as long as the number of susceptibles is nearly constant, the difference of the two models is small, but in the long run it matters.

A second topic in this paper is a comparison with the standard SIR model. This comparison has two aspects, a purely theoretical one by comparing the different fundaments on which the models are based, and a numerical one. For comparing two models it is helpful to derive them from similar inputs. For this we pass from the discrete model leading to difference equations to a continuous model, replacing difference equations by differential equations. These differential equations fit into the general approach developed by Kermack/McKendrick in [9], as we learnt from O. Diekmann. The analytic model resulting from our discrete model has been introduced independently by J. Mohring and coauthors [12] and, more recently, by B. Shayit and M. Sharma [20]. Also F. Balabdaoui and D. Mohr work with a discrete delay approach with additional compartments and a stratification into different age layers adapted to the Swiss context [3]. Recently y R. Feßler has written a paper [7] in which different differential equation models are discussed and compared, including the classical S(E)IR-model and the analytic version of our model. Some hints to earlier papers on the analytic delay approach can be found there.

In the second part of the present paper we apply the SEPAR model to selected countries and to the aggregated data of the world. To do so we first lay open how to pass from the data provided by the *Humdata* project of the JHU to the model parameters. The data themselves are obviously not reflecting the actual outspread correctly, which is most visible by the lower numbers of reported cases during weekends. But in addition there are aspects of the data which need to be corrected like for example a delay of reporting of recovered cases. All this is discussed carefully. Reliable data about the size of the dark sector are only available in certain countries where such studies were carried out. We found such studies for Germany and Switzerland, for other countries we estimate these numbers as good as we can. The case of Switzerland is particular interesting since the effect of the dark sector which started to play a non-negligible role for the overall dynamics of the epidemic in the later part of 2020 for the majority of the countries discussed here (India, USA, Brazil, France) can be studied there particularly well. For that reason we begin with this country and discuss the role of the dark sector in detail.

The paper closes with a discussion about what one can learn from the applications of the SEPAR model. We address three topics: The role of the constancy intervals, the role of the dark sector and the the influence of the time between infection and quarantine. The latter is perhaps the most striking application of our model offering a door for flattening the curves by sending people faster into quarantine, a restriction which imposes much less harm to the society than other means.

## PART I: Theoretical framework

### 1. The SEPAR model and its comparison with other models

#### 1.1. The SEPAR_*d*_ model

We begin by pointing out that we have changed our notation from [10]. The compartment consisting of those who are isolated after sent to quarantine or hospital, which there was called *I*, is now being denoted by *A* like *actually* infected, in some places also described – although a bit misleading – as “active” cases (e.g. in Worldometer). This is why we speak now of the SEPAR model rather than of SEPIR.

Let us first recall the compartments introduced in [10]. We observe 5 compartments which we call *S, E, P*, *A, R*, which people pass through in this order: *Susceptibles* in compartment *S* moving after infection to compartment *E*, where they are *exposed* but not infectious, after they are infected by people from compartment *P* which comprises the actively infectious people, those which *propagate* the virus. After *e* days they move from *E* to the compartment *P*, where they stay for *p* days. After diagnosis they are sent into quarantine or hospital and become members of the compartment *A*, where they no longer contribute to the spread of the virus although they are then often counted as the actual cases of the statistics. In order not to overload the model with too many details, we pass over the recording delay between diagnosis and the day of being recorded in the statistics. After another *q* days the recorded infected move from compartment *A* to the compartment *R* of *removed* (recovered or dead).

We add two more compartments reflecting the role of the dark sector. There are two types of infected people, those who will at some moment be tested and counted, and those who are never tested, which we call people in the dark. This suggests to decompose compartment *P* into two disjoint sub-compartments: *P*_*c*_ of people who after *p*_*c*_ days will be tested and counted and move to compartment *A*, and the collection *P*_*d*_ of people who after a longer period of *p*_*d*_ days get immune and so move into a new compartment *R*_*d*_ of removed people in the dark. To distinguish these removed people in the dark from those who come from compartment *P* after recovery or death we introduce another new compartment *R*_*c*_ of those removed people who occur in the statistics. Of course *R* = *R*_*c*_∪*R*_*d*_.

The introduction of the dark sector in addition to the sector of counted people leads to the picture that for the infected persons leaving compartment *E* there is a branching process: a certain fraction *α*(*k*) of people from *E* moves to compartment *P*_*c*_ at day *k*, whereas the fraction 1−*α*(*k*) of people moves to compartment *P*_*d*_.

The existence of these compartments is a fundamental assumption which distinguishes the SEPAR_*d*_ model from many other models including standard SIR. The existence of these compartments is closely related to our picture of an epidemic like Covid-19. Of course this is a simplification. If one assumes that the passage from compartment *S* to compartment *E* takes place at a certain moment, the duration of the stay in the next compartments varies from case to case. But it looks natural to take the average of these durations leading for the different lengths *e, p*_*c*_, *p*_*d*_, and *q*. All these have to be estimated from available information.

Once one has agreed to this there is another fundamental assumption. This concerns the dynamics of the epidemic. Each person in compartment *P* has a certain average number *κ*(*k*) of contacts at day *k*. Depending on the strength of the infectious power of an individual the contacts will lead to newly exposed people. It is natural to model this development of the strength of infectiousness by a function *A*(*τ*), which measures the strength *τ* days after entry into compartment *P*. Again we simplify this very much, by replacing *A*(*τ*) by a constant *γ*, the average value of this assumed function. We will discuss this assumption later on in the light of information available for Covid-19. Given the parameters *κ*(*t*) and *γ* our next assumption is that, if we ignore the dark sector and set *η*(*k*) := *γκ*(*k*), the dynamics of the infection can be described by the following formula:

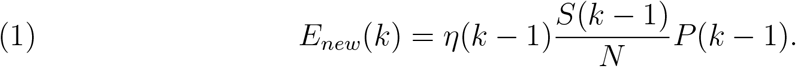

Here *E*_*new*_(*k*) is the number of additional members of compartment *E* at day *k* infected at day (*k*−1) by people from compartment *P* and *N* the total number of the population. This is a very plausible formula. We call *η*(*k*) the *daily strength of infection*. It is an integrated expression for the averaged contact behaviour of the population and the aggressiveness of the virus.

This is the dynamics if we ignore the dark sector. But members of compartment *P*_*d*_ also infect. We assume that the contacts are equal to those in compartment *P*_*c*_. But the average of the strength of infection of people from *P*_*d*_ may be smaller than for those in compartment *P*_*c*_, since in general they can be expected to stay longer in their compartment until they are immune and the strength of infection goes further down. Thus we introduce a separate measure *γ*_*c*_ for those in compartment *P*_*c*_ and *γ*_*d*_ = *ξγ*_*c*_, with 0≤*ξ*≤1, for those in compartment *P*_*d*_.

Using this the equation (1) has to be replaced by:

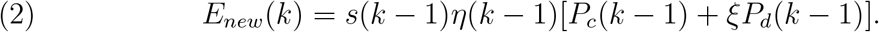

Given this infection equation the rest of the model just describes the time shifting passage of infected from one compartment into the next and counts their cardinality at day *k*. As usual we denote the latter by *S*(*k*), *E*(*k*), *P*_*c*_(*k*), *P*_*d*_(*k*), *A*(*k*), *R*_*c*_(*k*) and *R*_*d*_(*k*). How such a translation is justified is explained in [10]. So we can just write down the self explaining formulas here:

##### Introduction (definition) of the discrete SEPAR_*d*_ model

*Let e, p*_*c*_, *p*_*d*_, *q be integers standing for the duration of staying in the corresponding compartments*, 0≤*α*(*k*) ≤ 1 *be branching ratios at day k between later registered infected and those which are never counted, η*(*k*) *be positive real numbers describing the daily strength of infection for k* ≥ 0, *while η*(*k*) = 0 *for k <* 0, *and ξ* ≤ 1 *a non-negative real number. Using* 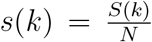 *the quantities S*(*k*), *E*(*k*), *E*_*new*_(*k*), *P*_*c*_(*k*), *P*_*d*_(*k*), *A*(*k*), *R*_*c*_(*k*), *R*_*d*_(*k*) *of the SEPAR*_*d*_ *model are given by*

a. *the start condition:* *since the model is recursive we need an input for the first e* + *p*_*d*_ *days (which we shift to negative values of k), i*.*e. start data E*_*start*_(*k*) *for* 1— (*e* + *p*_*d*_) ≤ *k* ≤0, *while E*_*start*_(*k*) = 0 *for all k >* 0, *P*_*c*_(*k*) = *P*_*d*_(*k*) = *A*(*k*) = *R*_*c*_(*k*) = *R*_*d*_(*k*) = 0 *(or some other well defined start values, cf. sec. 2*.*3) for k <* 1 − (*e* + *p*_*d*_);
b. *the recursion scheme for k* ≥ 1 − (*e* + *p*_*d*_):

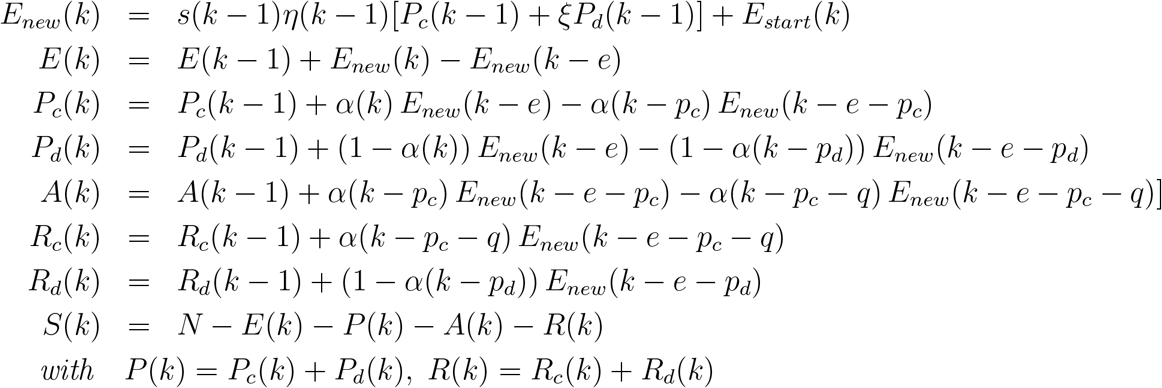

The reason for this definition is easy to see. Additional people in, for example, compartment *P*_*c*_ at the day *k* are (*P*_*c*_)_*new*_(*k*) = *α*(*k*)*E*_*new*_(*k*— *e*), while *α*(*k*)*E*_*new*_(*k*− (*e* + *p*)) move to the next compartment. Similar formulas hold for the compartments *P*_*d*_, *A, R*_*c*_ and *R*_*d*_.

An important parameter in an epidemic is the *reproduction number ρ*, the number of people infected by a single infectious person during its life time. If we assume that *κ*(*k*) and *s*(*k*) may be considered as constant during *p*_*d*_ days about *k*, we can derive this number from the equations. It is

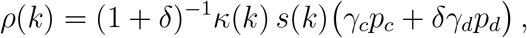

Where 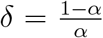. For *s*(*k*) = 1 it is usually called the *basic* reproduction number, in order to distinguish it from the *effective* reproduction number with *s*(*k*) *<* 1. In part I of this paper we usually mean the basic reproduction number when we speak of reproduction number, while in the part II the decreasing *s*(*k*) hast to be taken into account and we usually speak of the effective reproduction number, also without use of the attribute “effective”.

If we set *α*(*k*) = 1, *P*_*d*_ = 0 and *R*_*d*_ = 0, we obtain the SEPAR model without dark sector as a special case of the SEPAR_*d*_ model. Effects of vaccination can easily be implemented by sending the according number of persons directly from *S* to *R*.

For later use the following observation is useful. The number of people in a given compartment at day *k* is the sum of additional entries at previous days, for example

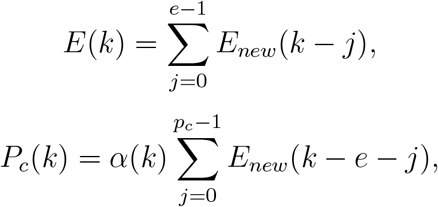

and so on for *P*_*d*_(*k*) and *A*(*k*).

We abbreviate

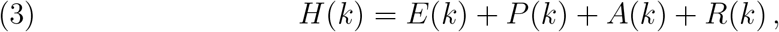

the number of *herd immunized* (without vaccination). Then the recursion scheme implies:

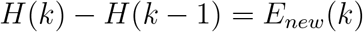

Putting this into the formula above: 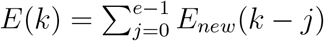, we obtain:

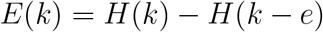

and similarly

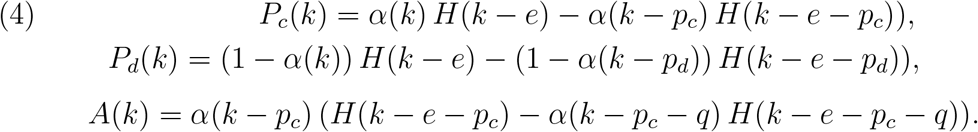

Using *R*_*c*_(*k*) − *R*_*c*_(*k* - 1) = *α* (*k* − *p*_*c*_ − *q*)*E*_*new*_(*k* − *e* − *p*_*c*_ − *q*) = *α* (*k* − *p*_*c*_ − *q*)*H*(*k* −*e* − *p*_*c*_ − *q*) − *α* (*k* − *p*_*c*_ − *q* − 1) *H*(*k* − *e* − *p*_*c*_ − *q* − 1) we conclude:

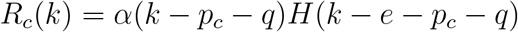

and similarly

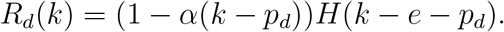

This gives a very simple structure of the model in terms of a single recursion equation.

##### SEPAR_*d*_-model

*The recursion scheme of the SEPAR*_*d*_ *model is given by a single recursion equation:*

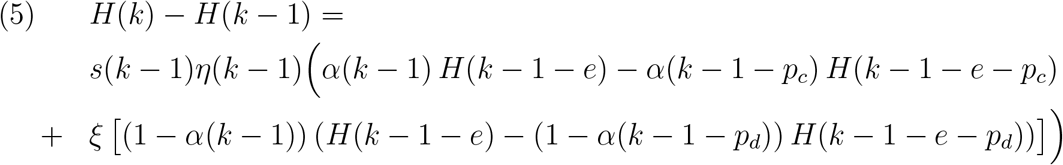

*and the functions S*(*k*), *E*(*k*), *P*_*c*_(*k*), *P*_*d*_(*k*), *A*(*k*), *R*_*c*_(*k*) *and R*_*d*_(*k*) *are given in terms of H*(*k*) *by the equations above*.

*If we pass from a daily recursion to a infinitesimal recursion, replacing the difference equation by a differential equation, we obtain the continuous recursion scheme, where now all functions are differentiable functions of the time t:*

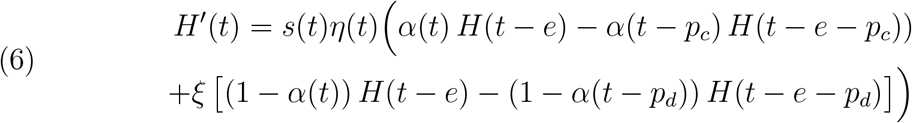

In both cases, discrete and continuous, consistent start conditions in an interval of length *e* + *p* have to be added. For the discrete case see sec. 2.3. If we remove the dark sector, the continuous model was independently obtained in [12].

The branching ration *α* and with it the number 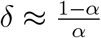 of unrecorded infected for each newly recorded one varies drastically in space and time, roughly in the range 1 ≤ *δ* ≤ 50. For Switzerland and Germany serological studies in late 2020 conclude *δ* ≈ 2, for the USA a recent study finds *δ* ≈ 8 and in part of India (Punjab) a serological study found values indicating *δ*≈ 50.^1^ For our choice of the model parameter see below, section 3.

Besides the determination of *α* one needs to know the difference between *p*_*c*_ and *p*_*d*_ and between *η*_*c*_(*k*) and *η*_*d*_(*k*), if one wants to apply the *SEPAR*_*d*_-model. As explained above we estimate *p*_*c*_ = 7. The mean time of active infectivity of people who are not quarantined seems to be not much longer, although in some cases it is. According to the study [21, p. 466] “no isolates were obtained from samples taken after day 8 (after occurrence of symptoms) in spite of ongoing high viral loads”. This allows to work with an estimate *p*_*d*_ = 10, and so it is not much larger than *p*_*c*_. A comparison of the *SEPAR*_*d*_ model with a simplified version, where we assume *p*_*c*_ = *p*_*d*_ =: *p* and *η*_*c*_(*k*) = *η*_*d*_(*k*) =: *η*(*k*) shows that with these values the difference is very small (see figure 1). In the following we therefore work with the simplified *SEPAR*:*d* model setting *p*_*c*_ = *p*_*d*_ =: *p*.

**Figure 1.**
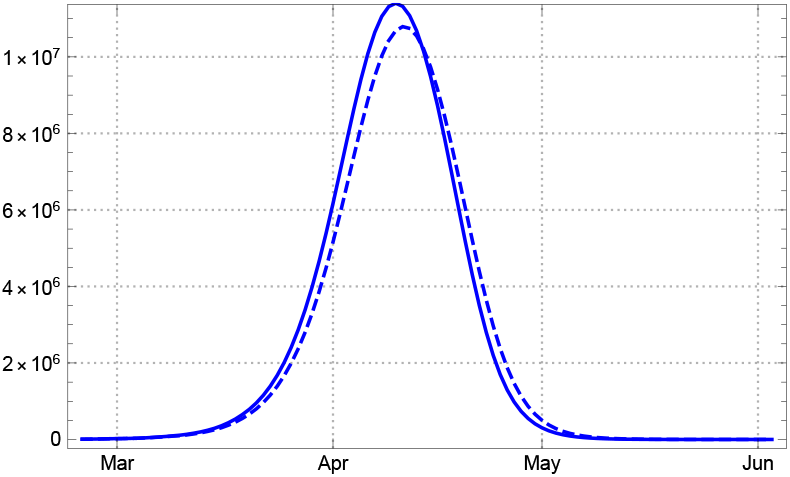
Comparison of SEPAR_*d*_ model for *A*(*k*) between dark sector with *p*_*c*_ = 7, *p*_*d*_ = 10, *ξ* = 0.9 (solid blue) and simplified dark sector *p*_*c*_ = *p*_*d*_ = *p* = 7, *ξ* = 1 (dashed blue), assuming constant *η*.

Recent studies indicate that the number of asymptomatic infected is often as low as about 1 in 5 symptomatic unrecorded and is thus much smaller than originally expected [13]. Although asymptomatic infected are there reported to be considerably less infective than the symptomatic ones, their relatively small number among all unreported cases justifies to work in the simplified dark model with the assumption *η*_*d*_(*k*) ≈ *η*_*c*_(*k*) =: *η*(*k*). If we set *P* (*k*) = *P*_*c*_(*k*) + *P*_*d*_(*k*) as above, we see that *P*_*c*_(*k*) = *α*(*k*)*P* (*k*) and *P*_*d*_(*k*) = (1 − *α*(*k*))*P* (*k*).

In part II we discuss how the time dependent parameter *η*(*k*) can be derived from the data and a rough estimate of the dark factor *δ* can be arrived at, although it lies in the nature of the dark sector that information is difficult to obtain.

A comparison of the SEPAR model (*δ* = 0) with the simplified SEPAR_*d*_ model is given in fig. 2 for a constant parameter *α* = 0.2, respectively dark factor *δ* = 4 and constant reproduction coefficient *ρ* = 3. This illustrates the influence of the dark factor from the theoretical viewpoint.

**Figure 2.**
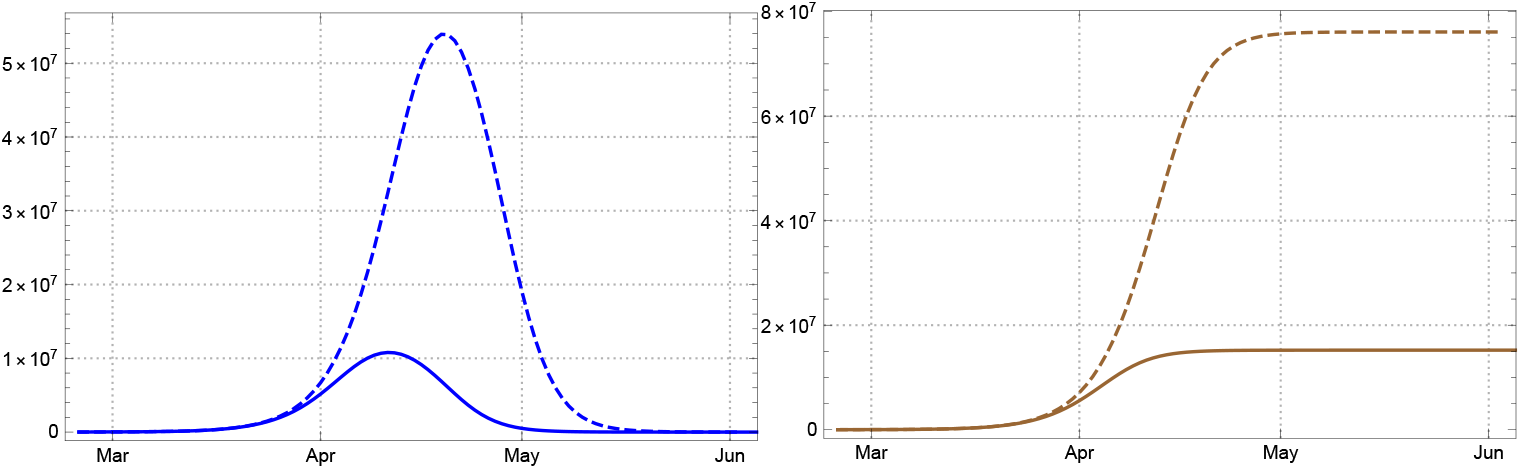
Comparison of the course of an epidemic with constant reproduction number *ρ* = 3 without dark sector (dashed), and with dark sector *δ* = 4 (solid lines): Left counted number of actual infected *A*_*c*_(*k*) (blue). Right: total number of confirmed infected *A*_*tot c*_(*k*) = *A*_*c*_(*k*) + *R*_*c*_(*k*) (brown).

#### 1.2. The S(E)IR models and their assumptions

Whereas the derivation of the SEPAR_*d*_-model is based on the idea of disjoint compartments, infected people pass through in time, there is a different approach with goes back to the seminal paper [9]. A special case is the standard SIR-model or SEIR model. It seems that most people use this as a black box without observing the assumptions on which it is built. One should keep these assumptions in mind whenever one applies a model. There is a modern and easy to understand paper by Breda, Diekmann, and de Graaf with the title: *On the formulation of epidemic models (an appraisal of Kermack and McKendrick)* [4], which explains the general derivation. In the introduction the authors state that the Kermack/McKendrick paper was cited innumerable times and continue: “But how often is it actually read? Judging from an incessant misconception of its content one is inclined to conclude: hardly ever! If one observes the principles from which the S(E)IR models are derived one should be hesitant to apply it to Covid-19.

Following [4] we shortly repeat the assumptions on which the general Kermack/McKendrick approach is based. The general model considers a function

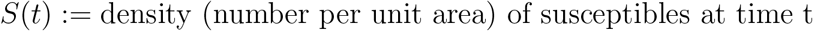

and related to this a function

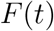

called the force of infection at time t. By definition, the force of infection is the probability per unit of time that a susceptible becomes infected. So, if numbers are large enough to warrant a deterministic description, we have

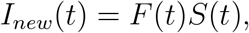

where *I*_*new*_(*t*) is defined as the number of new cases per unit of time and area. The functions *S* and *I* are related by the equation:

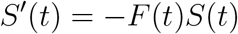

Then the central modelling ingredient is introduced:

*A*(*τ*) := expected contribution to the force of infection *τ* units of time ago.

Alone from this ingredient an integral differential equation is derived, which gives the model equations. For more details we also refer to a recent paper by Robert Feßler who derived the integral equation independently [7].

Already here we see a different view of an epidemic. No compartments and their cardinality are mentioned; in their place the authors mention only certain functions. If the function *A* is assumed to decay exponentially,

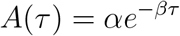

with constants *α, β*. The model derived from this input is called the *standard SIR-model*. It leads to two ordinary differential equations in the variable *t*:

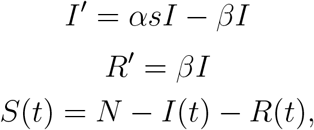

where *N* as before is the number of the population and 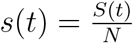.

For the *standard SEIR-model* there is an additional function *E*(*t*) measuring the. *exposed* and the input function is now

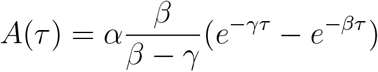

This leads to 3 ordinary differential equations in *t*

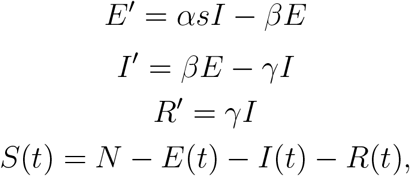

where *N* as before is the number of the population. The infection function *A*(*τ*) considered here determines the convolution part of an integral kernel in Feßler’s approach mentioned above.

If Breda et al. are right, readers should be critical to papers applying the S(E)IR models without explaining why the models, given their fundaments, are applicable. As far as we can see, the assumption of an exponential decay *A*(*t*) is often not mentioned by authors applying it in situations where it would be necessary to discuss whether this assumption can be reasonably made. In a situation like Covid-19 where infectious people are isolated as soon as possible, it seems questionable whether this assumption holds. We are surprised that in most of the papers we have seen, which apply the S(E)IR model to an analysis of Covid 19, this problem is not even mentioned. This includes the papers of the group around Viola Priesemann which play an important part in the discussion about how to deal with Covid 19 in Germany [6],[5].

#### 1.3. Comparing SIR with SEPAR

When we want to compare the SIR models with the delay SEPAR model we have to lower, in a first step, the number of compartments by removing *E, P* and *A* and to replace them by a single compartment, called *I*, of infected people which are at the same time infectious. For this model we assume that infected susceptibles move right away to compartment *I*, where they stay for *p* days. In contrast to the SEPAR model it is assumed that these people are counted as actual infected people at the moment they are infected. After *p* days they are counted as recovered or dead. So it is a strong simplification of the SEPAR model, but it follows the same pattern as the SEPAR model since it is a delay model. We call it *d-SIR-model* (“d-”for delay) to distinguish it from the standard SIR-model. The equations for this model are based on the same principles as the SEPAR model:

##### The continuous delay d-SIR model

*Let p be a positive real number standing for the duration of staying in the compartment I of infected and infectious people. Let η*(*t*) *be a differentiable function measuring the strength of infection (including the effects of social constraints). The quantities S*(*t*), *I*(*t*), *R*(*t*) *of the delay SIR model are given by*

a. *the start condition:* *A differentiable function I*(*t*) *for* 0≤ *t < p*,
b. *and the delay differential equations:*

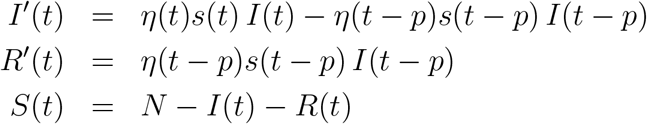

To compare this model with the standard SIR-model above we note that also the d-SIR model (like the continuous SEPAR_*d*_ model) can be derived from the principles of Kermack/McKendrick, as explained in [7]. One only has to take the product of the characteristic function of the interval [0, *p*] with *η* = *γκ* as the function *A*(*τ*).

To compare the two models one has to relate the input parameters. In the case of the d-SIR model they are *η* (for the comparison we assume that the contact rate is constant) and *p*, whereas for the SIR-model they are *α* and *β*. The role of *η* is that of *α* in the SIR-model, so we set *α* = *η*. There are several ways to relate the paramter *β* of the SIR model with *p* occurring in the d-SIR model. One is to assume that the total force of infection has to be the same if they describe the same developments, i.e. with the function *A*(*τ*) which is the product of the characteristic function of the interval [0, *p*] with *η* one has the condition:

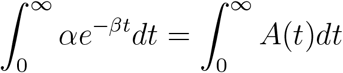

Then the second relation: 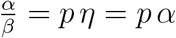 and thus

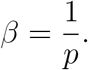

In both cases the reproduction number is 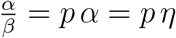.

If one applies this then there is a problem to find parameters so that at least at the beginning the two models are approximatively equal. Thus one can relate the tow models in a second way by choosing the parameters so that this is the case. For this we fix values for *α* and *β* and chose the start conditions of the d-SIR model so that they agree with the SIR-model during the first days. By construction of the SIR-model the function *I* is nearly an exponential function as long as the function *S* is nearly constant. Thus we chose the same exponential function as start values for the d-SIR model.

Then the question is whether there are differences of the model curves in the long run and how large the differences are. One should expect that the assumptions of an exponential decay regulating the strength of infection of an infectious person in the case of the SIR-model versus a period of *p* days, where the strength of infection is constant and after that goes immediately down to 0 in the case of the delay d-SIR, should result in higher values for the functions *I*(*t*) and *I*_*tot*_(*t*) = *I*(*t*) + *R*(*t*), the total number of infected until time *t* of the SIR model. The following graphics in which we assume a constant reproduction rate slightly above 1 show, in fact, a dramatic difference supporting the expectation. A similar observation can be found in [7, fig. 5, 6]

**Figure 3.**
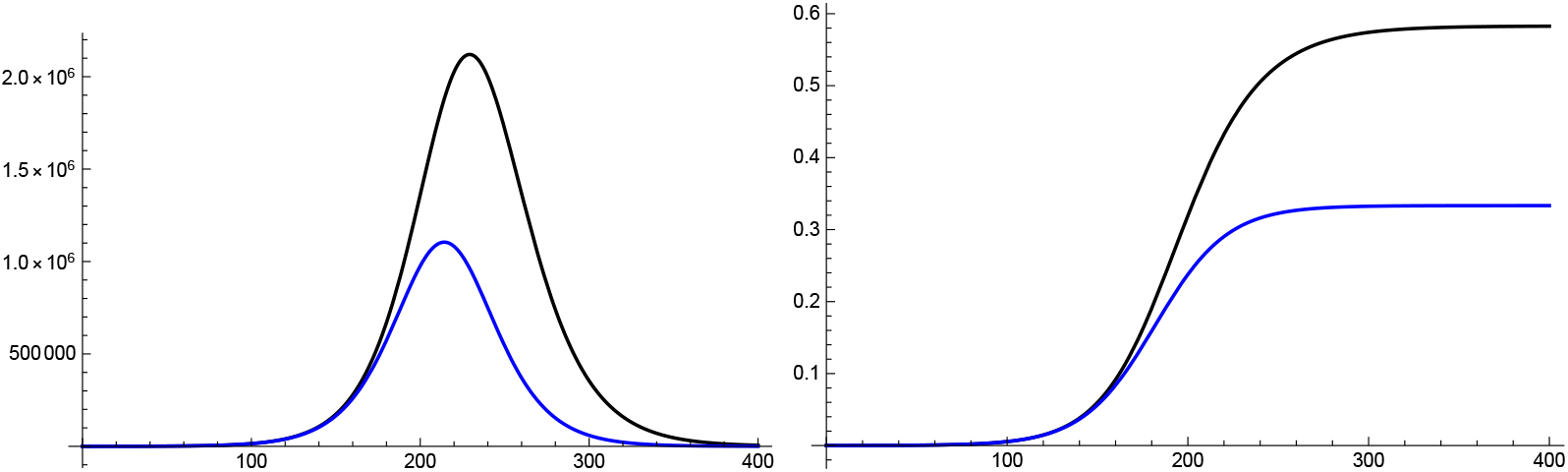
Comparison of SIR (black) and dSIR (blue) for *s*(*t*) ≈ 1 and constant coefficients with identical exponential increase and population size *N* = 80 M. Left: Number of infected *I*(*t*). Right: *H*(*t*)*/N* with *H*(*t*) total number of infected up to time *t*. Parameter values: *N* = 8 M, *I*_0_ = 1 k, *α* = *η* = 0.15, *β* = 0.1; *p* = 8.11 for dSIR.

**Figure 4.**
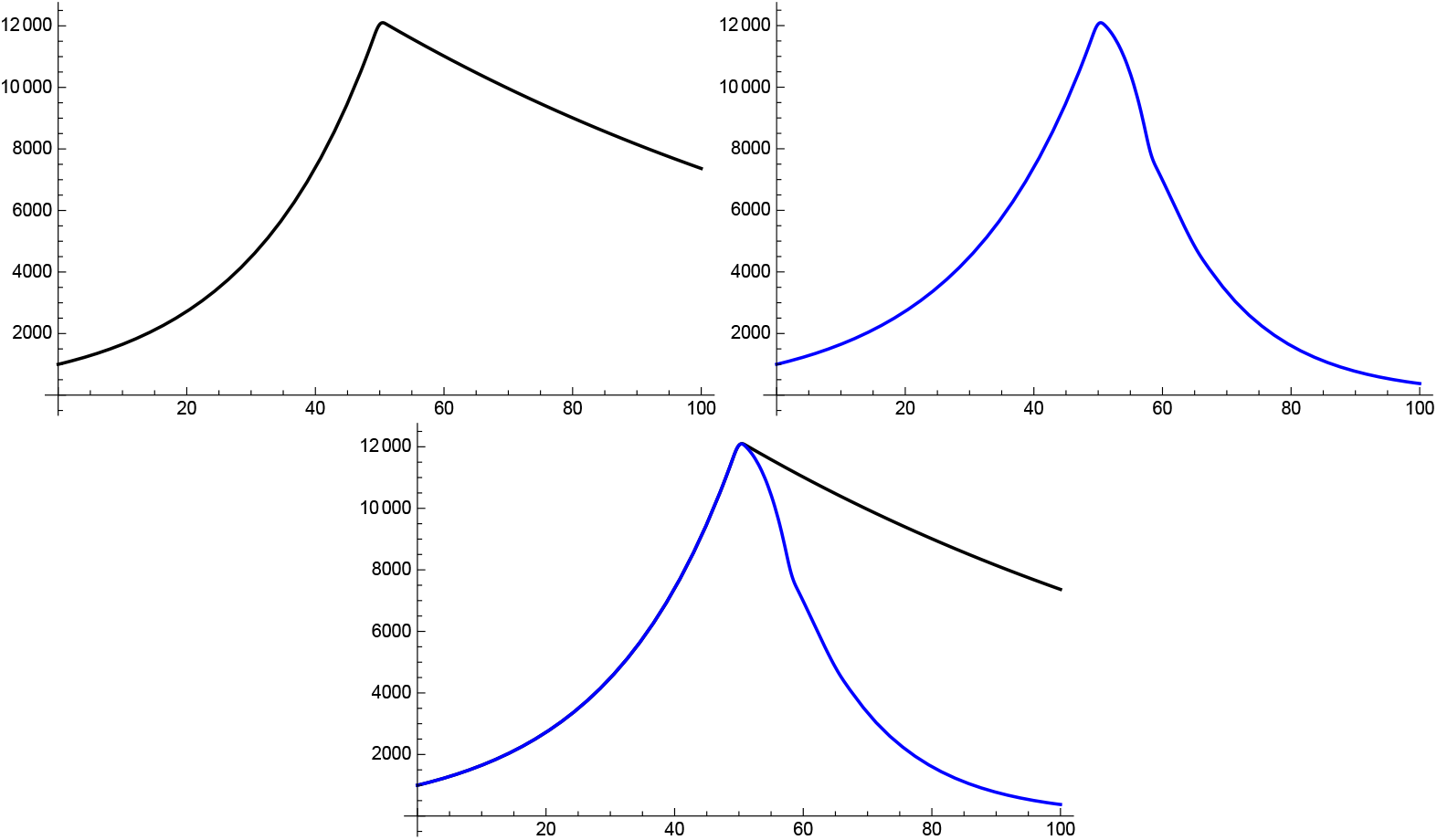
Comparison of *I*(*t*) for SIR (top left) and dSIR (top right) for *s*(*t*) ≈1 and constant coefficients for *t* ≤ 29 and *t* ≥ 31 with identical exponential increase in the initial upswing. Reduction of reproduction rate by 40 % in both cases. Bottom: SIR black, dSIR blue. Parameter values: *N* = 8 M, *I*_0_ = 1 k, *α*_1_ = *η*_1_ = 0.15, *β*_1_ = *β*_2_ = 0.1, *α*_2_ = *η*_2_ = 0.09, *β*_2_ = *β*_1_; dSIR *p* = 8.11.

**Figure 5.**
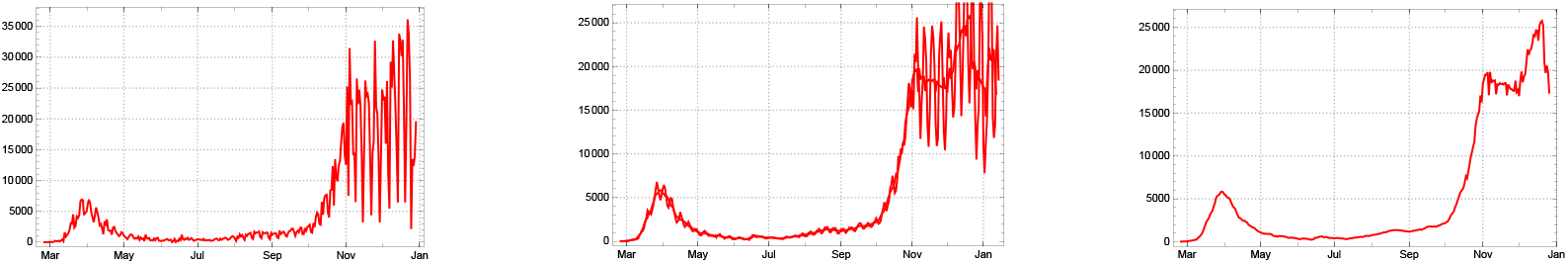
Acknowledged cases *Â*_*new*_(*k*) for Germany (left), sliding 3-day and 7-day averages *Â*_*new*,3_(*k*), *Â*_*new*,3_(*k*) (middle) and *Â*_*new*,7_(*k*) (right)

**Figure 6.**
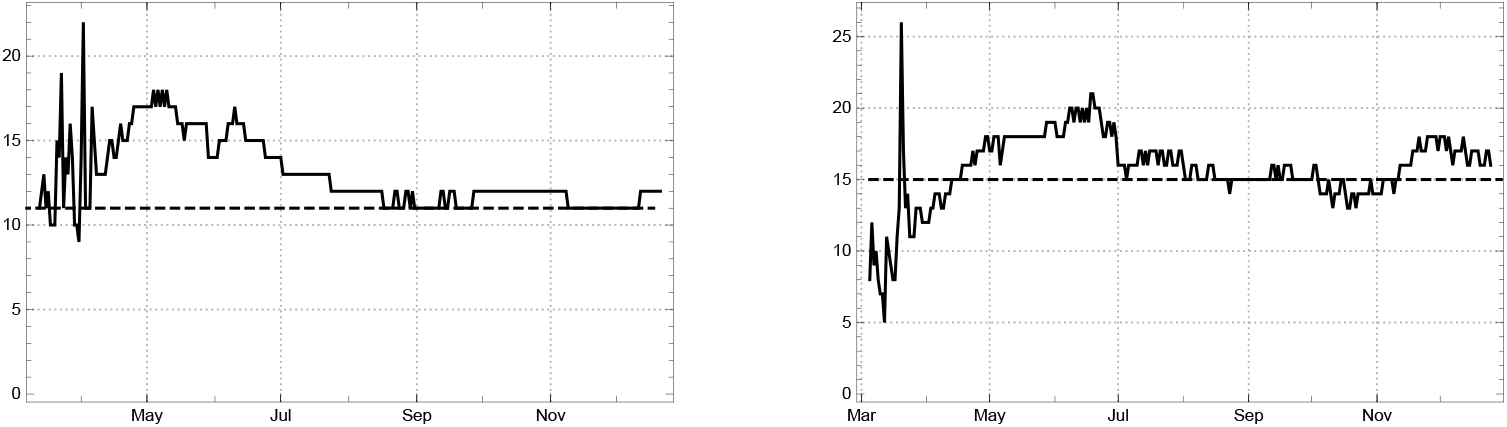
Daily values 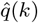 for the mean time of being statistically counted as an actual case for India (left) and Germany (right)

This has an interesting consequence for a situation in which a high rate of immunity is achieved either by “herd” effects or by vaccination. According to a simple SIR model with constant reproduction number *ρ* = 1.5 and a population of 80 million people (like in Germany) a little bit more than 0.6 80 = 24 million people would have to be infected or vaccinated to achieve herd immunity, whereas according to the delay model “only” about 0.3 80 = 12 million have to be infected (see fig. 3). The difference corresponds to the fact that equal initial exponential growth is related to different reproduction rates in the two models of the example given: 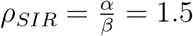 and *ρ*_*d−SIR*_ = *p η*≈ 1.2. Such a difference matters because, according to the plausibility arguments given above, the delay model may very well be more realistic than the SIR model for Covid-19.

Next we discuss the differences between the SIR model and the delay SIR model during a time when *s* is still approximately equal to 1, both have constant reproduction numbers, and *α* = *η* like above. Moreover we assume that the initial growths functions of both models are approximately identical to the same exponential function (because both are designed to modelling the same growth process).

In reality one observes longer periods in the data where the reproduction number is approximately constant until it changes in a short transition period to a new approximately constant value. Such changes may be due to containment measures (non-pharmaceutical interventions) imposed by governments, which influence the contact rate *κ*(*t*). In the next graphics we show the effect of such a change for both models.

In figure 4 we let the dSIR and the SIR curves start with identical exponential functions based on constant reproduction numbers. Then we lower the reproduction rate by 40 percent within three days. As expected the SIR curves have a cusp since one exponential function jumps into another, whereas the dSIR equation due to the delay character shows a slightly smoother transition. The second and more dramatic effect is that a similar phenomenon like in the long term comparison can be observed: the SIR solution is far above the dSIR solution. The reason seems to be the same, the different assumptions made by the two approaches about the decay of the strength of infection.

These considerations show that the choice of the model may result in important differences for the medium and long range development of the epidemic. We have given arguments why we consider the delay SIR model more realistic for Covid-19 than the standard SIR approach.

But also the delay SIR model has defects when comparing it to the data. The reason is that in reality it is not the case that an infected person gets infectious the same day, and also it takes some time until an infectious person shows symptoms. This speaks in favour of the delay SEPAR approach. In part II we apply the SEPAR_*d*_ model to data of selected countries. Here the model shows its high quality. Since data about the dark sector are insecure we check how much the dark sector influences the overall dynamics of the epidemic in the discussed countries up to the present (until the end of 2020) and choose the dark factor of the model on the basis of the analysis and given estimates for the respective countries.

## PART II: Applications of the SEPAR model

### 2. Determining empirical parameters for the model

#### 2.1. JHU data

##### The basic data sets (JHU)

The worldwide data provided by the Humdata project (Humanitarian Data Exchange) of the *Johns Hopkins University* provides data on the development of the Covid-19 pandemic for more than 200 countries and territories.^2^ The data are compressed into 3 basic data sets for each country/territory

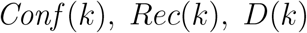

where *Conf* (*k*) denotes the total number of *confirmed cases* until the day *k* (starting from January 22, 2020), *Rec*(*k*) the number of reported *recovered* cases and *D*(*k*) the number of reported *deaths* until the day *k*. The last two entries can be combined to the number of *redrawn* persons of the epidemic, captured by the statistic,

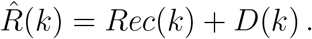

Empirical quantities derived from the JHU data set will be endowed with a hat, like 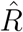, to distinguish them from the corresponding model quantity, here *R*.

The (first) differences of *Conf* (*k*) encode the daily numbers of *newly* reported and *acknowledged* cases:

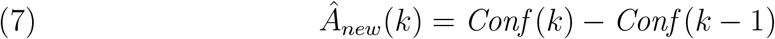

The other way round, the number of confirmed cases is the complete sum of newly reported ones, and may be considered as the total number of acknowledged cases

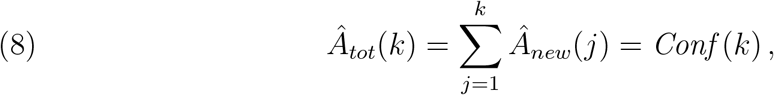

while the difference

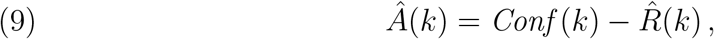

is the empirical number of *acknowledged*, not yet redrawn, *actual* cases. Some authors call it the number of “active cases”;^3^ but this is misleading because the phase of effective infectivity is usually over as soon as an infection is diagnosed and the person is quarantined.

The number *Rec*(*k*) of recovered people is often reported with much less care than the daily new cases and the deaths. By this reason the recorded number of redrawn, 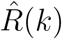, may be heavily distorted, with the result that neither itself nor the derived numbers *Â* (*k*) can be be taken at face value. The most reliable *basic data* remain therefore

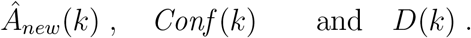

Even *Â*_*new*_ (*k*) has its peculiarities due to the weekly cycle of reporting activities. In this paper we abstain from discussing mortality rates and consider the first two data sets of the mentioned three only. 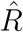 and *Â* play an important role for a complete image of an epidemic, but they are reliable only for a few countries; for the majority of countries they have to be substituted or complemented by more adequate quantities derived from the basic data (see eq. 12).

##### Smoothing the weekly oscillations of Â_new_

For all countries the reported number of daily new infections shows a characteristic 7-day oscillation resulting from the reduction of tests over weekends and the related delay of transmission of data. A 3-day sliding average suppresses fluctuations on a day-to-day scale and shows the weekly oscillations even more clearly.

In some countries these oscillations are corrected for transmission delay by central institutions,^4^ but such corrections are not implemented in the JHU data. A simple method for smoothing the weekly oscillations consists in using sliding *centred* 7-day averages:

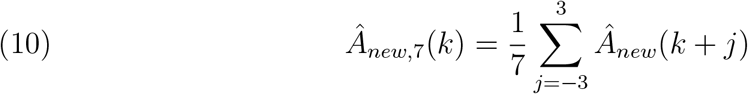

and similarly for the centred 3-day average *Â*_*new*,3_(*k*). Note that in order to avoid a time shift effect which would arise from using a purely backward sliding average, we use a sliding average over 3 days forward and 3 days back. For most countries this suffices for carving out the central tendency of the new infections quite clearly (fig. 5).

For some countries already the *daily* fluctuations of *Â*_*new*_are extreme. The French data even indicate *negative* values for *Â*_*new*_ for certain days, although this ought to be excluded by principle. Such effects indicate a highly unreliable system of data recording and transmission; they may be due to ex post corrections of earlier exaggeration of transmitted numbers. But even under such extreme conditions the sliding 7-day average leads to reasonable information on the mean motion of the new infections, so that we don’t have to exclude such countries from further consideration (sec. 3.1).

#### 2.2. Data evaluation

##### The “actual” cases in the statistical sense

The difference 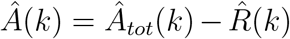 (8) can in principle be considered as an expression for the number of *actual* cases; but it is corrupted by the fact that the number of daily recovering *Rec*(*k*) is irregularly reported. For a critical investigation of this number we start from the truism that any actually infected person recorded at day *k* has been counted among the *Â*_*new*_ (*l*) at some earlier day, *l*≤*k*. The smallest number 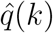 of days preceding *k* (including the latter) necessary for supplying sufficient large numbers of infected *Â* (*k*),

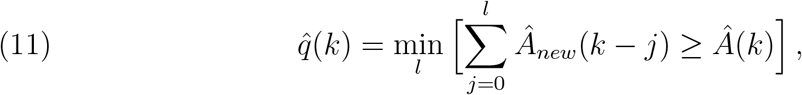

is a good indicator for the mean time of sojourn in the collective of infected which are recorded as “actual cases”. As long as the number of severely ill among all infected persons is relatively small and the time of severe illness well constrained, we may expect that the mean time of actual illness does not deviate much from the time of prescribed minimal time of isolation *q*_*min*_ for infected persons. In the case of Covid-19 *q*(*k*) surpasses *q*_*min*_≈14 only moderately for India, Germany, Switzerland etc. (fig. 6). For many other countries 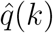 behaves differently. It starts near the time of quarantine or isolation but increases for a long time monotonically with the development of the epidemic, before often – although not always – it starts to decrease again after the (local) peak of a wave has been surpassed. This is the case, e.g., for Italy and the US; in the last case the deviation is extreme, 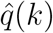 surpasses 100 and shoots up a little later (see fig. 7)

**Figure 7.**
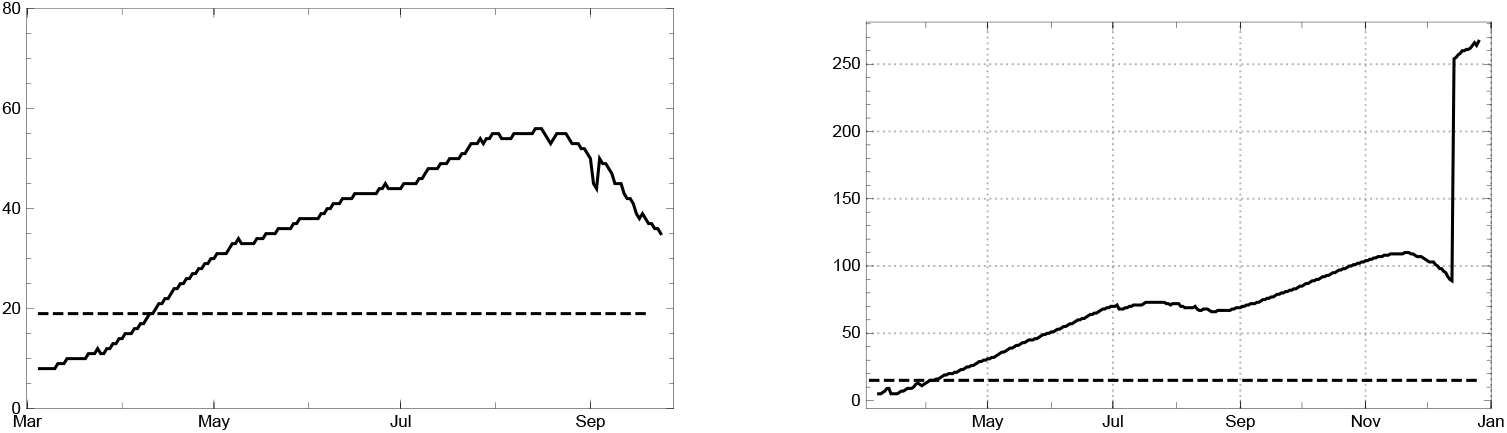
Daily values of the mean time 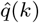 of being statistically counted as actual infected for Italy (left) and USA (right)

This effect cannot be attributed to medical reasons; the major contribution rather results from the unreliability of the statistical book keeping: With the growing overload of the health system, the time of recovery of registered infected persons is being reported with an increasing time delay, sometimes not at all (e.g., Sweden, UK). The difference 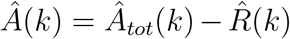 gets increasingly confounded by the lack of correctness in the numbers *Rec*(*k*). In these countries it is an expression of the number of *“statistically actual”* cases only with, at best, an indirect relation to the real numbers of people in quarantine or hospital.

The information gathered for Covid-19 proposes the existence of a stable mean time *q* of isolation of infected persons (including hospital) for long periods in each country. It is usually a few days longer than the official duration of quarantine prescribed by the health authorities. Given *q*, the sum

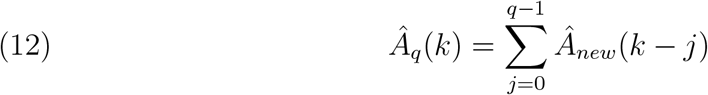

can be used as an estimate of the number of infected recorded persons who are in isolation or hospital at the day *k*. Here we do not use 7- or 3-day averages, because the summation compensates the daily oscillations anyhow. The accordingly corrected number of redrawn 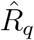 is of course given by

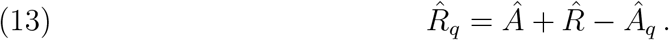

Figure 8 shows *Â*(*k*), *Â*_*q*_ (*k*) for the USA (with *q* = 15). It demonstrates the difference between *Â*(*k*) (dark blue) and *Â*_*q*_ (*k*) (bright blue) and shows that *Â*_*q*_ (*k*) is a more reliable estimate of actually infected than the numbers *Â*(*k*) (the “active cases” of the Worldometer).

**Figure 8.**
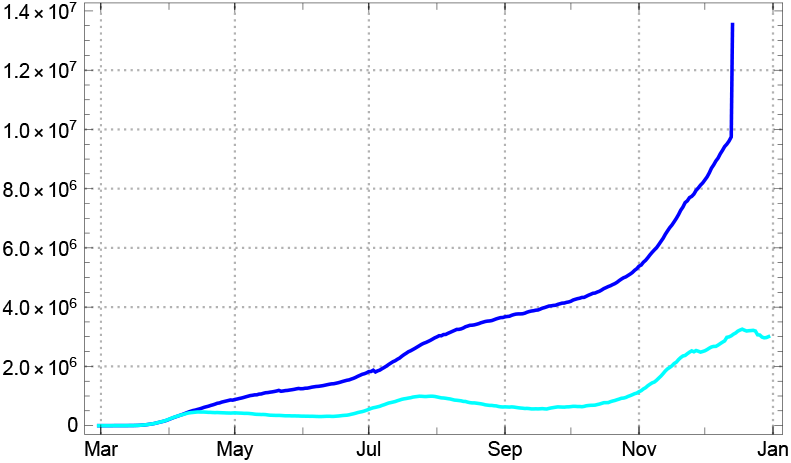
*Â*(*k*) (dark blue), *Â*_*q*_ (*k*) (bright blue) for the USA (*q* = 15)

For countries with reliable statistical recording of the recovered we find 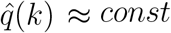 In this case we choose this constant as the value for the model *q*. For other countries one may use a default value, inferred from comparable countries with a better status of recording the *Rec*(*k*) data (i.e. *Â*(*k*) ≈ *Â*_*q*_ (*k*)).

##### Simplifying assumptions on the duration e of exposition and the duration p of effective infectivity

For Covid-19 it is known that there is a period of duration say *e* between the exposition to the virus, marking the beginning of an infection, and the onset of active infectivity. Then a period of propagation, i.e. effective infectivity, with duration *p* follows, before the infection is diagnosed, the person is isolated in quarantine or hospital and can no longer contribute to the further spread of the virus. Although one might want to represent the mentioned durations by stochastic variables with their respective distributions and mean values, we use here the mean values only and make the simplifying assumption of constant *e* and *p* approximated by the nearest natural numbers.

The Robert Koch Institute estimates the mean time from infection to occurrence of symptoms to about 4 days [16], (5.). This is divided into *e* plus the time from getting infectious to the occurrence of symptoms. According to studies already mentioned above the latter is estimated as 2 days, so as a consequence we estimate *e* = 2. In section 3 we generically use *p* = 7. We have checked the stability of the model under a change of the conventions of parameter choice inside the mentioned intervals.

*Estimate of the daily strength of infection*. As announced in part I we work with the simplified SEPAR_*d*_ model. This means that the duration in compartments *P*_*c*_ and *P*_*d*_ is equal, here denoted by *p*, and also the strength of infection is assumed to be equal: *η*_*c*_ = *η*_*d*_ = *η*. Furthermore, if *P* (*k*) = *P*_*c*_(*k*) + *P*_*d*_(*k*) there is a branching ratio *α*, which has to be estimated for each country, such that *P*_*c*_(*k*) = *αP* (*k*) and *P*_*d*_(*k*) = (1 − *α*)*P* (*k*). For every counted infected there are then

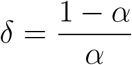

uncounted ones. We call *δ* the *dark factor*.

Once *e* and *p* are given (or fixed by convention inside their intervals) one can determine the empirical strength of infection *η*(*k*) using the model equations. Namely

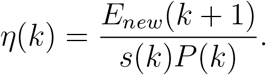

In terms of the total number of infected *H*(*k*) (see (4) and with constant *α* this is

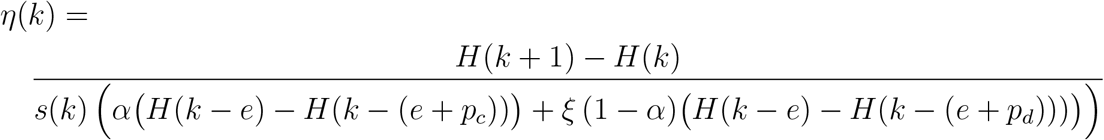

For the simplified SEPAR_*d*_ model we have *αE*_*new*_(*k*) = *A*_*new*_(*k* + *e* + *p*) and 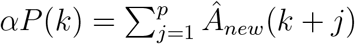. Thus *α* cancels and we obtain:

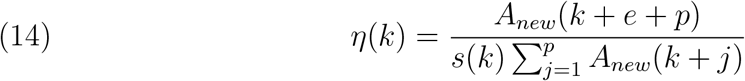

Denoting, as before, the values we obtain from the data by 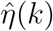 etc. we obtain

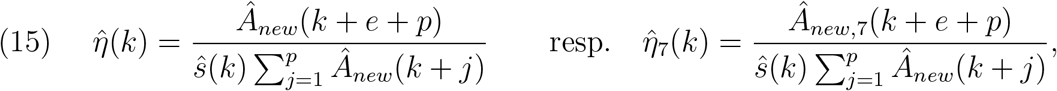

where in the second equation we work with the weekly averaged data.

An estimation of the total number of new infections induced by an infected person during the effective propagation time (and thus the whole time of illness) is then

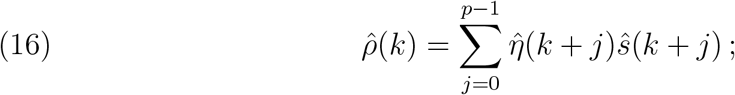

and similarly for 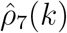. In the following we generally use the latter but write just 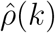. Note also that the determination of the strength of infection 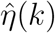 by (15) needs an estimation of the dark factor *δ* because the latter enters into the ratio of susceptibles *ŝ* (*k*),while it cancels in the calculation of the empirical reproduction rate 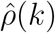 (17).

This is an empirical estimate for the *reproduction number ρ*(*k*). In periods of nearly constant daily strength of infection one may use the approximation

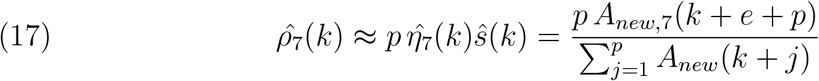

Inspection of transition periods between constancy intervals for Covid 19 shows that this approximation is also feasible in such phases of change. For *p* = 7 this variant of the reproduction number stands in close relation to the reproduction numbers used by the *Robert Koch Institut*, see appendix 4), which gives additional support to this choice of the parameter.

#### 2.3. SEPAR_*d*_ parameters

For modelling Covid-19 in the simplified dark approach we use the parameter choice *e* = 2, *p* (= *p*_*c*_ = *p*_*d*_) = 7 as explained in sec. 2.2. Where we differentiate between *p*_*c*_ and *p*_*d*_ we usually use *p*_*c*_ = 7 and *p*_*d*_ = 10. The value for *q* depends on the reported mean duration of reported infected being counted as “actual (active)” case for each country (sec. 2.1); in the following reports it usually lies between 10 and 17. For each country we let the recursion start at the first day *k*_0_ at which the reported new infections become “non-sporadic” in the sense that no zero entries appear at least in the next *e* + *p*_*d*_ days (Â_*new*_(*k*) ≠ 0 for *k*_0_≤ *k*≤ *k*_0_ +(*e*+*p*_*d*_)). For *k* ≥*k*_0_−1 the values 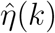 are calculated in the simplified case, *p* = *p*_*c*_ = *p*_*d*_ according to (15). Otherwise the formula above (14) has to be used.

##### Start values

For the numerical calculations we use the recursion (5), with start values given by time shifted numbers of the statistically reported confirmed cases, expanded by the dark factor, for *k* in the interval *J*_−1_ = [*k*_−1_, *k*_0_ − 1] where *k*_−1_ =*k*_0_ − 1 − (*e* + *p*_*d*_):

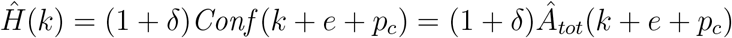

Note that the time step parametrization in the introduction/definition of the SEPAR model in sec. 1.1 works with *k*_0_ = 1.

If we set the model parameters *η*(*k*) = 0 for *k < k*_0_ − 1 and 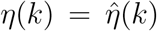 for *k* ≥*k*_0_− 1, the recursion reproduces the data exactly, due to the definition of the coefficients. Then and only then it becomes *tautological*. Already if we use coefficients 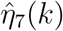 for *k* ≥*k*_0_ from time averaged numbers of daily newly reported according to eq. (15), the model ceases to be tautological. In this case the parameter *η*_0_ = *η*(*k*_0_− 1) may be used for optimizing (root mean square error) the result for the total number of reported infected *A*_*tot*_ in comparison with the empirical data (8). The model acquires conditional predictive ability, if longer intervals of constant coefficients are chosen.

One may prefer to replace *A*_*new*_ by the 7-day averages Â_*new*,7_ by introducing

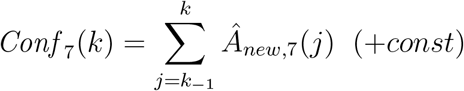

and Ĥ_7_(*k*) = (1 + *δ*) *Conf*_7_(*k* + *e* + *p*_*c*_). For constant *α*(*k*) = *α* the replacement of the Â_*new*_ in the denominator of (15) then boils down to defining

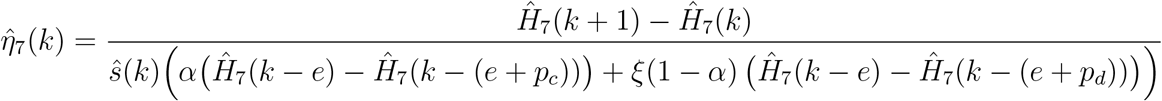

Then the model becomes tautological also for the use of daily varying 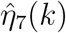 and ceases to be so only after introducing constancy intervals as described in the next subsection.

##### Main intervals

The number of empirically determined parameters can be drastically reduced by approximating the daily changing empirical infection coefficients 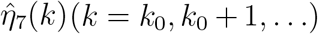 by constant model values *η*_*j*_ (1≤ *j* ≤*l*) in appropriately chosen intervals *J*_1_, … *J*_*l*_. We call them the *constancy, or main, intervals* of the model. Their choice is crucial for arriving at a full-fledged non-tautological model of the epidemic.

We thus choose time markers *k*_*j*_ (“change points”) for the beginning of such intervals and durations Δ_*j*_ for the transition between two successive constancy intervals, such that:

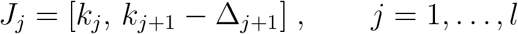

In the main interval *J*_*j*_ the *model strength of infection η*_*j*_ (this is the constant daily strength of infection during this time) are generically chosen as the arithmetical mean of the empirical values 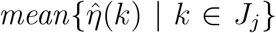. Small deviations of the mean, inside the 1 *σ* range of the 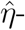fluctuations in the interval, are admitted if in this way the mean square error of the model *A*_*tot*_ can be reduced noticeably. The dates *t*_*j*_ of change points *k*_*j*_ can be read off heuristically from the graph of the 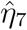 and may be improved by an optimization procedure. *k*_1_ is chosen as the first day of a period in which the daily strength of infection can reasonably be approximated by a constant. In the *initial interval J*_0_ = [*k*_0_, *k*_1_ − 1] the model uses the empirical daily strength of infection: 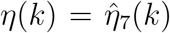 for *k* ∈ *J*_0_. In the transition intervals [*k*_*j*_ − Δ_*j*_, *k*_*j*_] the model strength of infection is gradually, e.g. linearly, lowered from *η*_*j*−1_ to *η*_*j*_.^5^

For the labelling of the days *k* there are two natural choices; the *JHU day count* starting with *k* = 1 at 01/22/2020, or a *country adapted count* such that *k*_0_ = 1, where *k*_0_ labels the first day for which the reported new infections become non-sporadic (see above). Both choices have their pros and contras; in the following we make use of both in different contexts, declaring of course which one is being used.

##### Influence of the dark sector

Proceeding in this way involves an indirect observance of the dark sector’s contribution to the infection dynamics of the visible sector. An unknown part of the counted new infections Ê_*new*_ (*k*) is causally due to contacts with infectious persons in *P*_*d*_ of the dark sector, eqs. (2) or (5)). According to estimates of epidemiologists there is a wide spectrum of possibilities worldwide, 0 ≤*δ* ≤100, for Covid-19, while we have only rough guesses for the different countries. In the following country reports we work with the simplified dark approach, *p*_*d*_ = *p*_*c*_ = *p* = 7 and *ξ* = 1 (*γ*_*c*_ = *γ*_*d*_) and use estimates for the dark factor *δ* explained in the respective country section.

## 3. Selected countries/territories

In our collection we include four small or medium sized European countries (Switzerland, Germany, France, Sweden), three large countries from three different continents (USA, Brazil, India), and a model for the aggregated data of all world countries and territories. For the first country analysed here, Switzerland, relatively reliable data on the dark sector are accessible. We take it as an example for a discussion of the effects of different assumptions on the branching ration *α*, respectively the factor *δ*, of the dark sector. For the other countries we lay open on which considerations our choice of the model the dark factor is based.

### 3.1. Four European countries; Switzerland Germany, France, Sweden

The four European countries discussed here show different features with regard to the epidemic: Switzerland and Germany have a relatively well organized health and data reporting system; the overall course of the epidemic with wave peaks in early April and in early November 2020 and a moderately controlled phase in between is is typical for most other European countries. France, in contrast, shows surprising features in the documentation of statistically recorded new infections (negative entries in the first half of the year); and Sweden has been chosen because of a containment strategy of its own. In the case of Switzerland and Germany first results of representative serological studies are available. They allow a more reliable estimate of the size of the dark sector than in most other cases. We therefore start our discussion with these countries.

#### Switzerland

The numbers of reported new infected ceased to be sporadic in Switzerland at February 29, 2020; we take this as our day *t*_0_ = 1. For the reported daily new infections (3-day and 7-day sliding averages) see figure 9. At Feb 28 recommendations for hygiene etc. were issued by the Swiss government and large events prohibited, including Basel *Fasnacht* (carnival). These regulations were already active at our day *t*_0_ = 1 (Feb 29) and explain the rapid fall of the empirical strength of infection (7-day averages) 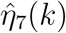 at the very beginning of our period. During the next 15 days a series of additional general regulations were taken: Mar 13, ban of assemblies of more than 100 persons, lockdown of schools; Mar 16 (*t* = 12) lockdown of shops, restaurants, cinemas; Mar 20 (*t* = 16), gatherings of more than 5 persons prohibited. This sufficed for lowering the growth rate quite effectively. The SEPAR reproduction number started from peak values close to 5 and fell down to below the critical value at March 23, the day *t*_1_ = 24 in the country count (fig. 10, left). Here it remained with small oscillations until mid May, after which it rose (we choose *t*_2_ = 85, May 23, as the next time mark), with strong oscillations until late June, before it was brought down to close to 1 in late June (*t*_3_ = 158, June 23). A long phase of slow growth (*ρ* ≈1.1) followed until mid September. An extremely swift rise of the reproduction number to values above 2 in late September (*t*_4_ = 242, Sep. 26) brought the number of new infections to heights formerly unseen in Switzerland. In spite of great differences among the differently affected regions (*Kantone*) the epidemic was brought under control at the turn to November (*t*_5_ = 272, Oct. 27).

**Figure 9.**
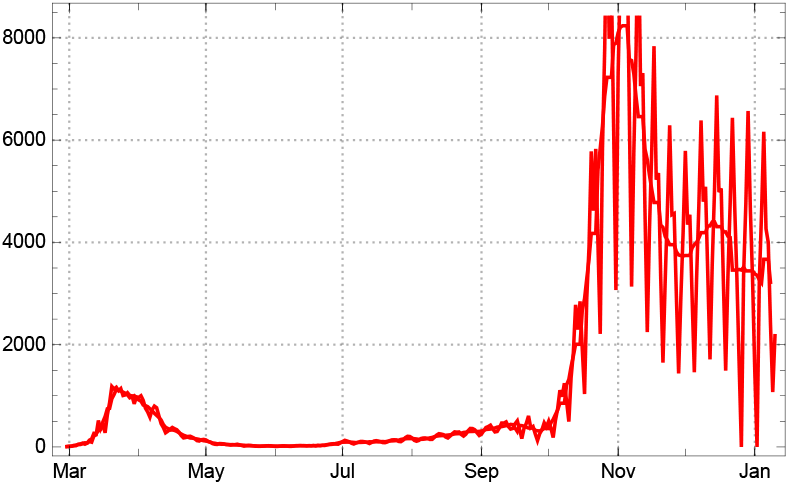
Daily new reported cases for Switzerland, 3-day sliding averages Â_*new*,3_(*k*) and 7-day averages Â_*new*,7_(*k*).

**Figure 10.**
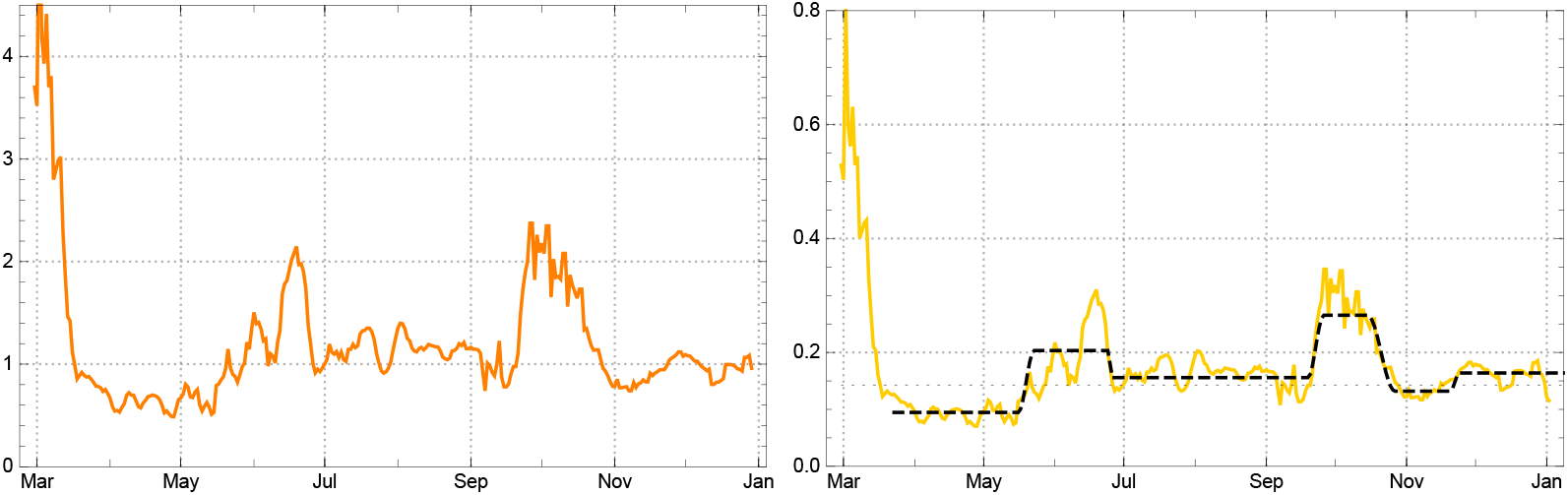
Left: Empirical reproduction rates 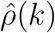 for Switzerland. Right: Daily strength of infection 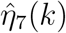 (yellow) with model parameters *η*_*j*_ (black dashed) in the main intervals *J*_*j*_ for *p* = 7 and *δ* = 2.

The empirical values of the infection strengths 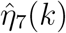 determined on the basis of the 7-day sliding averages Â_*new*,7_ depend on estimates of the dark factor *δ* (cf. eq. 15). Recent serological studies summarized in [11] conclude *δ* ≈2. We choose this value as generic for the simplified SEPAR_*d*_ model. The values of 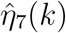, assuming a dark factor *δ* = 2, are shown in fig. 10, right. How its values are affected by different assumptions for the dark sector can be inspected by comparing with the results for *δ* = 0 and *δ* = 4 (fig. 11). The influence of the dark sector on 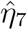 becomes visible only late in the year 2020.

**Figure 11.**
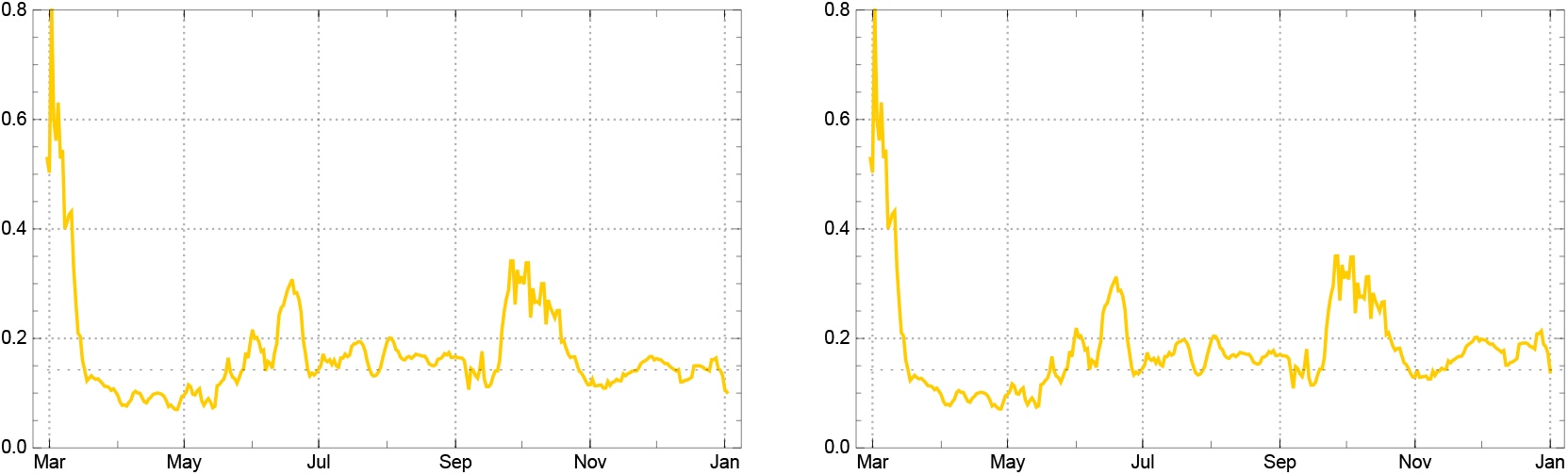
Comparison of empirical values 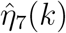 assuming dark factor *δ* = 0 (left) and *δ* = 4 (right); for comparison with generic choice *δ* = 2 see last figure.

With the time markers between different growth phases of the epidemic indicated above we choose the following main intervals for our model: *J*_0_ = [1, 22], *J*_1_ = [24, 78], *J*_2_ = [85, 117], *J*_3_ = [119, 204], *J*_4_ = [211, 230], *J*_5_ = [242, 264], *J*_6_ = [270, 321], end of data (*t*_*eod*_ = 321) at Jan 14, 2021. The model values *η*_*j*_ in the Intervals *J*_*j*_ are essentially the mean values of 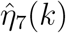 in the respective interval, where small deviations inside the 1 sigma domain are admitted if the (root mean square) fit to the empirical data Â_*tot*_ can be improved. They are given in the table below and indicated in fig. 10 (black dashed lines). (Note that *η*_0_ has no realistic meaning; it is a free parameter of the start condition for modelling on the basis of the 7-day averages 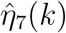, see sec. 2.3.) The reproduction rates *ρ*_*j*_ in the table refer to the beginning of the intervals; in later times the decrease of *s*(*k*) can lower the reproduction rates until the end of the intervals considerably. In the case of Switzerland the latter crosses the critical threshold 1 inside the last constancy interval (see below).

| Model $\eta_j$ and $\rho_j$ in intervals $J_j$ for Switzerland | | | | | | | |
| --- | --- | --- | --- | --- | --- | --- | --- |
| | $J_0$ | $J_1$ | $J_2$ | $J_3$ | $J_4$ | $J_5$ | $J_6$ |
| $\eta_j$ | -0.127 | 0.095 | 0.203 | 0.156 | 0.265 | 0.132 | 0.165 |
| $\rho_j$ | — | 0.66 | 1.41 | 1.08 | 1.82 | 0.86 | 1.01 |

The course of the new infections is well modelled with these values (fig. 12); the same holds for the total number of counted infected (fig. 15 below).

**Figure 12.**
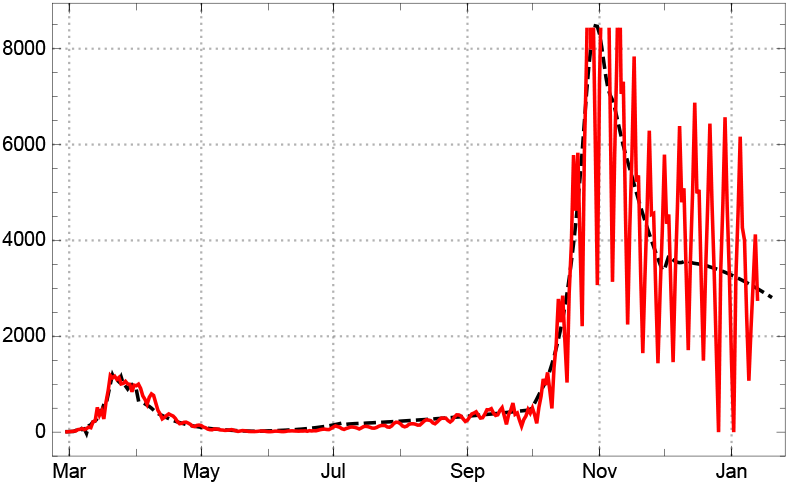
Model reconstruction for the 7-day averages of the number of new infected *A*_*new*_(*k*) (black dashed) in comparison with the empirical data, 3-day averages, Â_*new*,3_(*k*) (solid red) for Switzerland.

**Figure 13.**
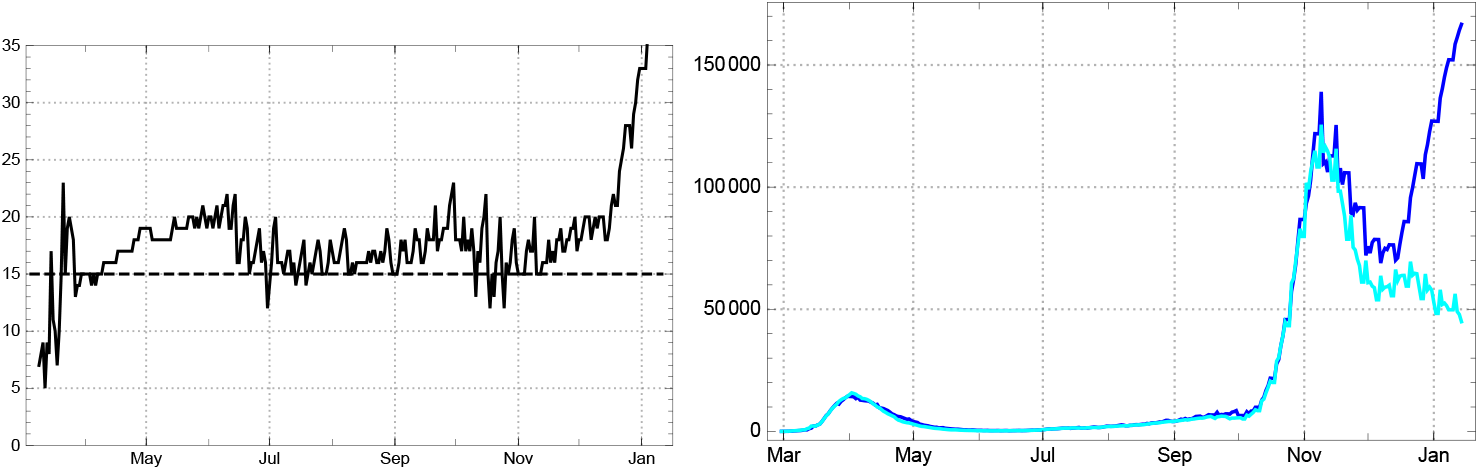
Left: Daily values of the mean time of statistically actual infection 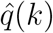 for Switzerland. Right: Comparison of reported infected Â (dark blue) and *q*-corrected number (*q* = 15) of recorded actual infected Â_*q*_ (bright blue) from the JHU data in Switzerland.

**Figure 14.**
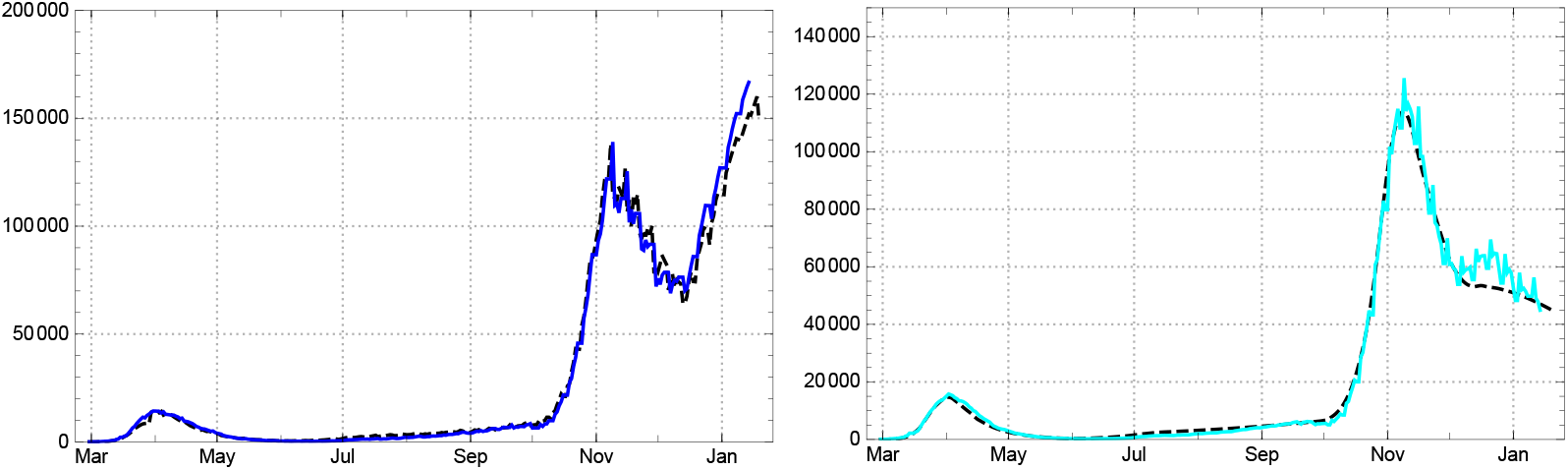
Left: Number of actual infected Â(*k*) (blue) for Switzerland, recorded by the JHU data, and model values calculated with time dependent 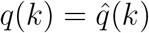 (black dashed). Right: Empirical values, *q*-corrected with *q* = 15, for statistically actual cases Â_*q*_ (*k*) and the corresponding model values *A*(*k*) (black dashed).

**Figure 15.**
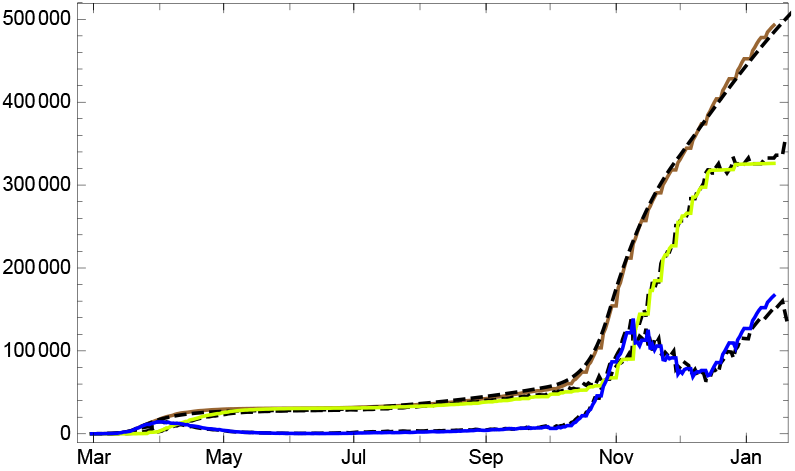
Empirical data (solid coloured lines) and model values (black dashed) in Switzerland for numbers of totally infected *Âtot* (brown), redrawn 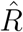 (bright green), and actual numbers Â, according to the statistic (blue).

The count of days 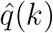, necessary for filling up the numbers of reported actual infected by sums of newly infected during the directly preceding days shows a relatively stable value, 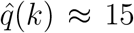, until early November. Since then the mean time of reported sojourn in the department *A* increases steeply (fig. 13, left). Initially this may have been due to a rapid increase of severely ill people during the second wave of the epidemic; but the number does not fall again with the stabilization in December 2021. A comparison of the statistically recorded actual infected Â(*k*) with the *q*-corrected one Â_*q*_ (*k*) (eq. 11) shows an ongoing increase of the first while the second one falls again after a sharp peak in early November (same figure, right). We conclude from this that from December 2020 onward the numbers of recovered are no longer reliably reported even in Switzerland.

Both quantities cam be well modelled in our approach (fig. 14), although we consider Â_*q*_ (*k*) as a more reliable estimate for actually ill persons.

On this basis, the SEPAR_*d*_ model expresses the development of the epidemic in Switzerland on the basis of only 6 constancy intervals for the parameters *η* for the whole year 2020. The three main curves of the total number of recorded infected, *A*_*tot*_(*k*), the number of redrawn *R*(*k*) and the number of diseased counted as statistical “actual” cases *A*(*k*) are shown in figure 15.

This may encourage to look at a conditional 30-day prediction for Switzerland given by the SEPAR_*d*_ model, the condition being the hypothesis of no considerable change in the contact behaviour of the population and no increasing influence of new virus mutations, i.e., a continuation of the recursion with *η*_6_, the strength of infection in the last constancy interval (fig. 16).

**Figure 16.**
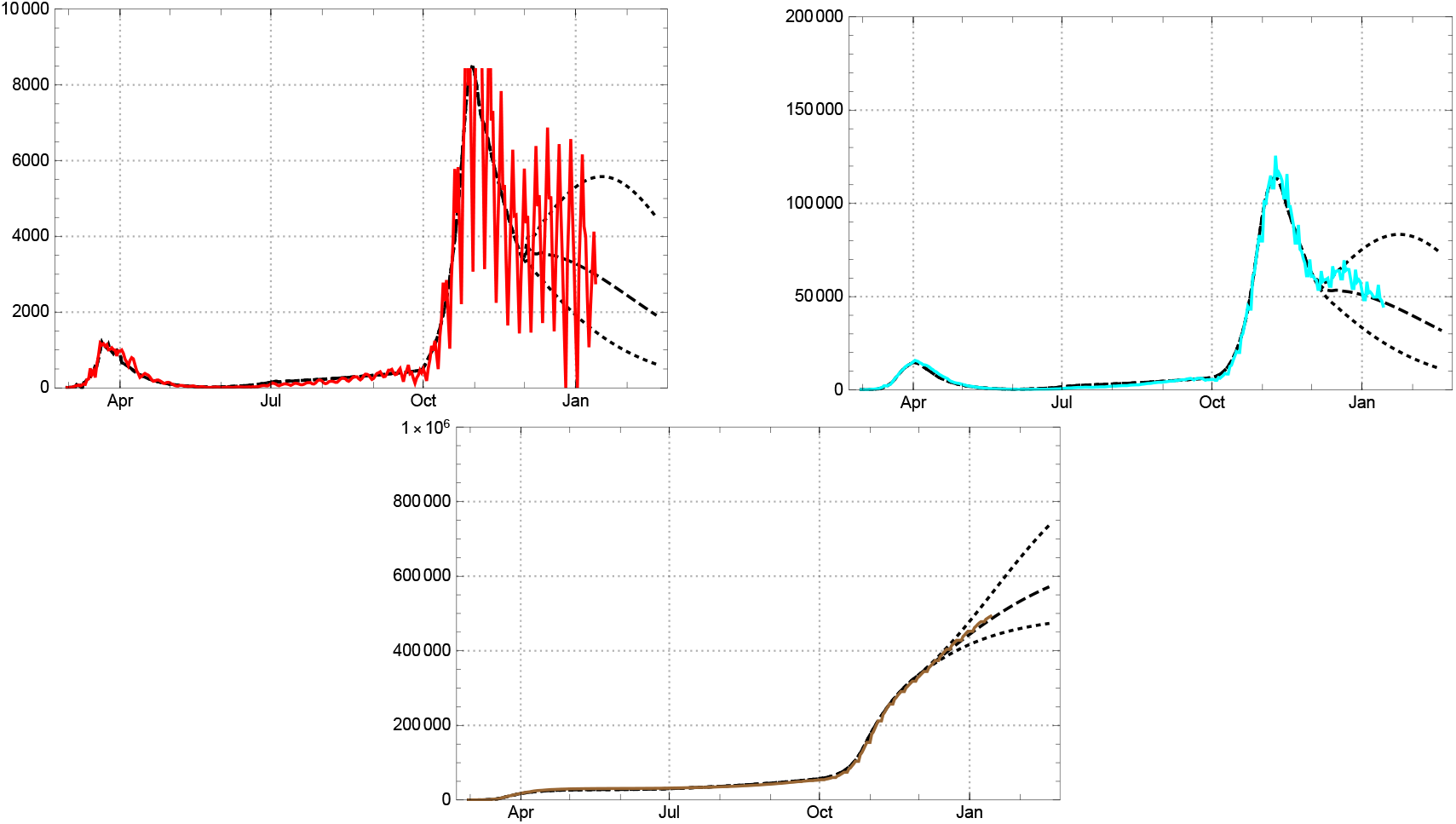
30-day prediction for *A*_*new*_, *A*_*q*_ (top) and *A*_*tot*_ (bottom) for Switzerland, assuming dark factor *δ* = 2; empirical values coloured solid lines, model black dashed (boundaries of 1-sigma region prediction dotted).

The figures show clearly that, under the generic assumption *δ* = 2 for Switzerland, the ratio of infected *s*(*k*) starts to suppress the rise of new and actual infections already in January 2021 even for the upper bound of the 1-sigma estimate for the parameter *η*_7_ (fig. 16, top). Of course the question arises what would be changed assuming different values for the dark factor. Figure 17 shows how strongly the ratio of susceptibles is influenced by the choice of *δ* already at the end of 2020.

**Figure 17.**
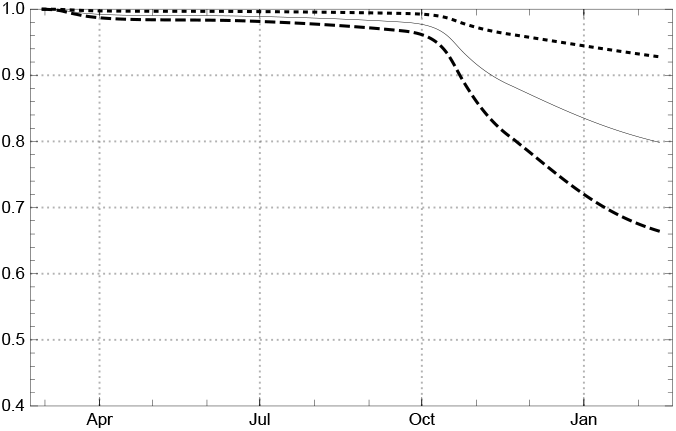
Development of ratio of susceptibles *s*(*k*) for Switzerland (model values), assuming dark factor *δ* = 0 (dotted), *δ* = 2 (solid line) and *δ* = 4 (dashed).

The results for *δ* = 0 and *δ* = 4 of the model values of reported new infected, *A*_*new*_(*k*) (black dotted or dashed as above) in comparison with the 3-day averages of the JHU data, Â_*new*,3_(*k*), is shown in figure 18. Remember that the model expresses the dynamics of 7-day averages of the newly infected.

**Figure 18.**
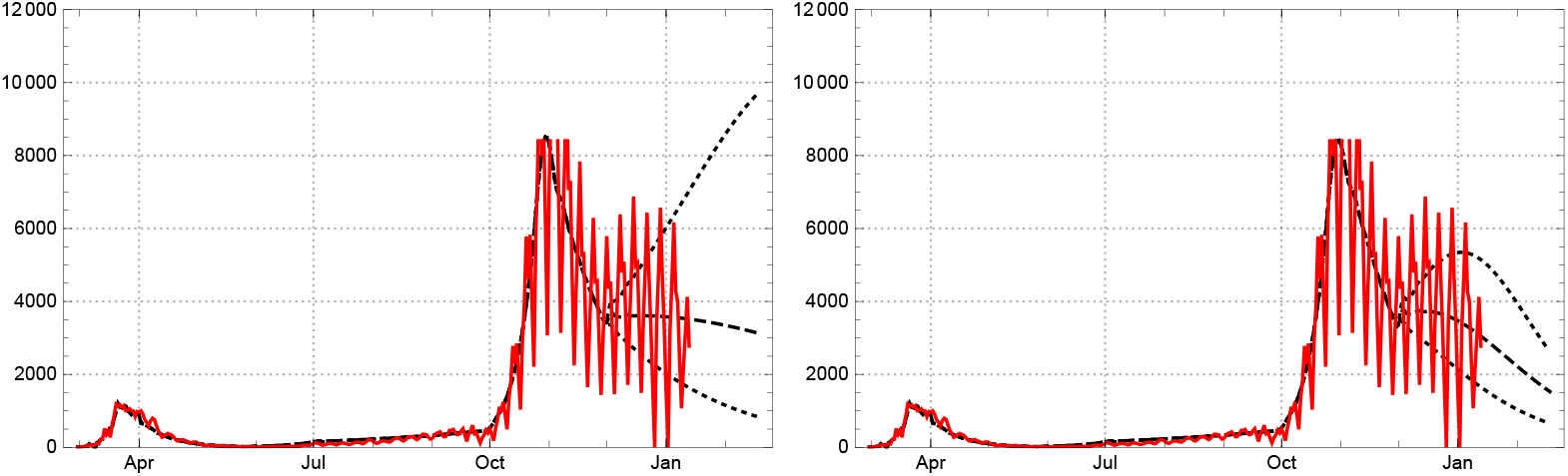
30-day prediction for *A*_*new*_ for Switzerland, assuming dark factor *δ* = 0 (left), respectively *δ* = 4 (right); empirical values Â_*new*,3_ coloured solid lines, model values black dashed (boundaries of 1-sigma region dotted).

Assuming a negligible dark sector (*δ* = 0) and the upper boundary of the 1-sigma interval of *η*_7_, the number of new infected would continue to rise deeply into the first quarter of 2021. In all other cases the effective reproduction number is suppressed below the critical value by the ratio of susceptibles *s*(*k*) already at the turn to the new year. We conclude that the role of the dark sector starts to have qualitative impact on the development of the epidemic in Switzerland already at this time.

#### Germany

The epidemic entered Germany (population 83 M) in the second half of February 2020; the recorded new infections seized to be sporadic at *t*_0_ = Feb. 25, the day *k*_0_ = 1 in our country count. With the health institutions being set in a first alarm state and public advertising of protective behaviour, the initial reproduction rate (as determined in our model approach) fell swiftly from roughly *ρ*≈ 4 to about 1.5 until mid March. At May 25 (*t*_1_ = 30) it dropped below 1 and stayed there, with an exceptional week (dominated by a huge infection cluster in the meet factory Tönnies) until late June 2020 (fig. 19 left). The peak of the first wave was reached by the 7-day averages of new infections Â_*new*,7_ at March 30; 6 days later, i.e. April 6, the local maximum for the actual numbers of reported infected Â followed.

**Figure 19.**
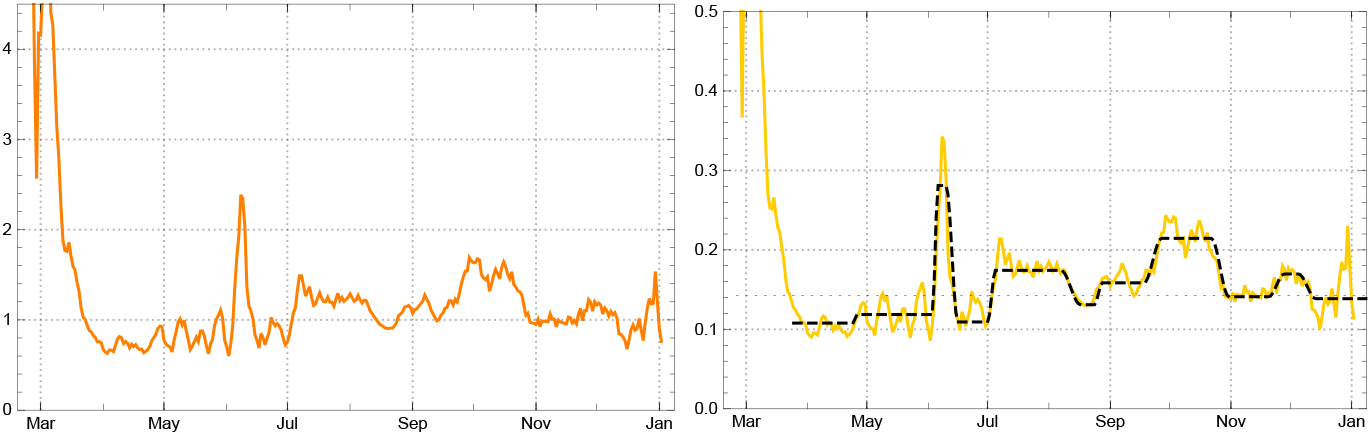
Left: Empirical reproduction rates 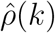 for Germany (yellow). Right: Daily strength of infection 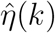 for Germany (yellow) with model parameters *η*_*j*_ in the main intervals *J*_*j*_ (black dashed); critical value 1*/p* of *η* dotted.

Serological studies in the region Munich indicate that during the first half of 2020 the ratio of counted people was about *α* = 0.25 in Germany, i.e. for each counted person there were 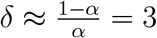 persons entering the dark sector [14]. Of course this ratio varies in space and time, for example if the number of available tests increases or, the other way round, it is too small for a rapidly increasing number of infected people. The rapid expansion of testing,in Germany during early summer seems to have increased the branching ratio to about *α* ≈0.5. In follow up investigations the authors of the Munich study come to the conclusion that during the next months the ratio of unreported infected has decreased considerably; this brought the factor *δ* down to ≈1 in early November 2020.^6^ Since the dark segment influences our model mainly through its contribution to lowering of the ratio of susceptibles *s*(*k*), it is nearly negligible in the first six months of the epidemic. We therefore simplify the empirical findings by setting *δ* = 1 for the SEPAR_*d*_ model of Germany.

Contrary to a widely pronounced assessment (including by experts) according to which the epidemic was well under control until September, the reproduction rate rose to *ρ* ≈1.3 already in July and the first half of August. Only the low level of daily new infections, reached in late May, covered up the expanding tendency and gave the impression of a negligible increase. After a short lowering interlude about mid August the rise came back in late August with *ρ* ≈1.2, before it accelerated in late September, brought the reproduction rate to about 1.5, and led straight into the second wave. Here the weekly oscillations of recorded new infections rose to an amplitude not conceived before (fig. 20). The levels of the mean daily strength of infection used in the model (proportional to the corresponding reproduction rates) are well discernible in the next figure 19, right.

**Figure 20.**
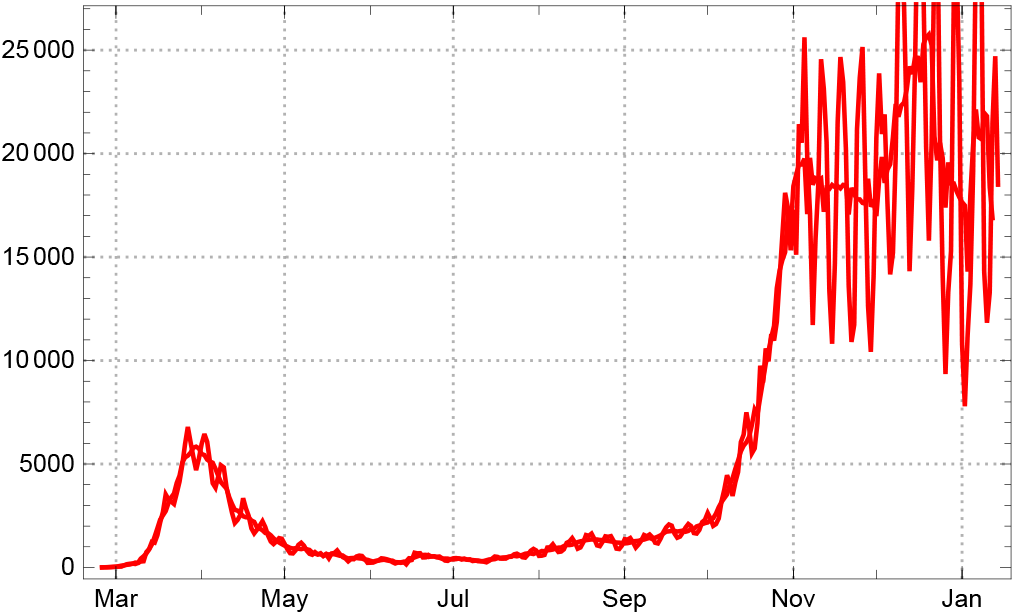
Daily new reported cases for Germany, 3-day sliding averages Â*new* (*k*) and 7-day averages Â_*new*,7_(*k*).

The dates *t*_*j*_ of the time markers *k*_*j*_ used for our model in the German case are *t*_0_ = 02/25, 2020, *t*_1_ = 03/24, *t*_2_ = 04/26, *t*_3_ = 06/06, *t*_4_ = 06/16, *t*_5_ = 07/05, *t*_6_ = 08/17, *t*_7_ = 08/28, *t*_8_ = 09/27, *t*_9_ = 10/31, *t*_10_ = 11/28, *t*_11_ = 12/14, end of data here *t*_*eod*_ = 01/15, 2021. In the country day count, where *t*_0_ = 1 (∼35 in JHU day count) the main intervals are *J*_0_ = [1, 27], *J*_1_ = [29, 59], *J*_2_ = [62, 100], *J*_3_ = [103, 108]; *J*_4_ = [113, 128], *J*_5_ = [132, 165], *J*_6_ = [175, 179], *J*_7_ = [186, 207]; *J*_8_ = [216, 241], *J*_9_ = [250, 270], *J*_10_ = [278, 285], *J*_11_ = [294, 326]. The strength of infection and reproduction numbers in the main intervals are given in the following table.

| $\eta_0$ and model $\eta_j, \rho_j$ in intervals $J_j$ for Germany | | | | | | | | | | | | |
| --- | --- | --- | --- | --- | --- | --- | --- | --- | --- | --- | --- | --- |
| | $\eta_0$ | $J_1$ | $J_2$ | $J_3$ | $J_4$ | $J_5$ | $J_6$ | $J_7$ | $J_8$ | $J_9$ | $J_{10}$ | $J_{11}$ |
| $\eta_j$ | 0 | 0.108 | 0.119 | 0.281 | 0.109 | 0.174 | 0.131 | 0.158 | 0.215 | 0.141 | 0.170 | 0.139 |
| $\rho_j$ | — | 0.75 | 0.83 | 1.96 | 0.76 | 1.21 | 0.91 | 1.10 | 1.49 | 0.97 | 1.15 | 0.93 |

With these parameters the SEPAR model reproduces (pre- or better “post”-dicts) the averaged daily new infections and the total number of reported infected well (fig. 21), while one has to be more careful for treating the reported number of actual infected *Â*.

**Figure 21.**
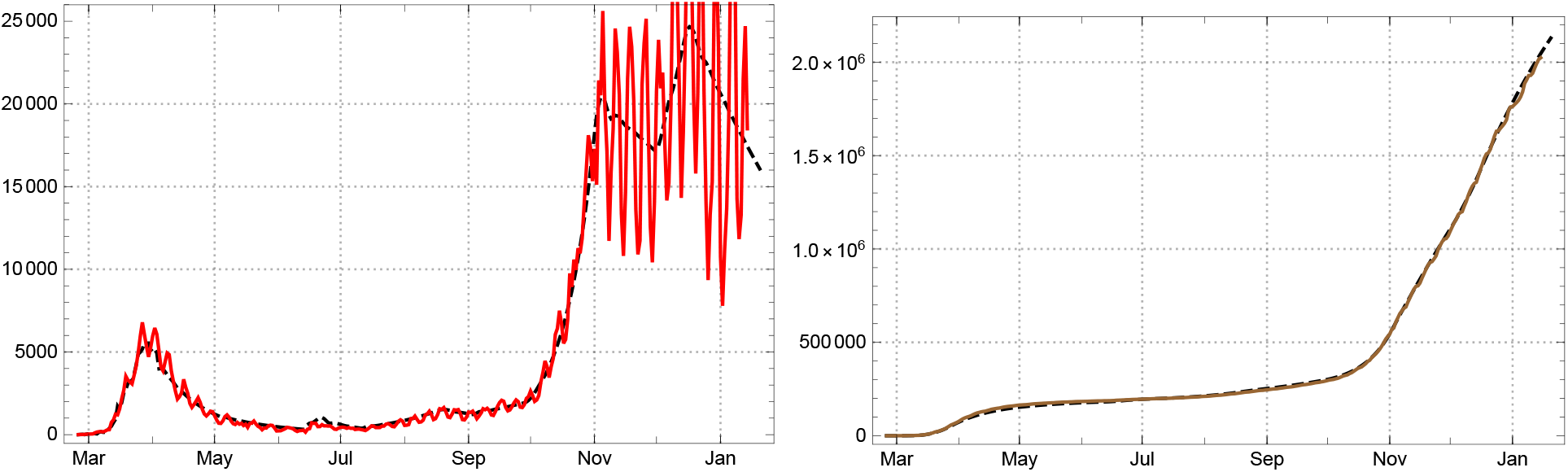
Left: Daily new reported infected (3-day averages) for Germany; empirical Â_*new*_ solid red, model *A*_*new*_ black dashed. Right: Total number of reported infected; empirical Â_*tot*_ solid brown, model *A*_*tot*_ black dashed.

If one checks the mean duration of being recorded as actual case in the JHU statistics for Germany by eq. (12) one finds a good approximation *q* ≈15 after the early phase; but the result also indicates that in May/June, and again in the second half of November, the reported duration of the infected state surpassed this value (fig. 22 left). Accordingly the empirical data Â and the corrected ones Â_*q*_ (*q* = 15) drop apart in late November (same figure, right).

**Figure 22.**
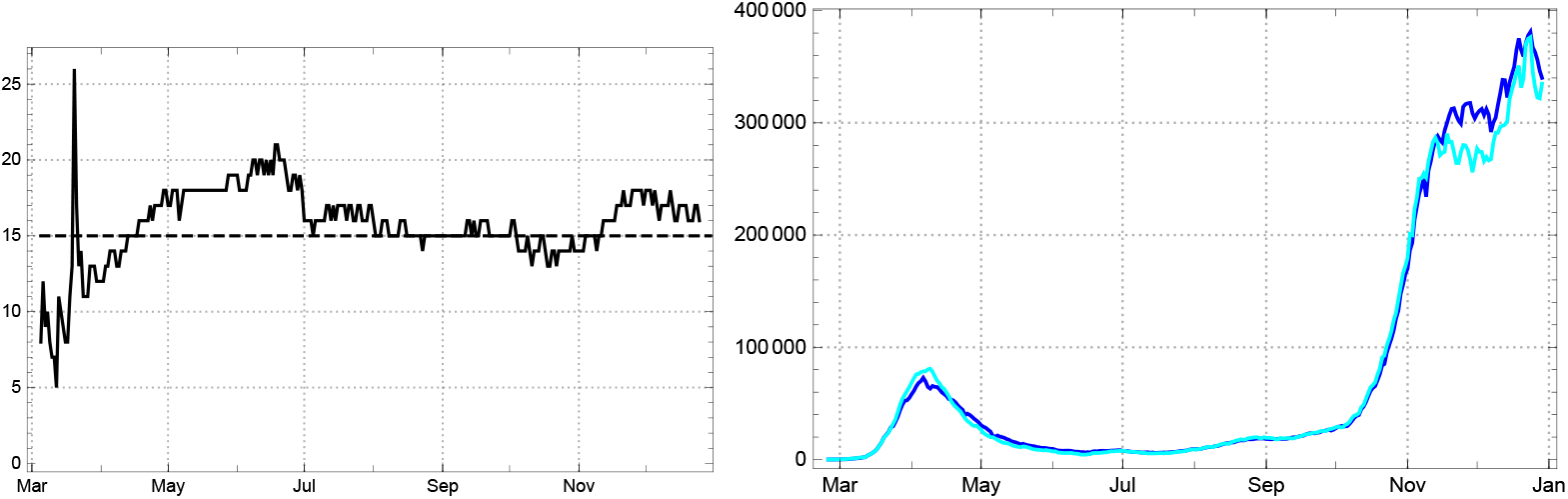
Left: Daily values of the mean time of statistically actual infection 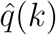 for Germany. Right: Comparison of reported infected Â (dark blue) and *q*-corrected number (*q* = 15) of recorded actual infected Â_*q*_ (bright blue) from the JHU data in Germany.

In consequence the model value for the actual infected *A*(*k*) agree with the JHU data *Â*(*k*) only if the model uses time varying values 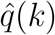 (fig. 23 left), while the *q*-corrected numbers *Â*_*q*_ (*k*) are well reproduced by the model with constant *q* = 15 (same figure, right).

**Figure 23.**
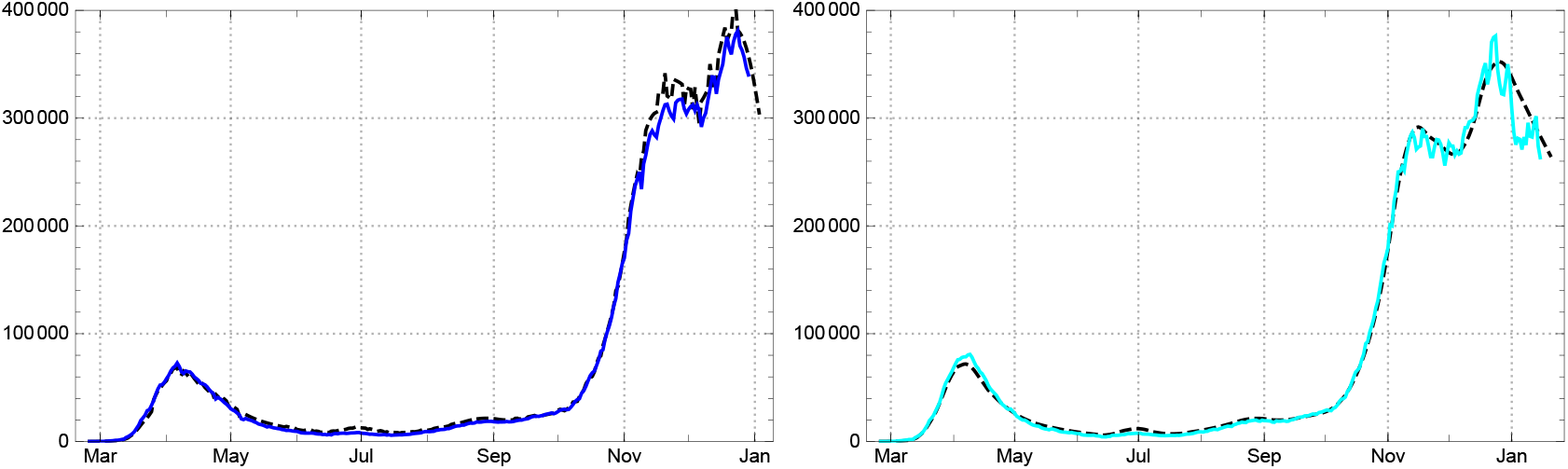
Left: Statistically actual cases Â(*k*) for Germany and the corresponding model values *A*(*k*) (black dashed) calculated with time dependent model values for *q* (see text). Right: Empirical values, *q*-corrected, for actual cases Â_*q*_ (*k*) and the corresponding model values *A*(*k*) (black dashed), *q* = 15.

As a result, the 3 model curves representing the total number of (reported) infected *A*_*tot*_, the redrawn *R* and the actual infected fit the German data well, if the last two are compared with the *q*-corrected empirical numbers *Â*_*q*_ (fig. 24).

**Figure 24.**
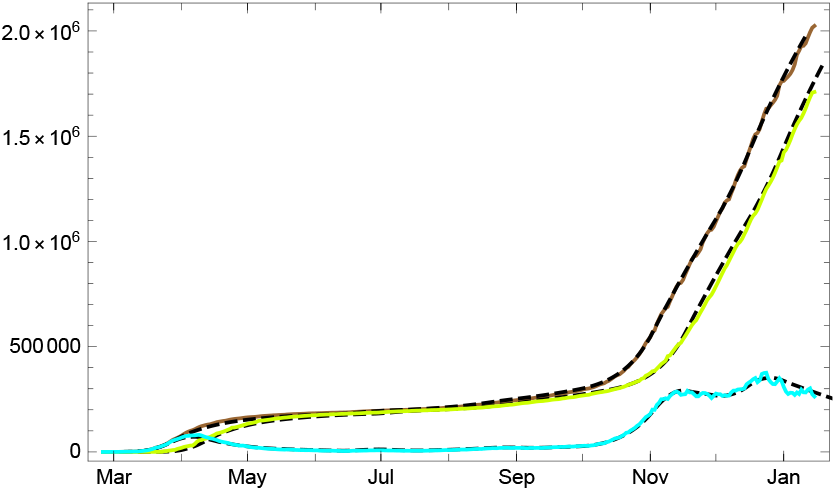
Empirical data (solid coloured lines) and model values (black dashed) for Germany: numbers of totally infected Â_*tot*_ (brown), redrawn 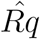 (bright green), and *q*-corrected actual numbers Â_*q*_ (bright blue).

Conditional predictions for *A*_*new*_, *A*_*q*_ and *A*_*tot*_, assuming no essential change of the behaviour, contact rates and the reproduction number from the last main interval *J*_10_ are given in fig. 25. The dotted lines indicate the boundaries of the prediction for the 1-*σ* domain for the variations of the values of 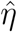; in the last main interval *J*_10_.

**Figure 25.**
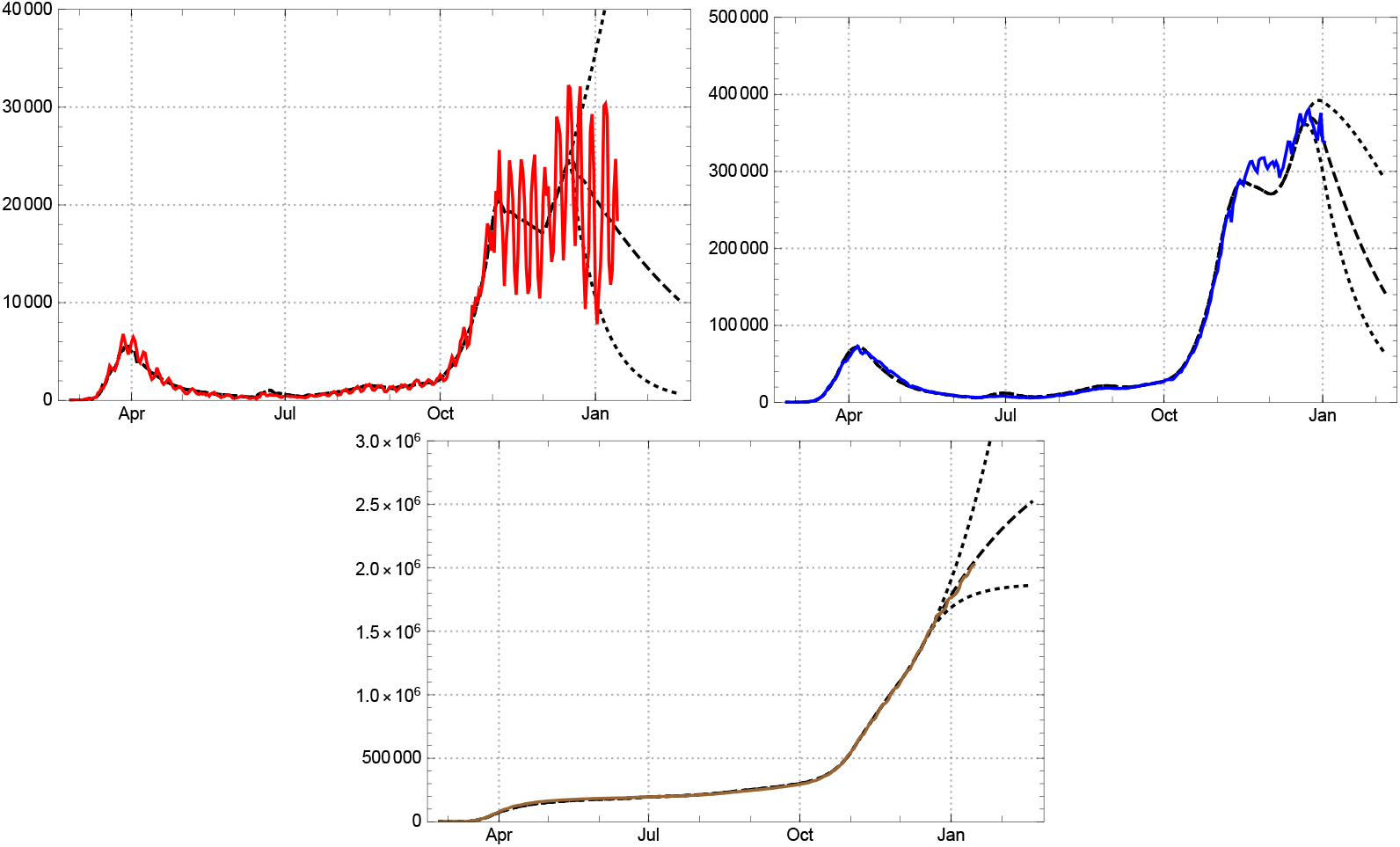
30-day prediction for *A*_*new*_, *A* (top) and *A*_*tot*_ (bottom) for Germany; empirical values coloured solid lines, model black dashed (boundaries of 1-sigma region prediction dotted).

#### France

The overall picture of the epidemic in France (population 66 M) is similar to other European countries. But the French JHU data show anomalies which are not found elsewhere: The differences of two consecutive values of the confirmed cases, which ought to represent the number of newly reported, is sometime *negative*! This happens in particular in the early phase of the epidemic (until June 2020) where, e.g.,*Â*_*new*_(58) = −2206, and there are other days with negative entries. Presumably this is due to ex-post data corrections necessary in the first few months of the epidemic. Later on the control over the documented data seems to have been improved; negative values are avoided, although null entries in *Â*_*new*_ still appear. So the French data are a particular challenge to any modelling approach. Even in this extreme case the smoothing by 7-day sliding averages works well, as shown in fig. 26.

**Figure 26.**
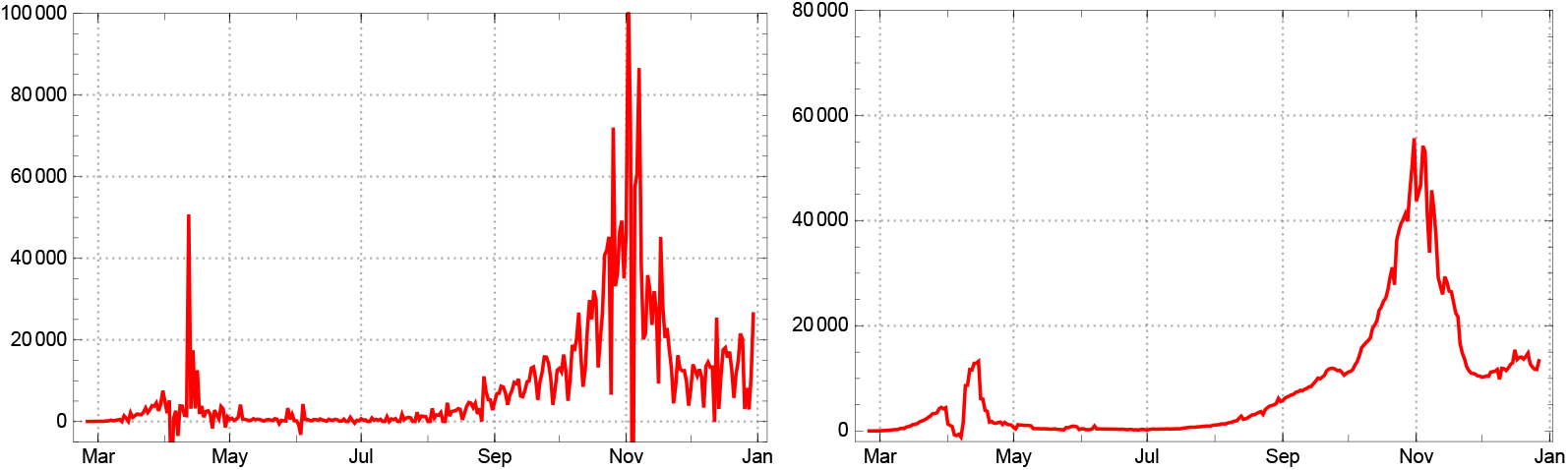
Left: Number of daily newly reported in France Â_*new*_ (*k*). Right: 7-day sliding averages of new infections Â_*new*,7_(*k*) for France.

Another surprising feature of the French (JHU) statistics is an amazing increase in the number of days which infected persons are being counted as “actual cases”. It starts close to 15, but shows a monotonous increase until late October where a few downward outliers appear, before the curve turns moderately down in early November 2020 (fig. 27, left).

**Figure 27.**
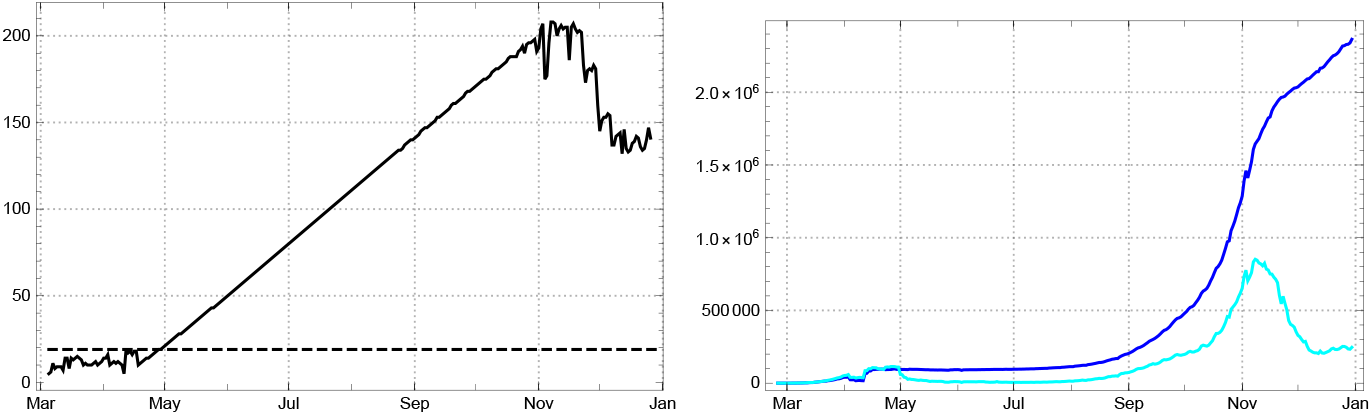
Left: Daily values of the mean time of statistically actual infection 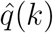 for France. Right: Comparison of reported infected Â (dark blue) and *q*-corrected number (*q* = 19) of recorded actual infected Â_*q*_ (bright blue) from the JHU data in France.

As a consequence the peak of the first wave in early April (clearly visible in the number of newly reported) is suppressed in the curve of the actual infected; Â(*k*) has no local maximum in the whole period of our report. It even continues to rise, although with a reduced slope, after the second peak of the daily newly reported, *Â*_*new*_, in early November. The decrease of the slope of Â starts shortly after this peak, accompanied by a local maximum of the *q*-corrected values for the actual infected reaches Â_*q*_ (fig. 27). Both effects seem to be due to a downturn of 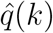. This extreme behaviour of the data cannot be ascribed to medical reasons; quite obviously it results from a high degree of uncertainty in data taking and recording in the French health system.

The reproduction numbers are shown in figure 28, left. For the determination of the daily strength of infection we have to fix a value for the dark factor. Lacking data from representative serological studies in France we assume that it is larger than in Switzerland and choose as a reference value for the model *δ* = 4. With this value the determination of the 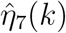 are given in figure 28, right, here again with dashed black markers for the periods modelled by constancy intervals in our approach.

**Figure 28.**
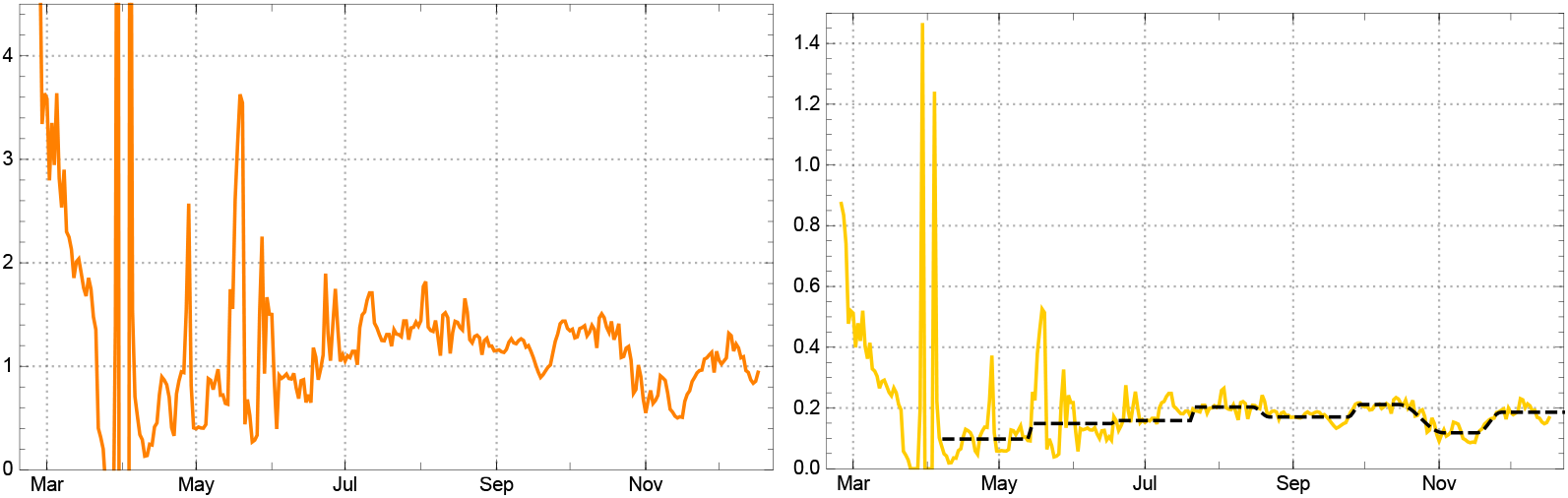
Left: Empirical reproduction rates 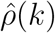 for France. Right: Daily strength of infection are shown in figure 28 for France (yellow) with model parameters *η*_*j*_ in the main intervals *J*_*j*_ (black dashed).

The markers of change times are here *t*_0_ = 02/25, 2020, *t*_1_ = 05/17, *t*_2_ = 06/15, *t*_3_ = 07/21, *t*_4_ = 08/22, *t*_5_ = 09/29, *t*_6_ =11/03, *t*_7_ =11/27, end of data *t*_*eod*_ = 12/30, 2020. In the country day count, *t*_0_ = 1 (∼35 in JHU day count), the main intervals are *J*_0_ = [1, 82], *J*_1_ = [83, 110], *J*_2_ = [112, 146], *J*_3_ = [148, 173]; *J*_4_ = [180, 213], *J*_5_ = [218, 234], *J*_6_ = [253, 267], *J*_7_ = [277, 310]. The model strength of infection *η*_*j*_ and corresponding reproduction numbers *ρ*_*j*_ for the main intervals are given in the following table.

| $\eta_0$ and model $\eta_j, \rho_j$ in intervals $J_j$ for France | | | | | | | | |
| --- | --- | --- | --- | --- | --- | --- | --- | --- |
| | $a_0$ | $J_1$ | $J_2$ | $J_3$ | $J_4$ | $J_5$ | $J_6$ | $J_7$ |
| $\eta_j$ | 2.008 | 0.149 | 0.158 | 0.203 | 0.170 | 0.211 | 0.118 | 0.186 |
| $\rho_j$ | — | 1.03 | 1.09 | 1.39 | 1.16 | 1.40 | 0.71 | 1.07 |

An overall picture of the French development of new infections and total number of recorded cases is given in fig. 29. The so-called “actual” cases are well modelled in our approach (fig.30), if the reference are the *q*-corrected numbers *Â*_*q*_ (*k*) of actual infected (or if time dependent durations *q*(*k*) (read off from the JHU data, 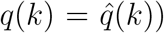 are used). For a combined graph of the 3 curves see fig. 31.

**Figure 29.**
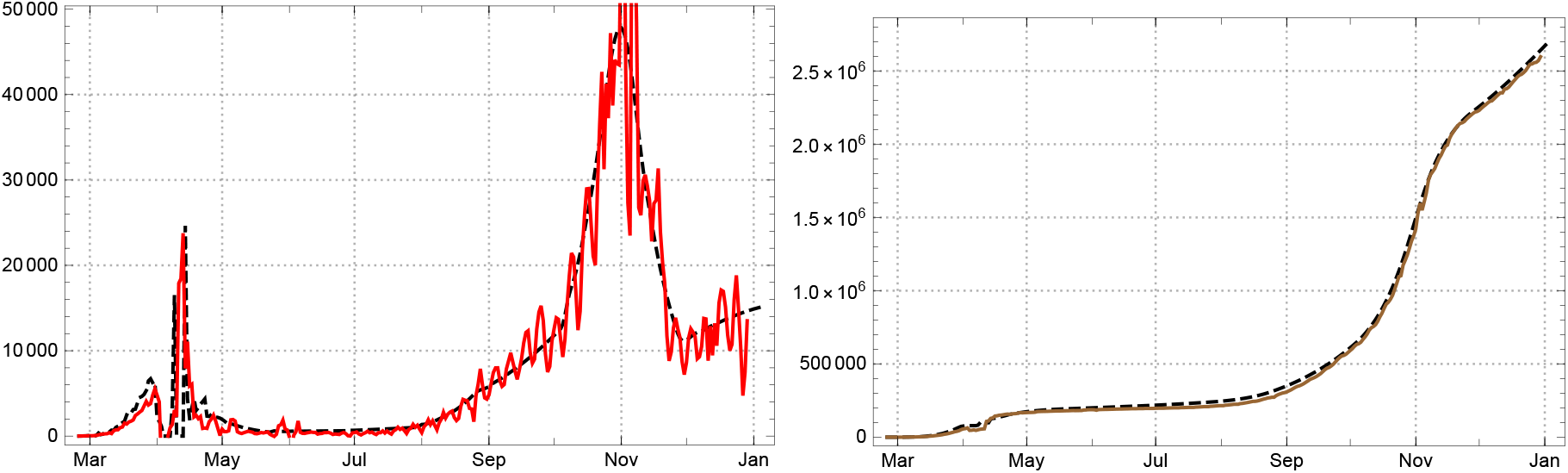
Left: Daily new reported number of infected for France (7-day averages); empirical Â_*new*_ solid red, model *A*_*new*_ black dashed. Right: Total number of reported infected (brown); empirical Â_*tot*_ solid, model *A*_*tot*_ black dashed.

**Figure 30.**
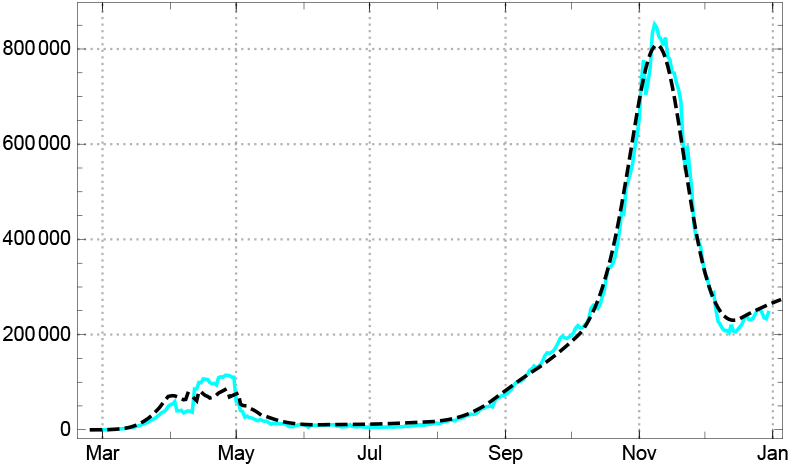
Empirical values for statistically actual *q*-corrected cases Â_*q*_ (*k*) and the corresponding model values *A*(*k*) (black dashed) for France.

**Figure 31.**
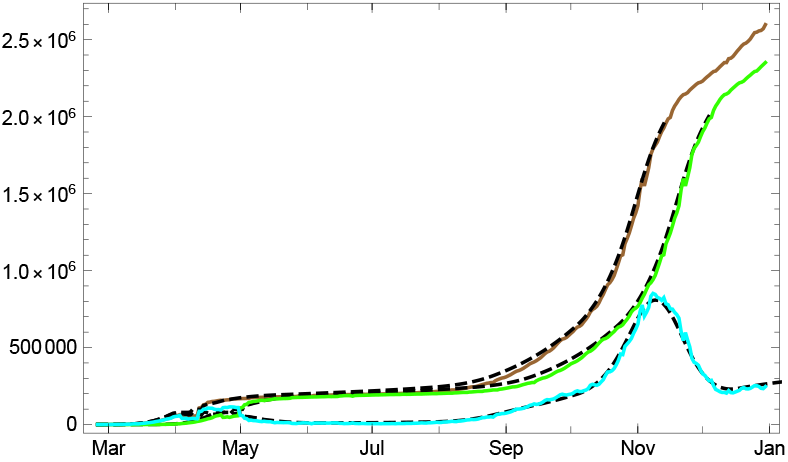
Empirical data (solid coloured lines) and model values (black dashed) in France for numbers of totally infected Â_*tot*_ (brown), redrawn 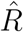 (bright green), and *q*-corrected actual numbers Â_*q*_ (bright blue).

#### Sweden

Sweden (population 10 M) has chosen a path of its own for containing Covid-10, significantly different from most other European countries. In the first half year of the epidemic no general lockdown measures were taken; the general strategy consisted in advising the population to reduce personal contacts and to go into self-quarantine, if somebody showed symptoms which indicate an infection with the SARS-CoV-2 virus. One might assume that the number of undetected infected, the dark sector, could be larger than in other European countries. As we see below such a hypothesis is not supported by the analysis of the data in the framework of our model.

Under the conditions of the country (in particular the relative low population density in Sweden) the first wave of the epidemic was fairly well kept under control, if we abstain from discussing death rates like in the rest of this paper. Once the initial phase was over (with reproduction numbers already lower than in comparable countries, but still up to about *ρ* ≈2), the reproduction rate was close to 1 for about two weeks in late June, and even lower in early July (fig. 32, right). In the second half of October 2020, however, Sweden was hit by a second wave like all other European countries. After a sharp rise of the daily new infections in early October (fig. 32, left), the Swedish government decided to decree a (partial) lockdown. By this the reproduction number, which already in late August had risen to above 1.2 and went up to 1.45 in mid October, was brought down to the former range (*ρ* ≈1), although now on a much higher level of actual infected than during the first wave.

**Figure 32.**
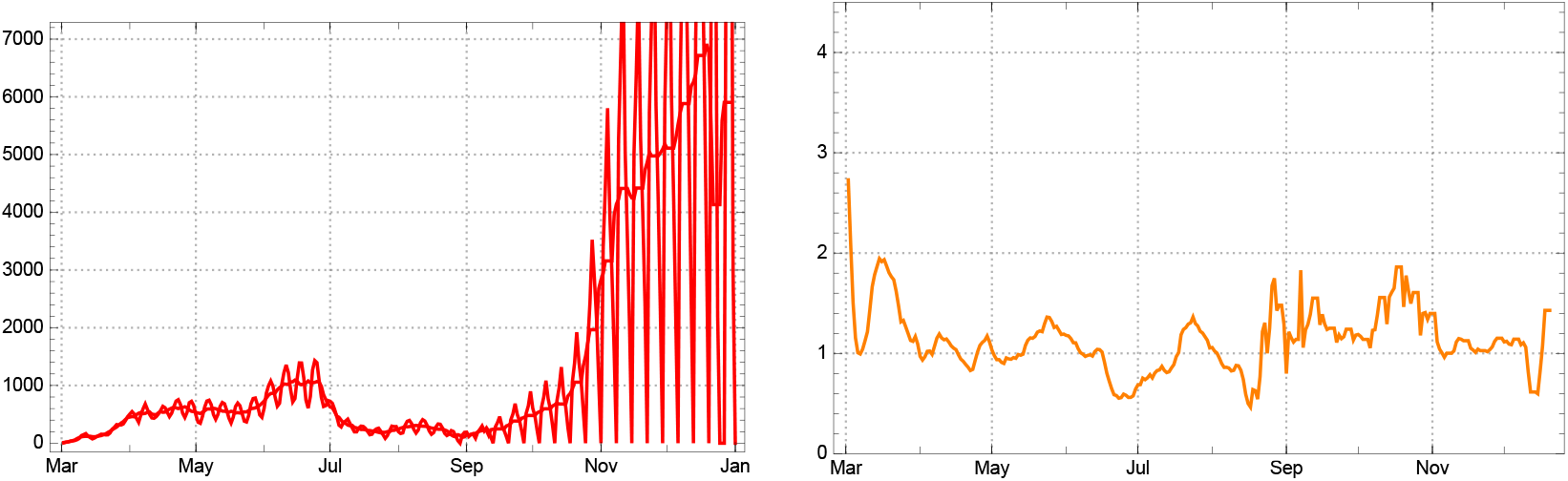
Left: Daily new reported cases Â_*new*_ (*k*) (JHU data) for Sweden and 7-day sliding averages Â_*new*,7_(*k*). Right: empirical reproduction rates 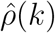 for Sweden.

The higher the assumptions for the dark sector, the larger the calculated values for 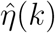 on the basis of the same data, and vice versa. Figure 33 shows the differences of 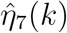 in the case of Sweden under the hypotheses *δ* = 0, 4, 15, 25.

**Figure 33.**
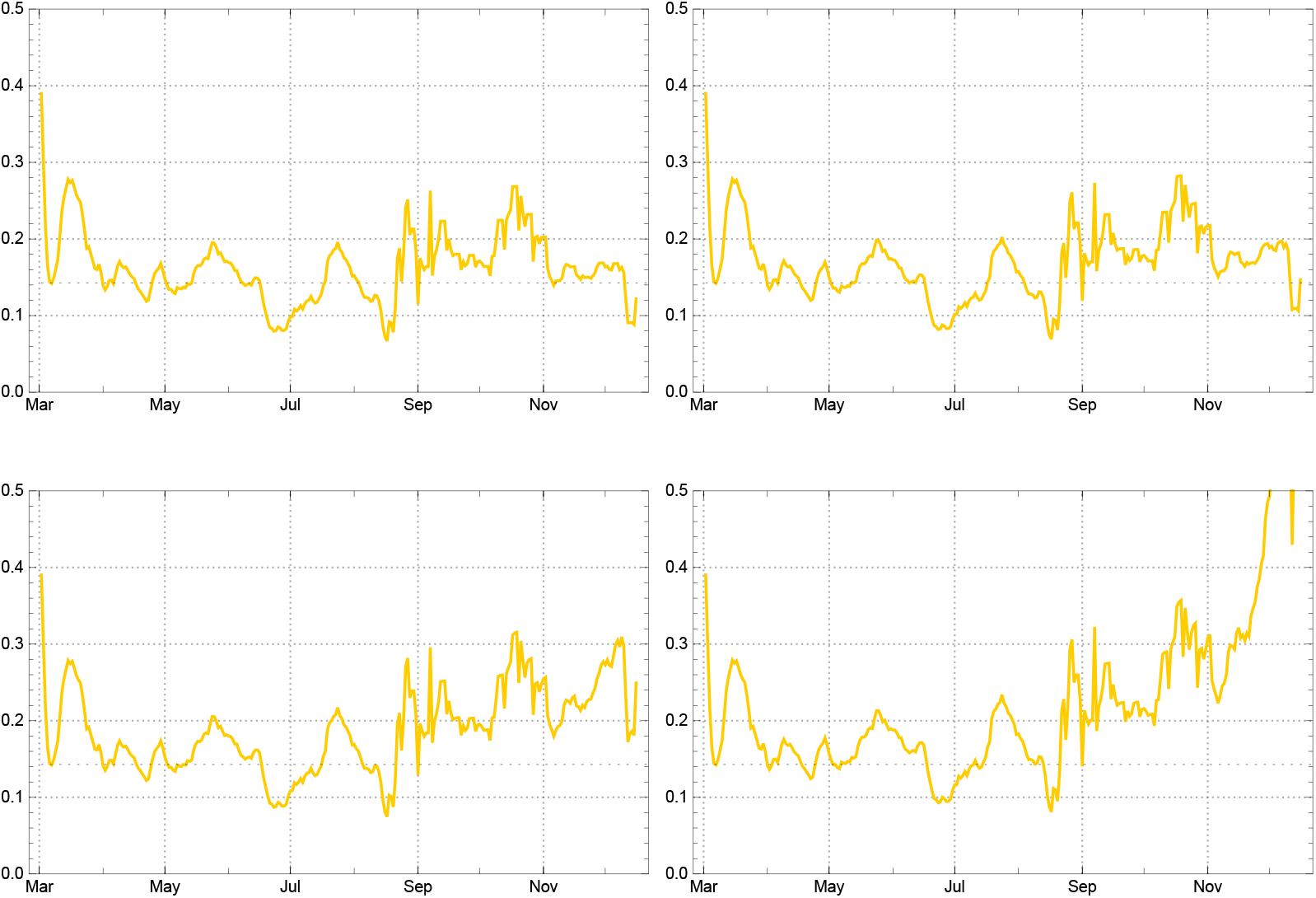
Daily strength of infection (sliding 7-day averages) 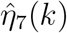 for Sweden, assuming different dark factors *δ*. Top: *δ* = 0 (left) and *δ* = 4 (right). Bottom: *δ* = 15 (left) and *δ* = 25 (right).

Until July/August 2020 the four curves show minor differences and indicate relative stable values for the infection strengths leading to reproduction rates close to 1. In September/October the values rose considerably; they went down after the October lockdown only under the assumption of a small dark sector, *δ* = 0 or 4, while for the larger dark factors *δ* = 15, 25 the values of the daily strength of infection continues to increase. This seems implausible.^7^ We therefore choose *δ* = 4 also for Sweden.

In our definition the new infections in Sweden ceased to be sporadic at *t*_0_ = 03/02, 2020, the day 41 in the JHU day count. After a month of strong ups and downs of the strength of infection, the approach of constancy intervals gains traction; with time markers of the main intervals *t*_1_ = 03/31, *t*_2_ = 05/17, *t*_3_ = 06/18, *t*_4_ = 07/17, *t*_5_ = 08/25, *t*_6_ = 10/10, *t*_7_ = 11/05, *t*_8_ = 12/12, end of data *t*_*eod*_ = 12/30, 2020. In the country count the main intervals are *J*_0_ = [1, 29], *J*_1_ = [30, 75], *J*_2_ = [77, 99], *J*_3_ = [109, 126]; *J*_4_ = [138, 169], *J*_5_ = [177, 218], *J*_6_ = [223, 214], *J*_7_ = [223, 284], *J*_8_ = [286, 303].

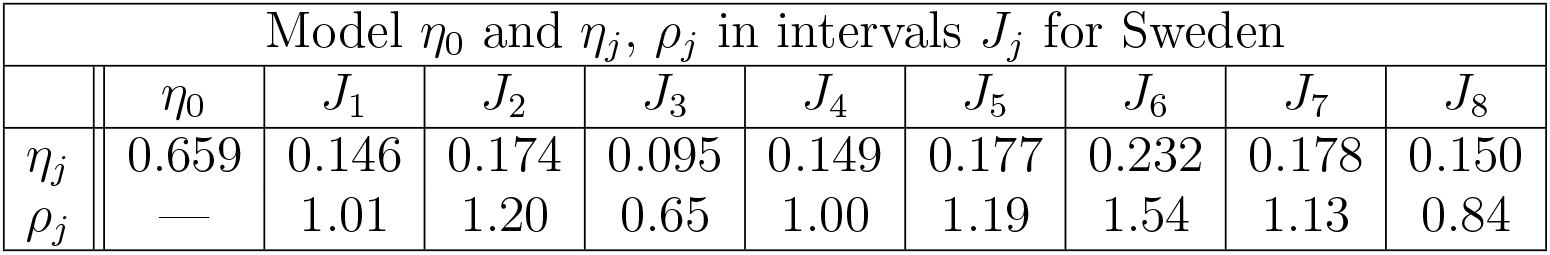

Although one might want to refine the constancy intervals, already these intervals lead to a fairly good model reconstruction of the mean motion of new infections and the total number of infected (fig. 34). Note that since early September the reported numbers of new infections show strong weekly oscillations between null at the weekends and high peaks in the middle of the week.

**Figure 34.**
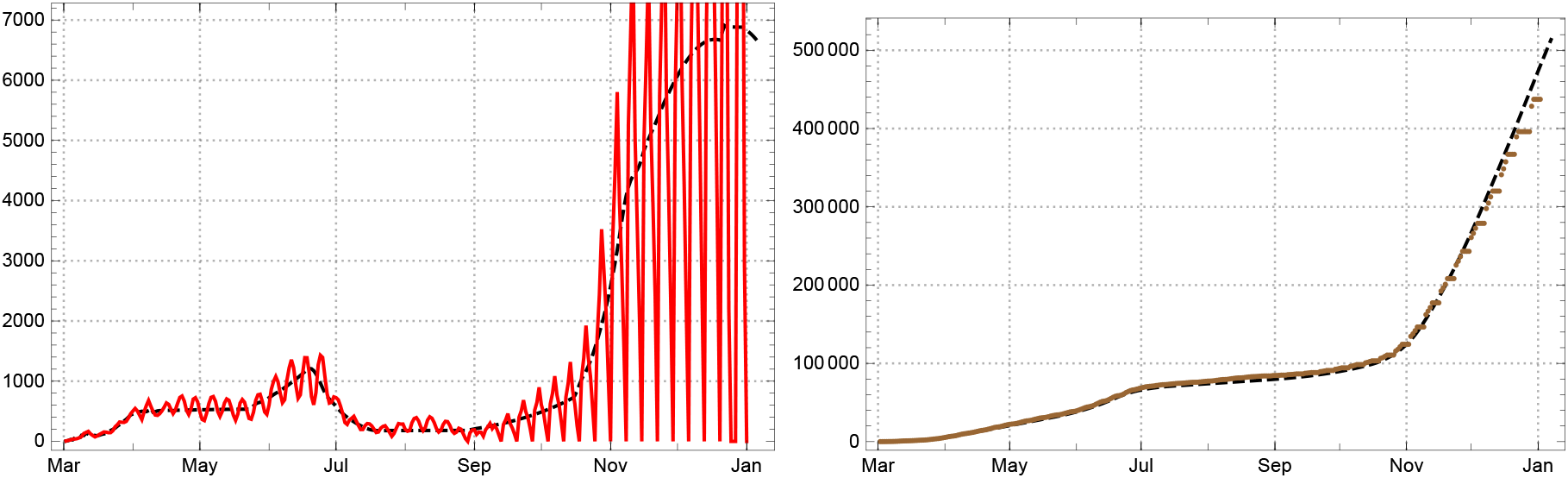
Left: Daily new reported infected for the Sweden, empirical 3-day averages Â_*new*,3_ solid red; model *A*_*new*_ black dashed. Right: Total number of reported infected (brown); empirical Â_*tot*_ solid, model *A*_*tot*_ dashed.

The JHU statistics does not register recovered people for Sweden at all; only deaths are reported. In consequence the usual interpretation of (9) as characterizing the “actual” infected breaks down for Sweden^8^ and the estimation (11) for the mean duration of illness becomes meaningless (fig. 35, left). An indication of the extent of reported actual diseased is given by the *q*-corrected number *Â*_*q*_ (same figure, right).

**Figure 35.**
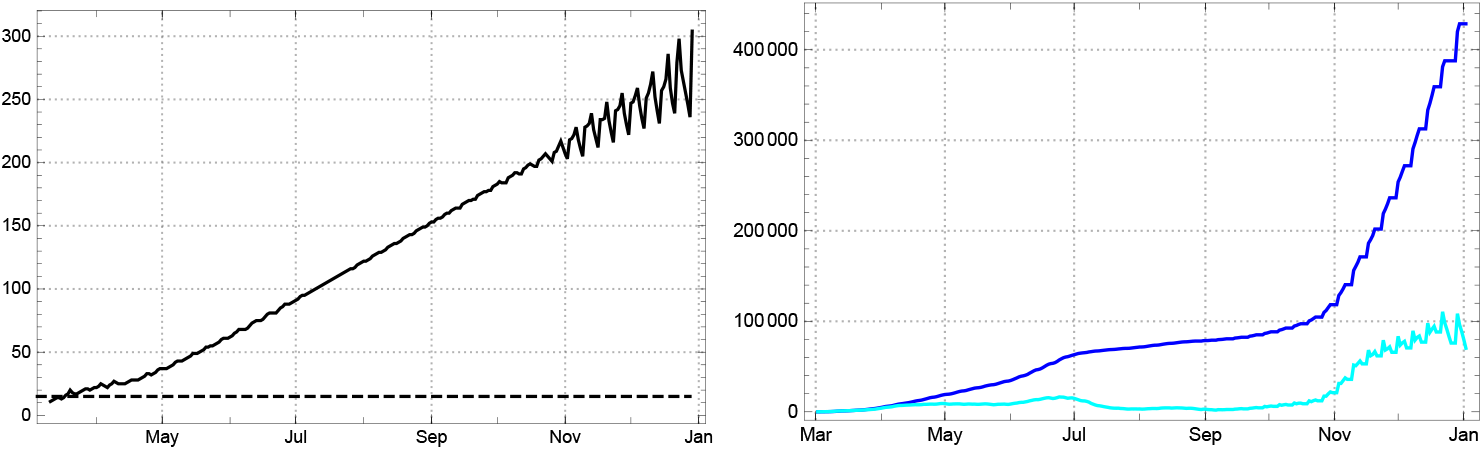
Left: Daily values of the mean time of statistically actual infection 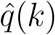 for Sweden. Right: Comparison of reported infected *Â*(dark blue) and *q*-corrected number (*q* = 15) of recorded actual infected *Â*_*q*_ (bright blue) from the JHU data in Sweden.

In this sense, the synopsis with a collection of the “3 curves” can be given for Sweden like for any other country (fig. 37). Of course the model reproduces the empirical values Â(*k*) even in such an extreme case if the time dependent empirical values of (11) are used for the the model calculation, 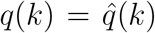, while it reconstructs the *q*-corrected numbers for the estimate of actually infected *Â*_*q*_ (*k*) if the respective constant is used, here *q* = 15 (fig. 36).

**Figure 36.**
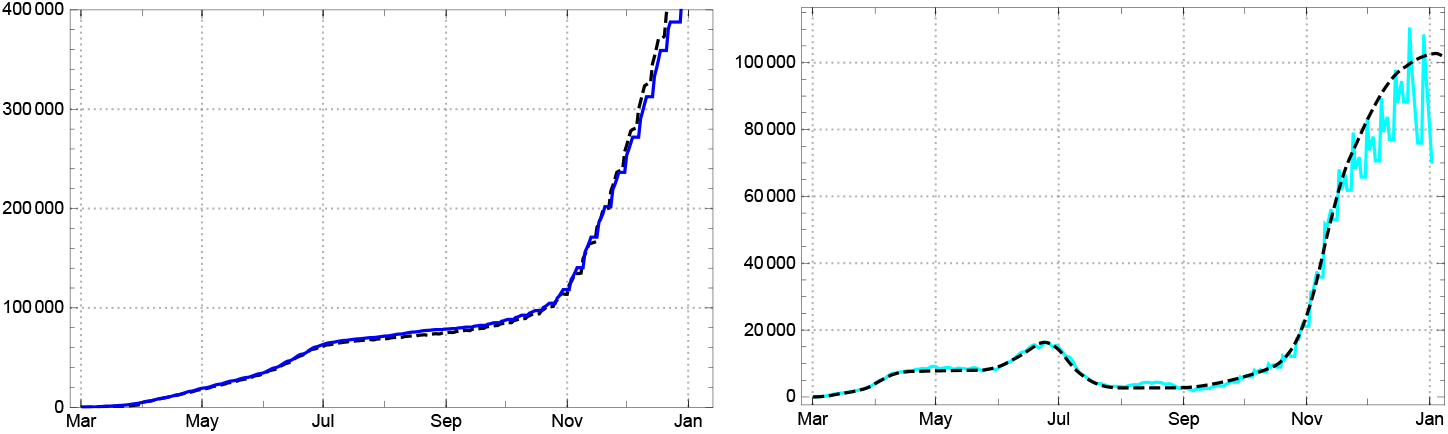
Left: Empirical data *Â*(*k*) for Sweden (blue) and model values *A*(*k*) determined with time varying 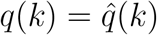 (black dashed). Right: Empirical values, *q*-corrected, for statistically actual cases *Â*_*q*_ (*k*) and the corresponding model values *A*_*q*_(*k*) (black dashed).

**Figure 37.**
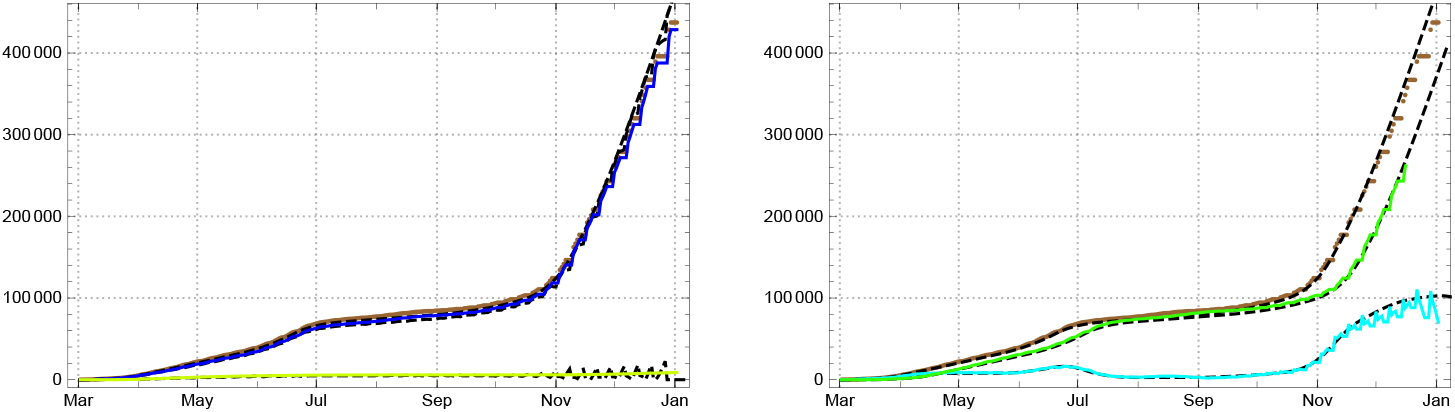
Left: Numbers of totally infected *Â*_*tot*_ (brown), reported redrawn 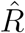 (bright green) – here deaths only – and the difference *Â* (blue) for Sweden; model values black dashed. Right: the same for *Â*_*tot*_ (brown), *q*-corrected *Â*_*q*_ (bright blue) and the redrawn 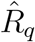 (green) as the difference (model values black dashed).

### 3.2. The three most stricken regions: USA, Brazil, India

In this section we give a short analysis of the course of the pandemic during 2020 for the three countries which have to bemoan the largest numbers of deceased and huge numbers of infected (USA, Brazil, India). We expected higher dark factors *δ* than for the European countries discussed above and checked this expectation by the same heuristic approach as used for Sweden, i.e. by a comparative judgement of the changes of the empirically determined strength of infection 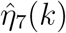, which result from different assumptions of the values for *δ*. To our surprise we found no clear evidence for an overall larger dark factor for the USA than for the European countries and work here with *δ* = 4, while for India there are strong indications of a large dark factor which we estimate as *δ* ≈35 (see below). For Brazil we consider *δ* ≈8 a reasonable choice for the overall development of the epidemic. Don’t forget, however, that all these are plausibility considerations which are not based on representative serological studies.

*USA*. At the beginning of the pandemics the United States of America (population 333 M) suffered a rapid rise of infections with an initial reproduction rate well above 5. In early April 2020 this dynamics was broken and a slow decrease started for about 2 months. In mid June a second wave with an upswing for about a month and a reproduction rate shortly below 1.3 followed. In late August and early September the subsequent downswing faded out. After a short phase of indecision the beginnings of a third wave became clearly visible; it lasted until (at least) mid December. Figure 38 shows the daily new infections, the reproduction rate 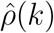 determined from the JHU data and the strength of infection 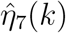 assuming *δ* = 4.

**Figure 38.**
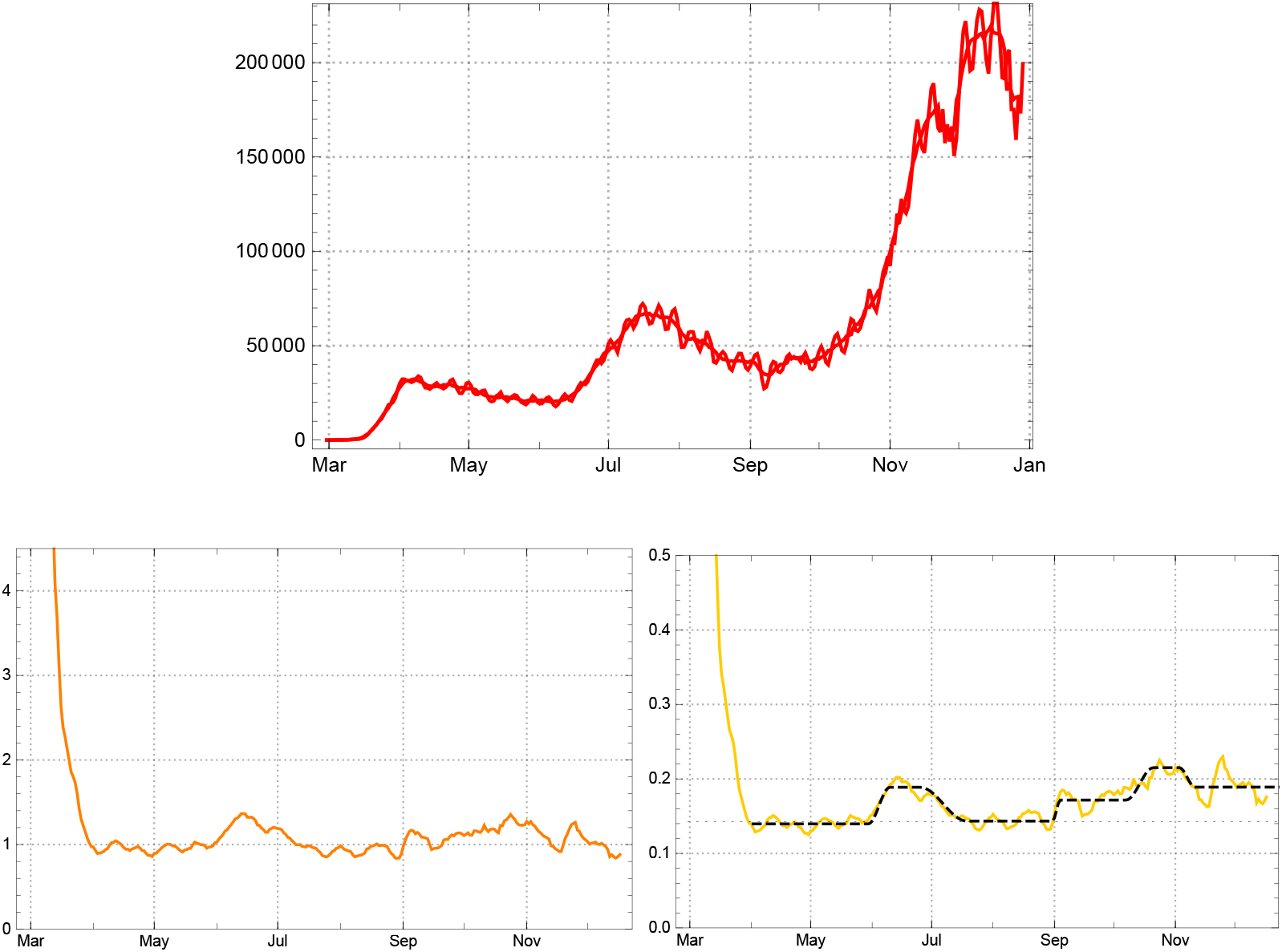
Top: 3-day and 7-day sliding averages of daily new reported cases, *Â*_*new*,3_(*k*), *Â*_*new*,7_(*k*), for the USA. Bottom, left: Empirical reproduction rates 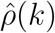 for the USA. Right: Daily strength of infection 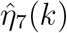 for the USA (yellow), with *δ* = 4; model parameters *η*_*j*_ in the main intervals *J*_*j*_ black dashed.

Figure 39 displays three variants of 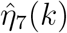 for *δ* = 0, 4, 8. The third one shows an implausible increase for the strength of infection at the end of the year, which would seem reasonable only if one of the new, more aggressive mutants of the virus had started to spread in the USA in September 2020 already. Without further evidence we do not assume such a strong case. As *δ* = 0 contradicts all evidences collected on unreported infected, we choose *δ* = 4 for the SEPAR_*d*_ model of the USA. Also here we find a moderate increase of the mean level of the 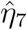. To judge whether this may be due to the inconsiderate behaviour of part of the US population (supporters of the outgoing president) or the first influences of a virus mutation and/or still other factors is beyond the scope if this paper and our competence.

**Figure 39.**
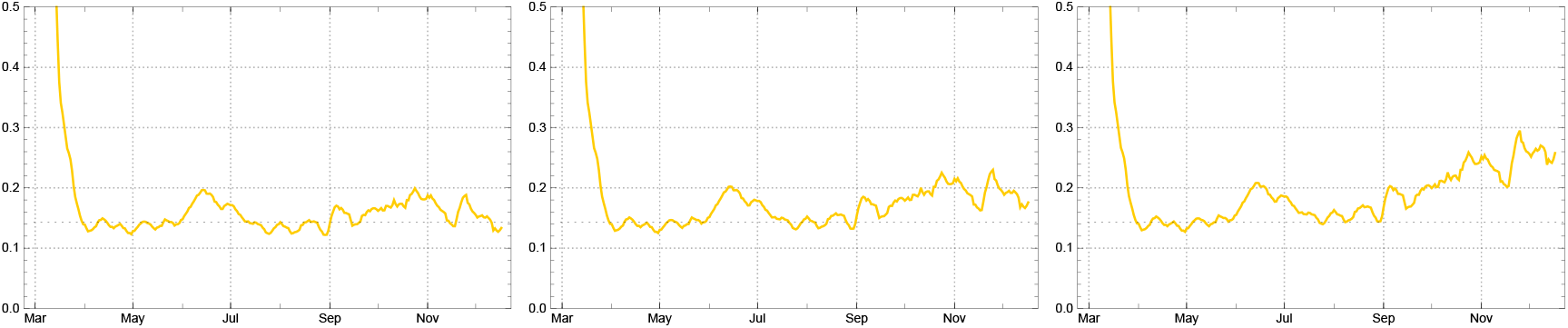
Daily strength of infection (sliding 7-day averages) 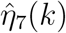 for USA, assuming different dark factors: *δ* = 0 (left), *δ* = 4 (middle), and *δ* = 8 (right).

In section 2 it was already noted that the estimation of the time of sojourn in the “actual” state of infectivity, suggested by the statistics for the USA, leads to surprising effects. It rises from about 15 in March 2020 to above 100 in early November, with a moderate platform in between; then it starts falling,before it makes an abrupt jump (fig 40, left). The jump of 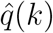 is an artefact of a change in the record keeping: from December 14, 2020 onward the reporting of data of recovered people was given up (*Rec*(*k*) = 0 for date of *k* after 2020/12/14). Of course this jump is also reflected in the numbers of recorded actual infected *Â*(*k*) (same figure, right).

**Figure 40.**
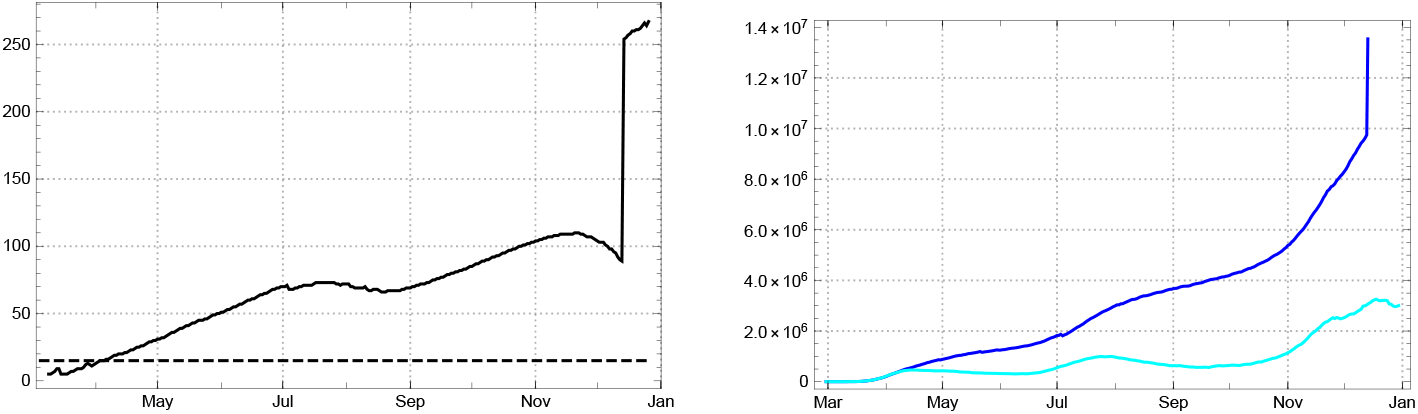
Left: Daily values of the mean time of statistically actual (“active”) infection 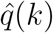 for USA. Right: Empirical data (JHU) for statistically actual cases *Â*(*k*) (dark blue) versus *q*-corrected ones, *q* = 15, *Â*_*q*_ (*k*) (bright blue) for the USA.

The main (constancy) intervals of the model are visible in the graph of the daily strength of infection 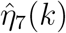 in fig. 38, bottom right. The dates of the time marker between the intervals are: *t*_0_= 02/29 2020; *t*_1_ = 04/02, *t*_2_ = 06/10, *t*_3_ = 07/19, *t*_4_ = 09/04, *t*_5_ = 10/21 *t*_6_ = 11/11, end of data *t*_*eod*_ = 12/30. Expressed in terms of the country day count with *k*_0_ = 1 (∼39 in the JHU day count) the main intervals for the USA are: *J*_0_ = [1, 33], *J*_1_ = [34, 92], *J*_2_ = [103, 118], *J*_3_ = [142, 185], *J*_4_ = [189, 222], *J*_5_ = [236, 249], *J*_6_ = [257, 322].

The start parameter for the strength of infection *η*_0_ and a slightly adapted choice of parameter values *η*_*j*_ inside the 1-sigma domain of the respective interval *J*_*j*_ are given by the following table.

| Model $\eta_0$ and $\eta_j$, $\rho_j$ in intervals $J_j$ for the USA | | | | | | | |
| --- | --- | --- | --- | --- | --- | --- | --- |
| | $\eta_0$ | $J_1$ | $J_2$ | $J_3$ | $J_4$ | $J_5$ | $J_6$ |
| $\eta_j$ | -1.011 | 0.140 | 0.189 | 0.143 | 0.172 | 0.215 | 0.194 |
| $\rho_j$ | — | 0.97 | 1.28 | 0.94 | 1.08 | 1.30 | 1.12 |

Here, as for the other countries, the *ρ*_*j*_ denote the model reproduction rates at the beginning of the respective intervals. Assuming the dark factor *δ* = 4, the effective reproduction rate *ρ*(*k*) changes considerably from the beginning of *J*_6_ until the end of the year, *s*(11*/*11 2020) = 0.82 to *s*(12*/*31) = 0.67, (cf.fig. 41).

**Figure 41.**
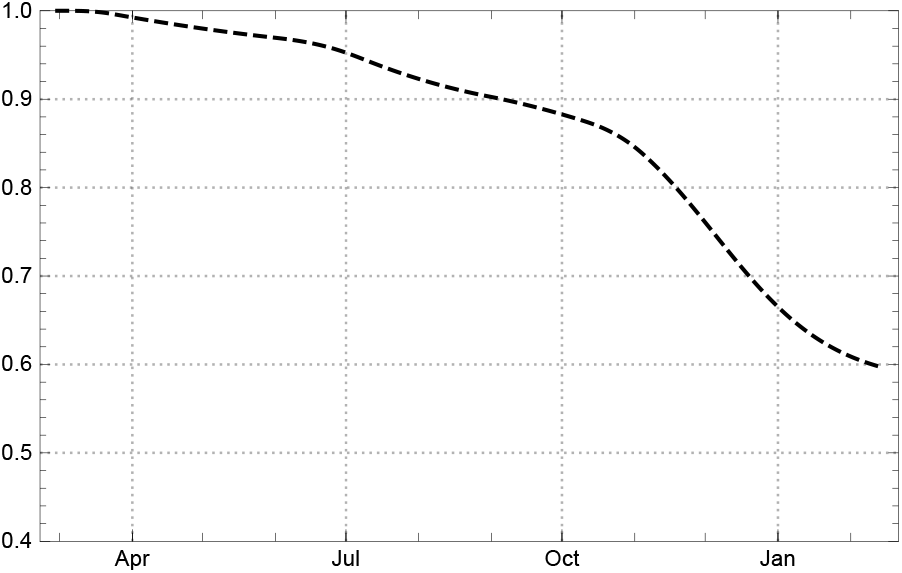
Ratio of susceptibles *s*(*k*) (model values) for the USA, *δ* = 4.

In consequence, the reproduction rate at the end of the year is down to *ρ*(12*/*31) = 0.86; and the newly reported infected are expected to reach a peak in late December (fig. 42, right). Apparently this does not agree with the data, while the total number of infections is well reproduced by the SEPAR_*d*_ model (same figure, left). The difference for the new infected may be an *indication that our estimate of the dark factor δ is wrong or the strength of infection at the beginning of the new year 2021 is drastically boosted*, e.g., by a rapid spread of a new, more aggressive mutant of the virus.

**Figure 42.**
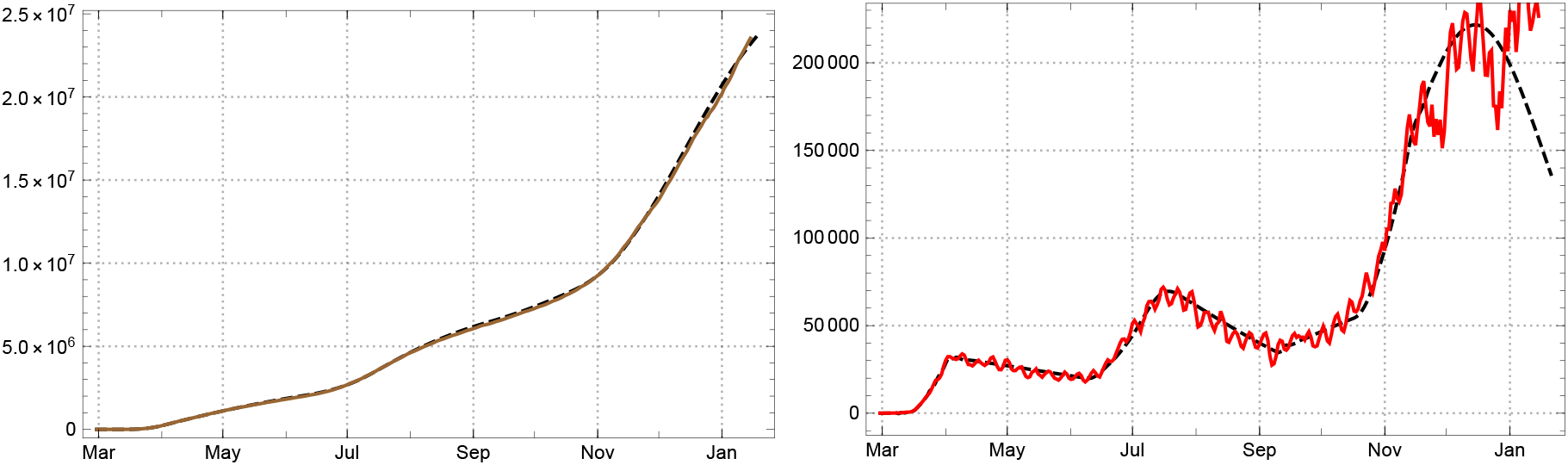
Left: Total number of infected (brown); empirical *Â*_*tot*_ solid, model *A*_*tot*_ dashed. Right: Daily newly reported for the USA, 7-day averages (red); empirical *Â*_*new*_ solid red, model _*new*_ black dashed (*δ* = 4).

As noted above, the recovered people are documented with increasingly large time delays in the records for the USA. Therefore it seems preferable to compare the model value of actually infected, *A*(*k*), with *Â*_*q*_(*k*) rather than with *Â*(*k*) (fig. 43). The empirical determined values of *Â*_*q*_ (*k*) are shown in bright blue in the figure. They are marked by three local maxima indicating the peak values of three waves of the epidemics in the USA. These peaks are blurred in the graph of *Â*because too many of the effectively redrawn are dragged along as acute cases in the statistics. Due to under-reporting at the end of the year, the local extremum in December may be fuzzier than it appears here. But keep in mind that the SEPAR_*d*_ model with *δ* = 4 *predicts a local maximum* inside the interval *J*_6_, if the contact behaviour and the resulting strength of infectivity *η*_6_ do not change considerably.

**Figure 43.**
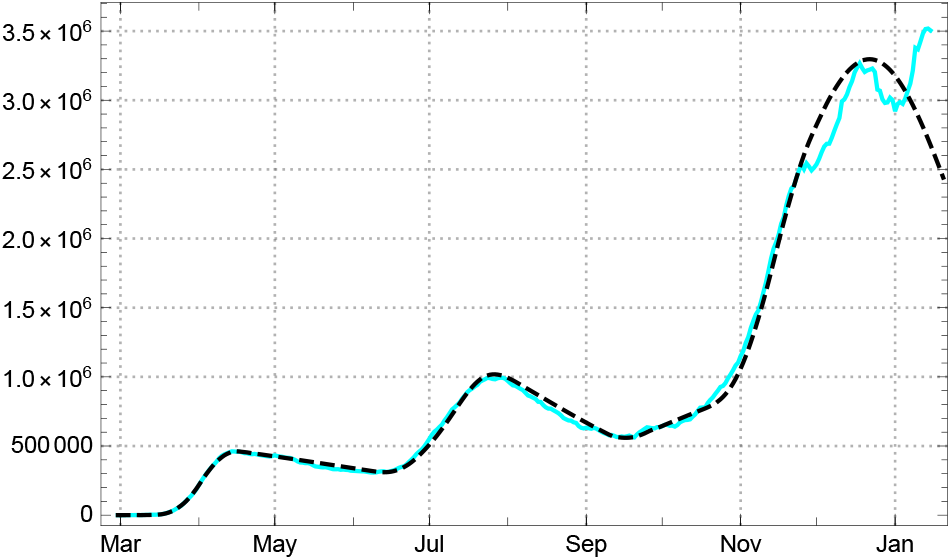
Number of *q*-corrected actual infected for the USA; empirical *Â*_*q*_ (bright blue) and model values *A*_*q*_ (black dashed).

Figure 44, left, shows the 3 curves for the model values (black dashed) of the total number of infected *A*_*tot*_, the actually infected *A* in terms of estimates with constant *q* = 15, and the redrawn *R*, all of them compared with the corresponding values 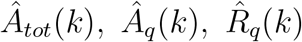 determined from the JHU data (coloured solid lines). By using time dependent values *q*(*k*), like, e.g., in the case of Germany, SEPAR_*d*_ is able to model the statistically “actual” cases also here (fig. 44, right). Because of the growing fictitiousness of the numbers *Â*(*k*) in the case of the USA we prefer, however, to look at the corrected values *Â*_*q*_ (*k*), as stated already.

**Figure 44.**
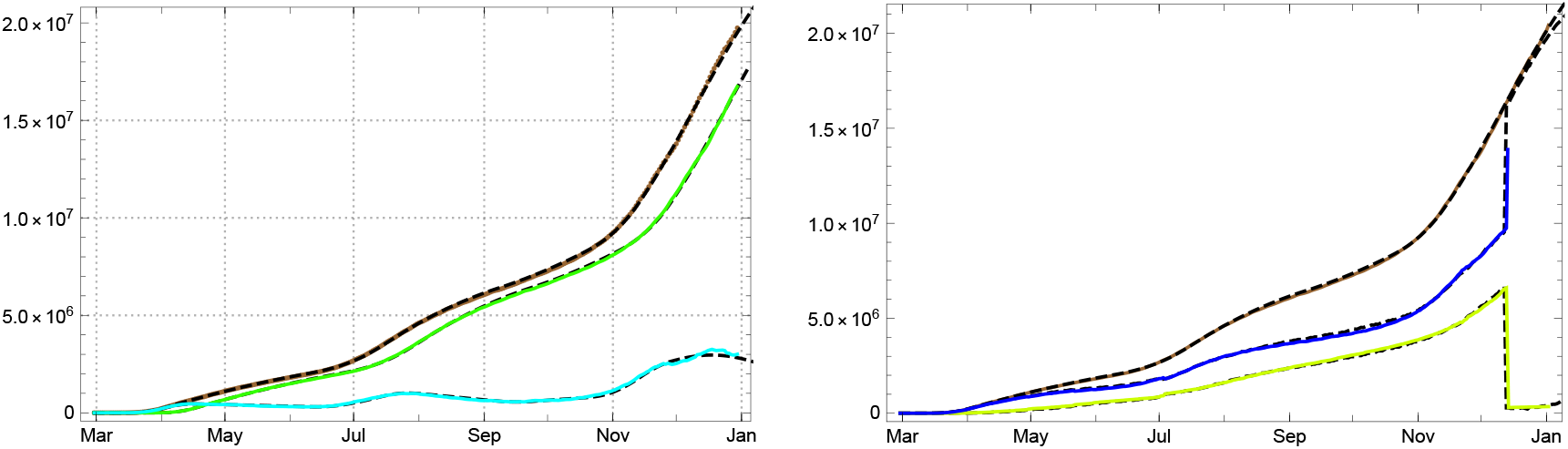
Left: numbers of totally infected *Â*_*tot*_ (brown), redrawn 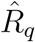 (green), and *q*-corrected actual numbers *Â*_*q*_ (bright blue). Right: Numbers of totally infected *Â*_*tot*_(brown), reported redrawn 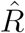 (bright green), and actual numbers *Â*of the statistic (blue) for the USA. Empirical data (solid coloured lines) and model values (black dashed).

#### Brazil

The documentation of newly reported became non-sporadic in Brazil (population 212 M) at *t*_0_ = March 15, 2020. A first peak for the officially recorded number of actual infected *Â*(*k*) was surpassed in early August 2020 with a decreasing phase until late October, after which a second wave started (fig. 45 left). In contrast to the USA we find here a comparatively stable estimate for the mean time 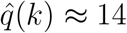 (fig. 45 right).

**Figure 45.**
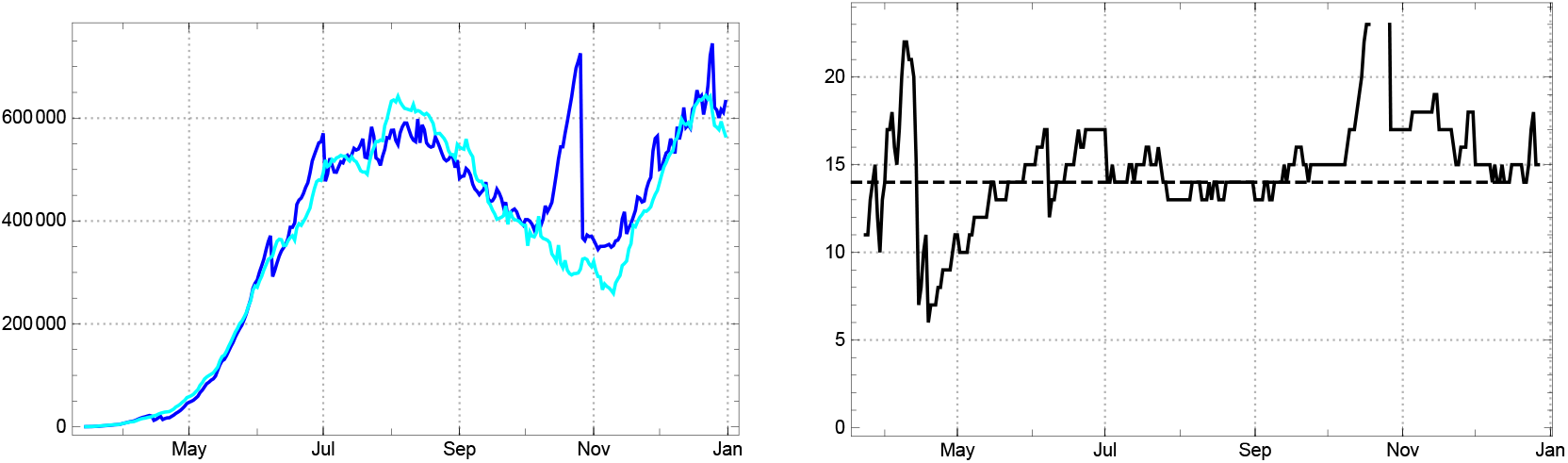
Left: Acute infected *Â*(*k*) recorded by the statistics (dark blue) in comparison with *q*-equalized number *Â*_*q*_ (*k*) (bright blue) for Brazil (*q* = 14). Right: Empricial estimate 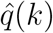 of mean duration of active infective according to the statistics for Brazil.

The outlier peak of *Â*(*k*) about October 25 appears also as an exceptional peak in the 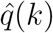. Apparently it is due to an interruption of writing-off actual infected to the redrawn (compare fig. 51). Up to this exceptional phase there is a close incidence between the *Â*(*k*) and the *q*-corrected number *Â*_*q*_ (*k*). The minimum of the mean square difference is acquired for *q* = 14. The numbers of newly reported *Â*_*new*_ (*k*) show strong daily fluctuations which are smoothed by the 7-day sliding average *Â*_*new*,7_(*k*) (fig. 46).

**Figure 46.**
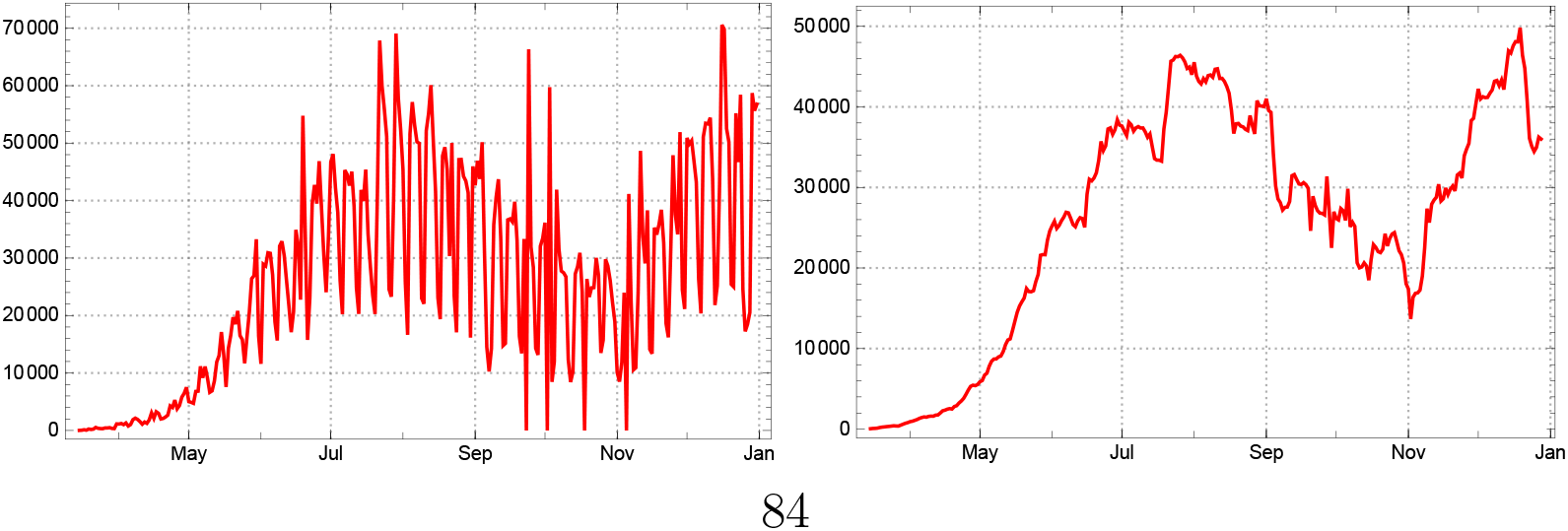
Daily varying numbers *Â*_*new*_ (*k*) (left)versus 7-day sliding averages *Â*_*new*,7_(*k*) (right) of new infections for Brazil.

**Figure 47.**
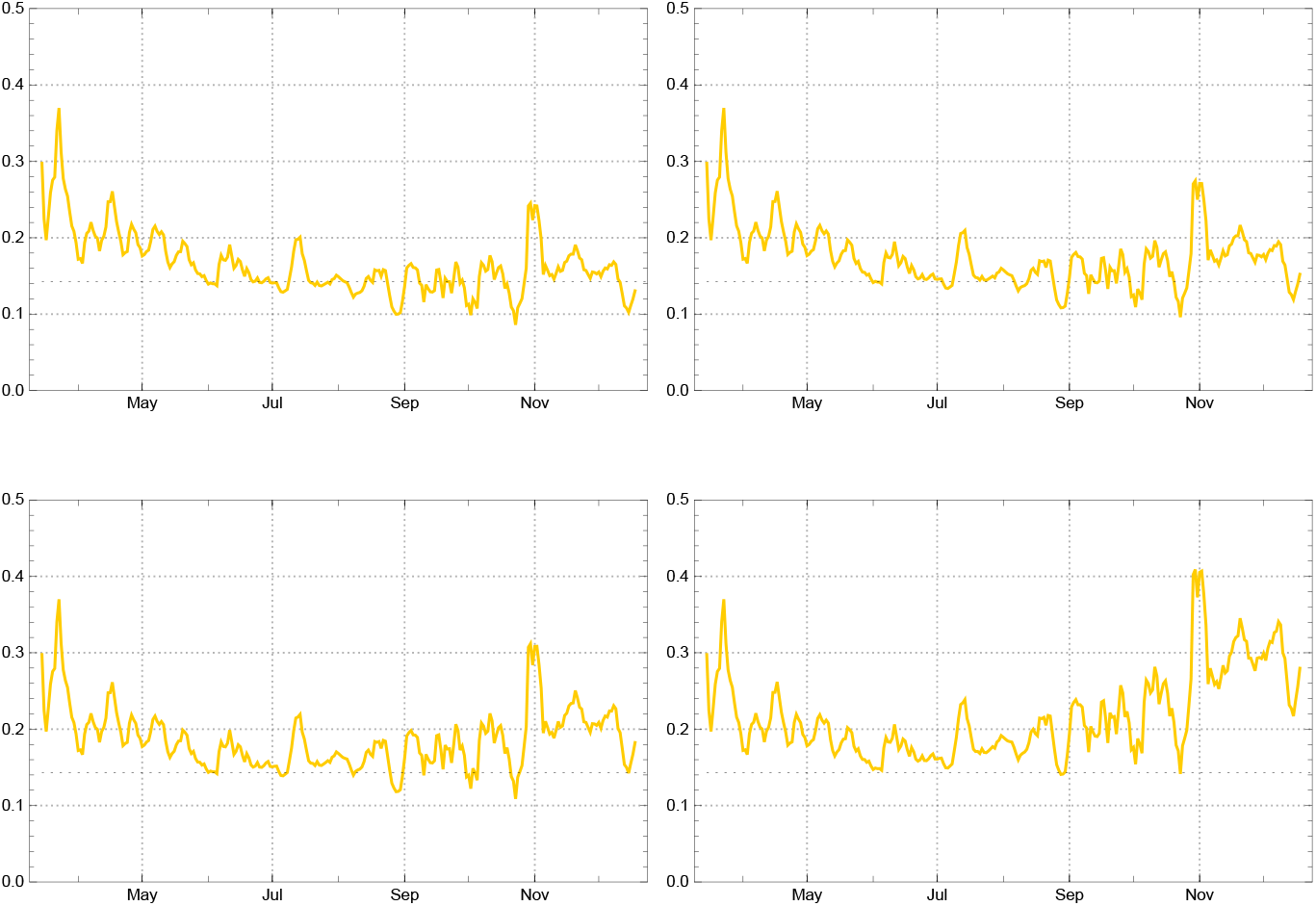
Daily strength of infection (sliding 7-day averages) 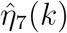 for Brazil, assuming different dark factors. Top: *δ* = 0 (left) and *δ* = 4 (right). Bottom: *δ* = 8 (left) and *δ* = 15 (right).

**Figure 48.**
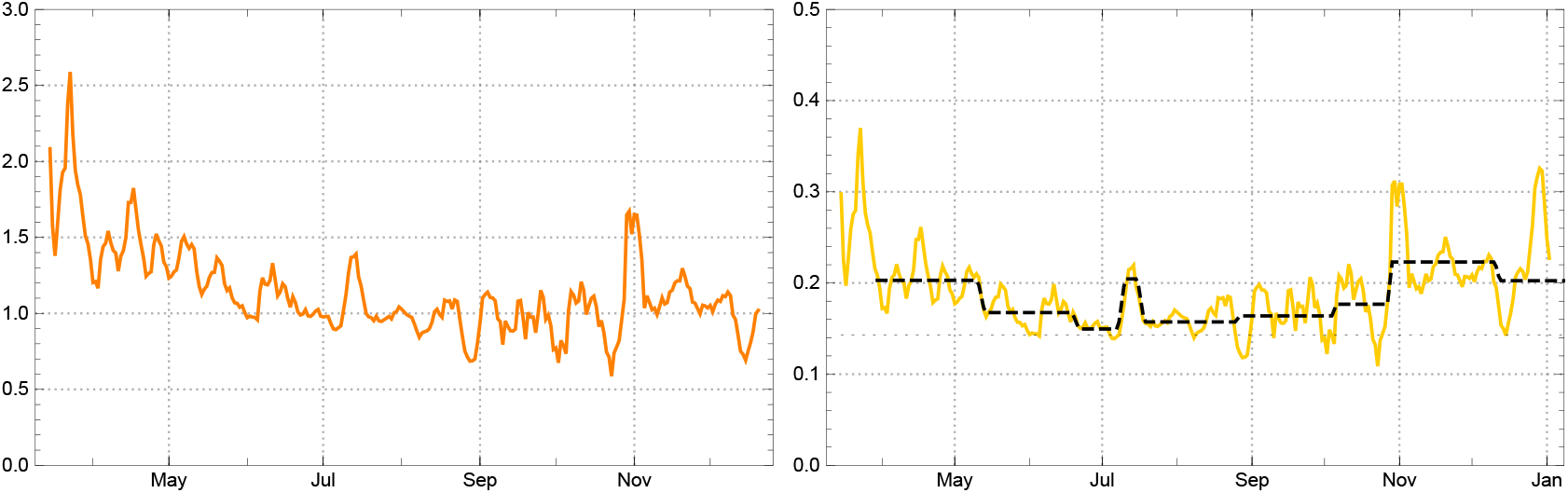
Left: Empirical reproduction rates 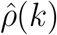 for Brazil (orange). Right: Daily strength of infection, empirical 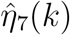 (yellow), for Brazil assuming *δ* = 8, and model parameters *η*_*j*_ in the main intervals *J*_*j*_ (black dashed).

**Figure 49.**
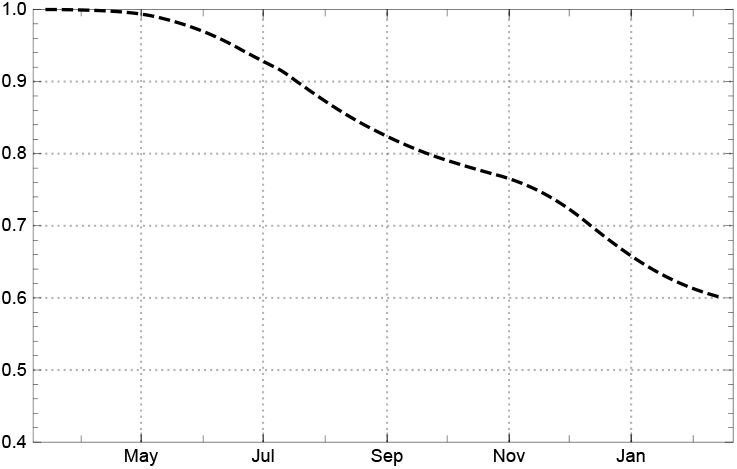
Model values of the ratio of susceptibles *s*(*k*) for Brazil (*δ* = 8).

**Figure 50.**
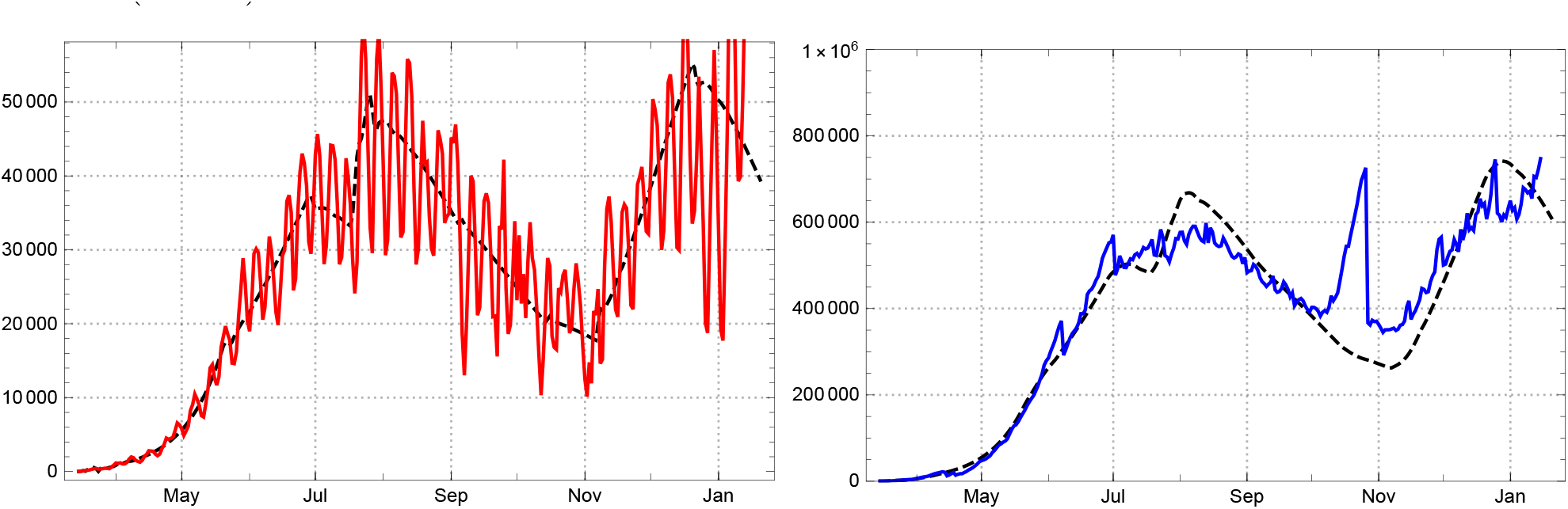
Left: Empirical values (3-day average) for daily newly reported for Brazil *Â*_*new*,3_ (solid red line)) and model values black dashed). Right: Actual cases *Â*(blue), model values (black dashed).

**Figure 51.**
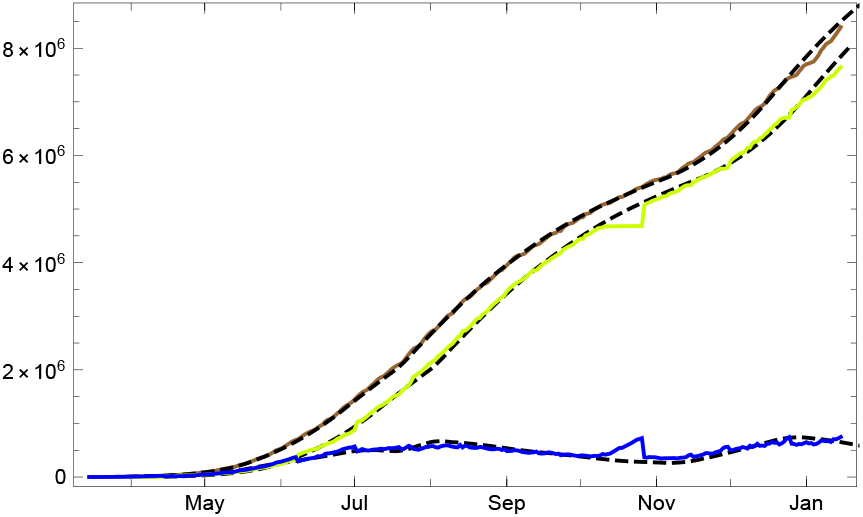
Empirical data (coloured solid lines) for the numbers of totally infected *Â*_*tot*_(brown), redrawn 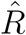 (bright green), actual infected *Â*(blue) and the respective model values (black dashed) for Brazil (*δ* = 8).

A comparison of different strengths of infection 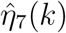 indicates a value between *δ* = 0 and *δ* = 8 as a plausible choice (fig. 47). For higher values an unnatural increase of the infection strength would appear close to the end of the year (if not due to a new mutant. As we assume that in Brazil the dark factor is higher than in European countries, we use *δ* = 8 as a plausible model hypothesis.

The daily reproduction numbers 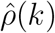 (independent of *δ*) start from a lower level than for many other countries, slightly above 2. They show a relatively stable downward trend, falling below 1 for a few days in early June and for longer periods after June 22, 2020 (fig. 48, left). But already at the end of March, when the reproduction rate was still considerably above 1 (*t*_0_ = March 30), its downward trend was already slow enough to allow for approximation by constancy intervals.

In the case of Brazil the initial interval starts at *t*_0_ = 03/15, 2020. The following time can be subdivided into main intervals *J*_*j*_ in which the averaged daily strength of infection 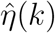 can be replaced by their mean values *η*_*j*_, starting with *t*_1_ = 03/30. The main intervals are separated by the days *t*_2_ = 05/14, *t*_3_ = 06/22, *t*_4_ = 07/11, *t*_5_ = 07/19 *t*_6_ = 08/27, *t*_7_ = 08/10, *t*_8_ = 10/29, *t*_9_ = 12/13, 2020.

They can well be discerned in fig. 48, right, showing the daily strength of infection 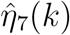 (yellow) and their mean values (black dashed) in these intervals. In the country day count *k*_0_ = 1 (∼70 in the JHU count) the main intervals are *J*_0_ = [1, 15], *J*_1_ = [16, 59], *J*_2_ = [61, 97], *J*_3_ = [100, 115], *J*_4_ = [119, 122], *J*_5_ = [127, 164], *J*_6_ = [166, 204], *J*_7_ = [206, 220], *J*_8_ = [222, 271], *J*_9_ = [274, *t*_*eod*_], here with the end of data *t*_*eod*_ = 292.

The start parameter *η*_0_ and the model reproduction numbers *ρ*_*j*_ in the respective interval *J*_*j*_ are

| Model $\eta_0$ and $\eta_j$ and $\rho_j$ in $J_j$ for Brazil | | | | | | | | | | |
| --- | --- | --- | --- | --- | --- | --- | --- | --- | --- | --- |
| | $\eta_0$ | $J_1$ | $J_2$ | $J_3$ | $J_4$ | $J_5$ | $J_6$ | $J_6$ | $J_8$ | $J_9$ |
| $\eta_j$ | 0.031 | 0.202 | 0.164 | 0.142 | 0.191 | 0.143 | 0.141 | 0.148 | 0.180 | 0.172 |
| $\rho_j$ | — | 1.42 | 1.14 | 0.98 | 1.30 | 0.96 | 0.94 | 0.97 | 1.17 | 1.00 |

Also here the *ρ*_*j*_ designate reproduction numbers at the beginning of the *j*-the interval and the fall of *s*(*k*) makes the reproduction number cross the critical value in the last main interval (cf. 49). This does not mean that it will stay there.

The resulting model curves and their relationship to the empirical data for new infections and acual infections are shown in fig. 50. A panel of the *three curves A*_*tot*_, *A, R* is shown in fig. 51. Here one sees clearly that the outlier bump of *Â*(*k*) is accompanied by an inverse outlier in 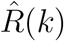.

#### India

The recorded data on *Â*_*new*_ (*k*) for India (population 1387 M) start to be non-sporadic at *t*_0_ = March 4, 2020. From this time on we find a steady growth of the number of reported actual infected *Â*(*k*) until early September. Because the size of the country and the life conditions in large parts of it a comparatively high number of unrecorded infected may be assumed, with a dark factor at the order of magnitude *δ* ∼10 probably 20 ≤ *δ* ≤ 50.^9^ In early September the tide changed and a nearly monotonous decline of actual infected started. With the exception of a short intermediate dodge the decline continues at the end of 2020 (fig. 52, left). Although in late December 2020 there were only about 0.7 % recorded infected in India, the high quota of unreported infected poses the question whether the downturn in late summer may already be due to a the decrease of the fraction of susceptibles *s*(*k*).

**Figure 52.**
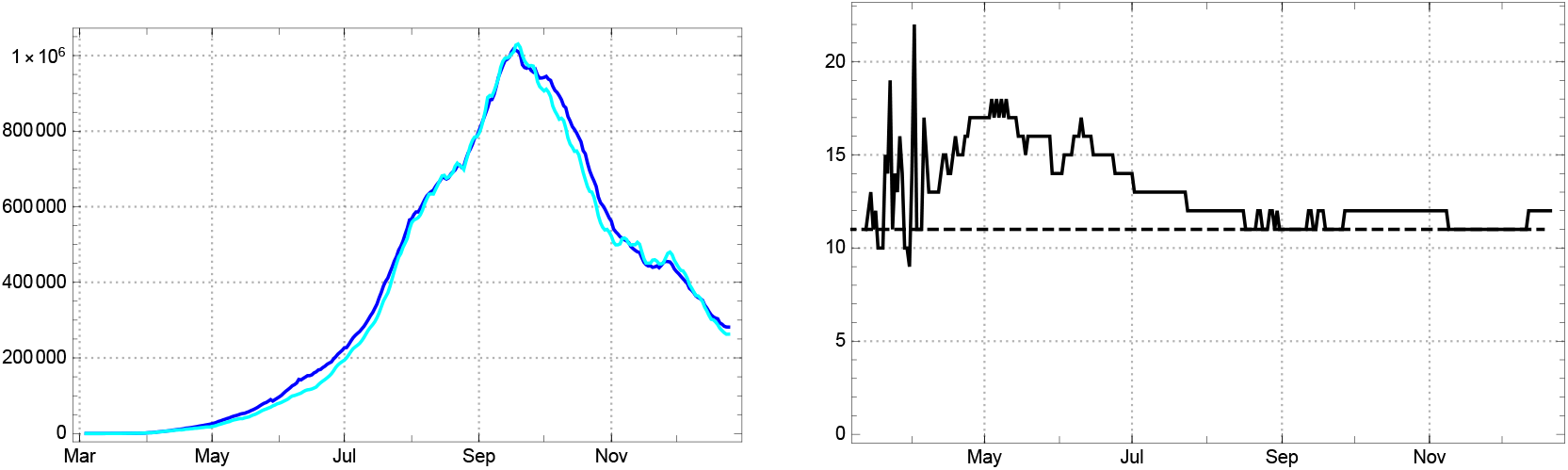
Left: Actual infected *Â*(*k*) recorded by the statistics (dark blue) in comparison with *q*-equalized number *Â*_*q*_ (*k*) (bright blue) for India (*q* = 11). Right: Empiricial estimate 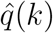 of mean duration of actual infectived according to the statistics, i.e. in *Â*, for India.

Before we discuss this point let us remark that the time of being statistically recorded as actual case is relatively stable in the Indian data, with a good approximative constant value *q* ≈11 (fig. 52, right). In consequence *Â*(*k*) does not differ much from *Â*_*q*_ (*k*) (same fig. left). This allows to use the recorded data *Â*in the following without the proviso to be made in the case of the USA.

The reproduction numbers and the corresponding daily strength of infection of the model are derived from the 7-day sliding averages of newly reported. Figure 53 shows both the daily varying *Â*_*new*_ and the averaged *Â*_*new*,7_.

**Figure 53.**
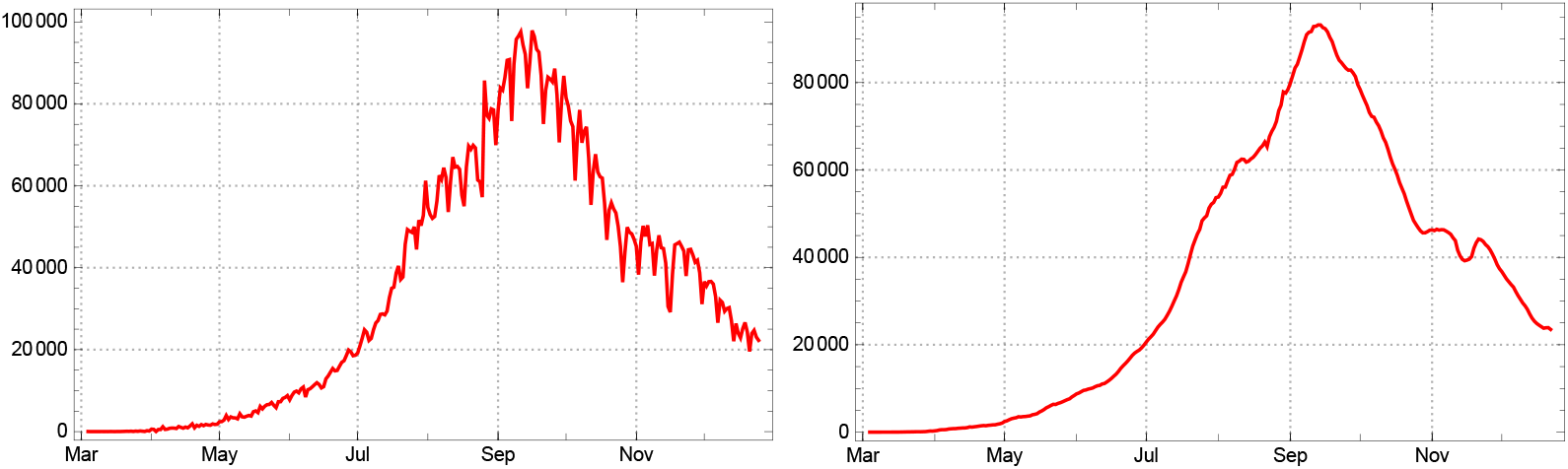
Top: Empirical number of daily new infections *Â*_*new*_ and 7-day sliding average *Â*_*new*,7_ for India.

The reproduction number fell rapidly from roughly 3.5 at the beginning to below 1.5 in early April, and 1.2 in late May, after which it continued to decrease with minor fluctuations. In early September it dropped below the critical value 1, where it stayed with few exceptional fluctuations until December (fig. 54). It runs, of course, parallel to the daily strength of infection 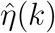 calculated from the 7-day averages if abstraction is made from the dark sector, *δ* = 0 (fig. 55, left). Here we confront it with the more realistic graph of 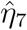 calculated under the assumption of a dark sector.

**Figure 54.**
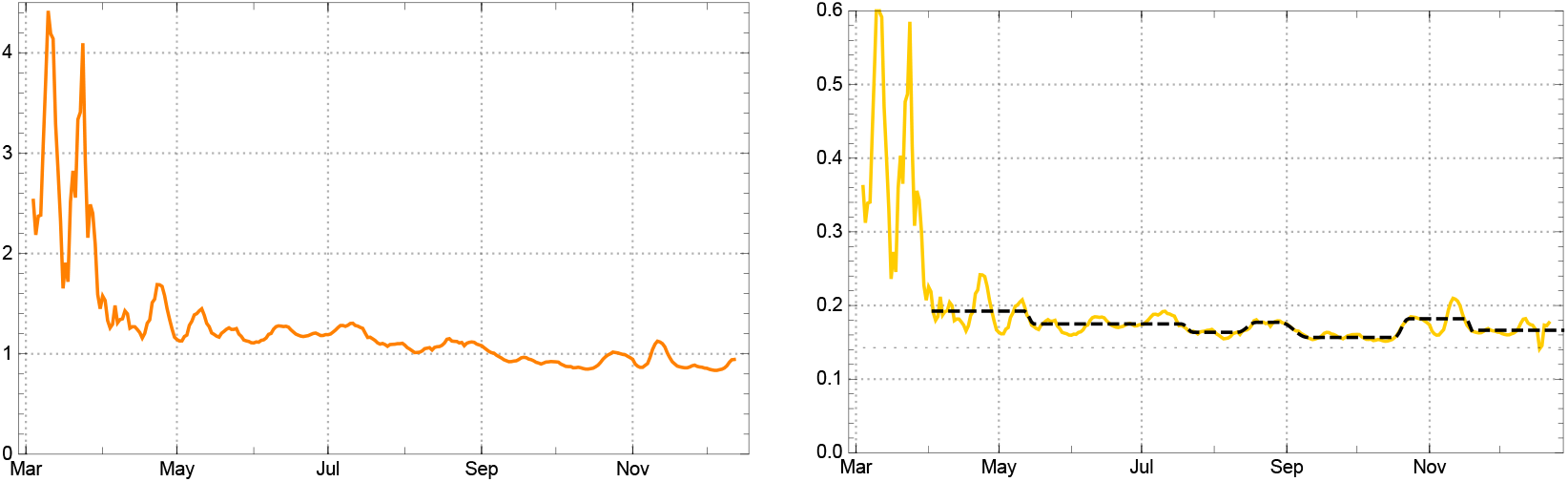
Left: Empirically determined reproduction rates 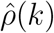 for India (iorange). Right: Empirical infections strength 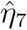 (yellow) assuming a dark sector with *δ* = 35; model values for *η* black dotted.

**Figure 55.**
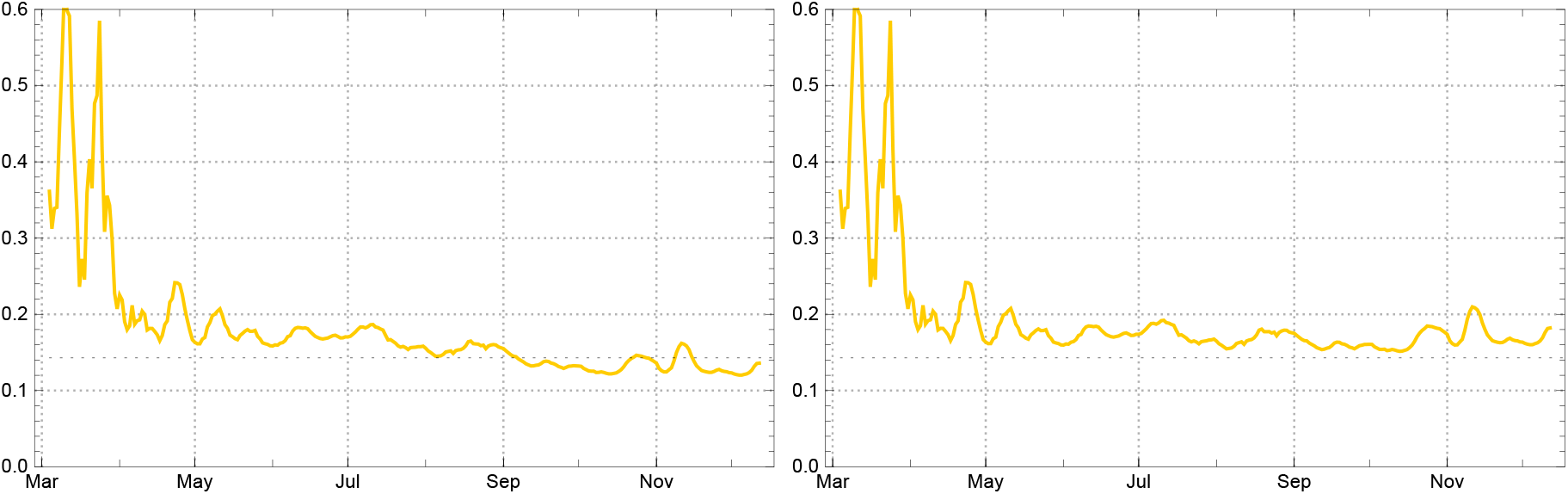
Left: Empirical daily strength of infection 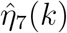 for India (yellow) with no dark sector, i.e. assuming *δ* = 0. Right: Empirical strength of infection 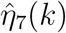 for India (yellow), assuming a dark sector with factor *δ* = 35

Such an idealized scenario with *δ* = 0 is shown in fig 55, left. If, on the other hand, the empirical daily strength of infection are determined under the more realistic assumption of a non-negligible dark sector, e.g. *δ* = 35, the picture is different (same figure, right). Here one finds a daily strength of infection moderately fluctuating in a narrow band between 10 and 20 % above the critical value 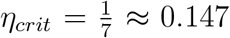, rather than dropping below it in early September like in the first case. We do not know of any indications for a changing contact behaviour of the population in India; neither can we assume a decreasing aggressiveness of the virus. Therefore the first scenario (*δ* ≈0) looks highly unrealistic. In both cases the empirically determined reproduction rates 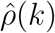 are the same (fig. 54). In the second case the fall of 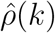 below 1 in early September is due to the lowering of *ŝ* (*k*), i.e. as an effect of an incipient herd immunization.

But how can that be with a herd immunization quota of 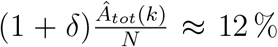, even with *δ* = 35, in September 2020?^10^ The reason lies in the comparatively low overall daily strength of infection. Between May and December 2020 it fluctuated between 10 and 20 % above the critical level (fig. 55, right). Even if part of the low level had to be ascribed to an intentional under-reporting of the *Â*_*new*_ (*k*), this would mean an increasing size of the dark sector; the overall effect would be the same.^11^ For modelling the epidemic in India we can do with 7 constancy intervals. After the initial interval *J*_0_ = [1, 30] in the day count of the country (*k*_0_ ∼43 in the JHU count) the main intervals are *J*_1_ = [31, 70], *J*_2_ = [74, 137], *J*_3_ = [142, 161], *J*_4_ = [169, 179], *J*_5_ = [191, 228], *J*_6_ = [234, 256], *J*_7_ = [260, *k*_*eod*_], with end of data *k*_*eod*_ = 297. The date of the interval separators are *t*_0_ = 03/04, *t*_1_ = 04/03, *t*_2_ = 05/16, *t*_3_ = 07/23, *t*_4_ = 08/19, *t*_5_ = 09/10, *t*_6_ = 10/23, *t*_7_ = 11/18, end of data *t*_*eod*_ = 12/26 2020.

Similar to the Brazilian case, the reproduction rate surpasses 1 in the first four main intervals; only in mid September a downswing of the epidemic started, interrupted by an intermediate dodge at the beginning of November. In mid September the total number of acknowledged infected was roughly *A*_*tot*_(240) ≈5 *M*, about 3.8 per mill of the total population; but with a dark quota of *δ* = 35 the total number of infected had probably already risen above the 10 % margin (see above).

The start parameter of the model *η*_0_ is chosen according to the best adaptation to the 7-day averaged data (without a claim for a directly realistic interpretation) and the parameter values *η*_*j*_ essentially as the mean values of the 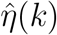 in the respective interval *J*_*j*_. They are given in the table.

| Model $\eta_0$ and $\eta_j, \rho_j$ in $J_j$ for India | | | | | | | | |
| --- | --- | --- | --- | --- | --- | --- | --- | --- |
| | $\eta_0$ | $J_1$ | $J_2$ | $J_3$ | $J_4$ | $J_5$ | $J_6$ | $J_7$ |
| $\eta_j$ | -0.138 | 0.192 | 0.175 | 0.164 | 0.177 | 0.157 | 0.182 | 0.166 |
| $\rho_j$ | — | 1.34 | 1.2 | 1.07 | 1.12 | 0.90 | 0.99 | 0.87 |

With these parameters the SEPAR model leads to a convincing reconstruction of the epidemic in India. This is shown by the graph showing the three curves *A*_*tot*_, *A, R* (fig. 56).

**Figure 56.**
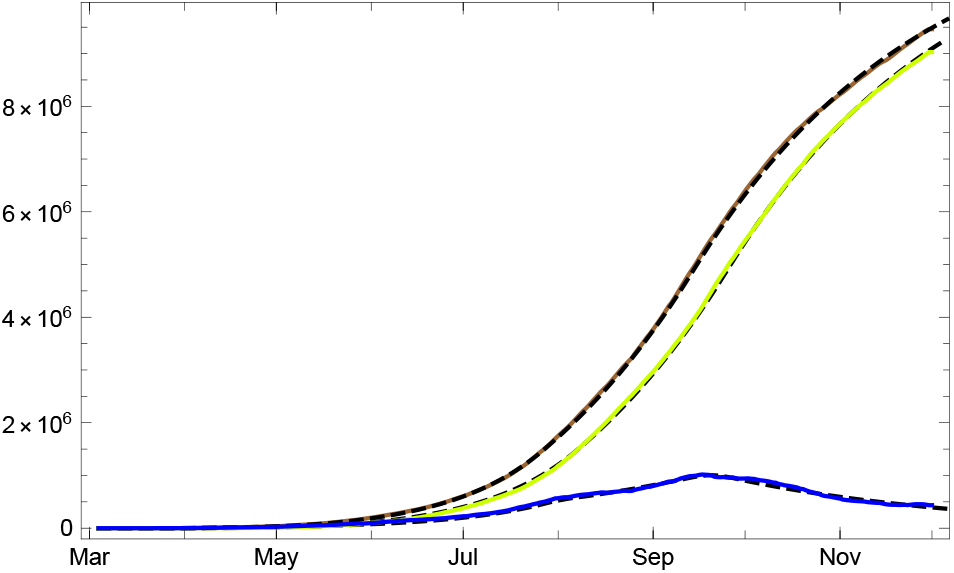
Empirical data (colored solid lines) for the numbers of totally infected *Â*_*tot*_(brown), redrawn 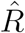 (bright green), actual infected *Â*(blue), and the respective model values (black dashed) for India. All with dark sector, *δ* = 35 and constancy intevals (see main text).

The numbers of newly reported *Â*_*new*_ (*k*) and the number of actual cases *Â*(*k*), including a conditional prediction for the next 30 days on the basis of the last infection strength *η*_6_ (fig. 57). Black dotted the boundaries of the 1 σ domain for the 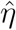 -variation in *J*_6_. In the case of India the data show exceptional low variability inside the constancy intervals. Thus the width of the 1 σ domain is smaller than in any of the other countries considered.

**Figure 57.**
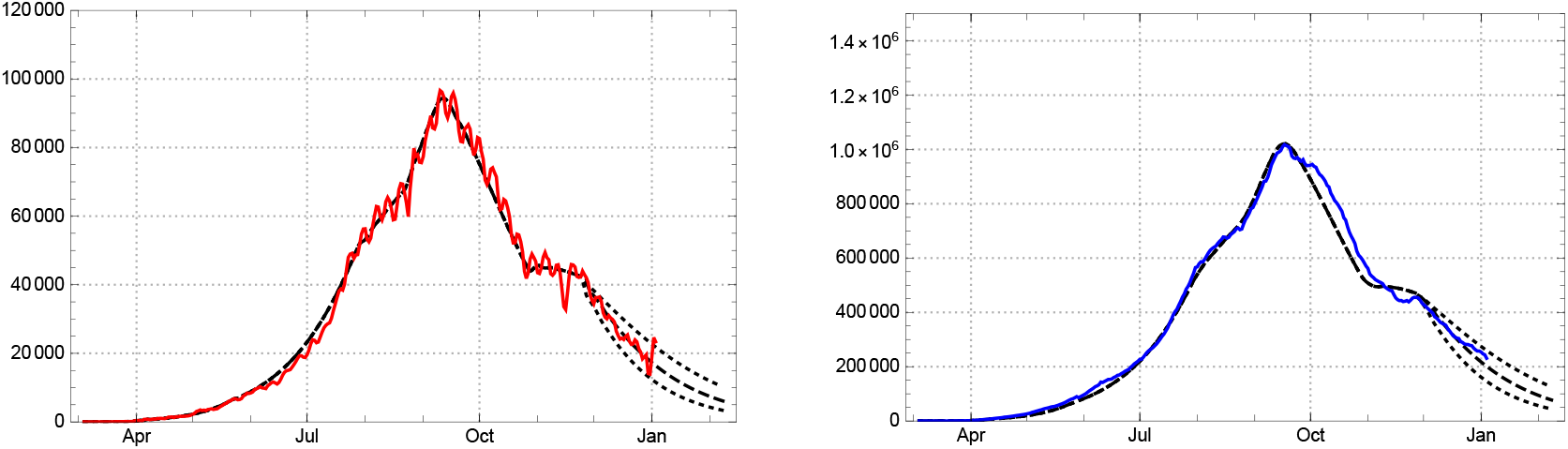
Left: Daily newly reported for India, empirical *Â*_*new*_ (solid red line) and model *A*_*new*_ (black dashed). Right: Reported actual cases for India, empirical *Â*(blue) versus model *A* (black dashed). Both with dark sector, *δ* = 35 and 30-day conditional prediction assuming no large vchange of the infection strenght in *J*_6_, the last main interval.

With a dark sector roughly as large as assumed in the model (*δ* = 35) the ratio of susceptibles went down in late 2020 to *s*(*k*) ≈0.7 (fig 58). This is the background for the reassuring prognosis for the development in India at the beginning of 2021 (fig. 57).

**Figure 58.**
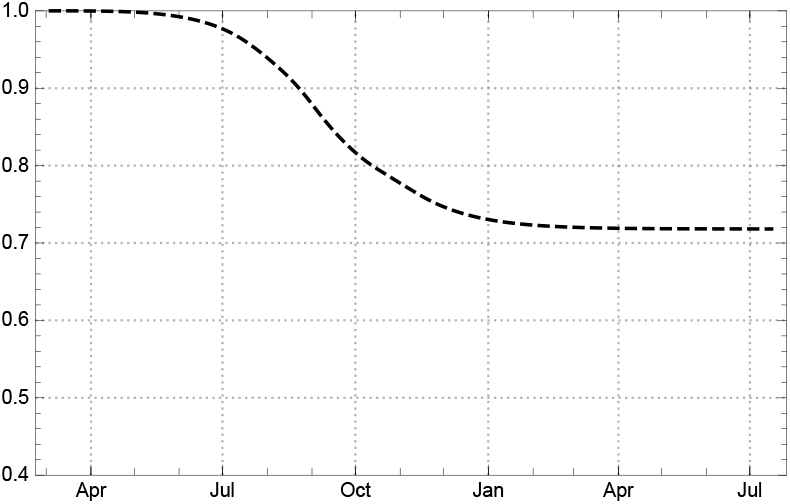
Ratio of susceptibles *s*(*k*) for India.

### 3.3. Aggregated data of the World

Let us now see how the aggregated data of all countries and territories documented in the JHU data resource can be analysed in our framework, and how they are reproduced by the SEPAR_*d*_ model. For the sake of simplicity we speak simply of *the World*. The number of daily new infections *Â*_*new*_ shows clearly three or four steps, expressed by phases of accelerated growth of *Â*_*new*_ between February and December 2020 (fig. 59): March (European countries), May to July (two waves in the US, bridged by rising numbers in Brazil and India), October (second wave in Europe and Brazil, third wave in US), and less visible the January/February wave in China and South-Korea.

**Figure 59.**
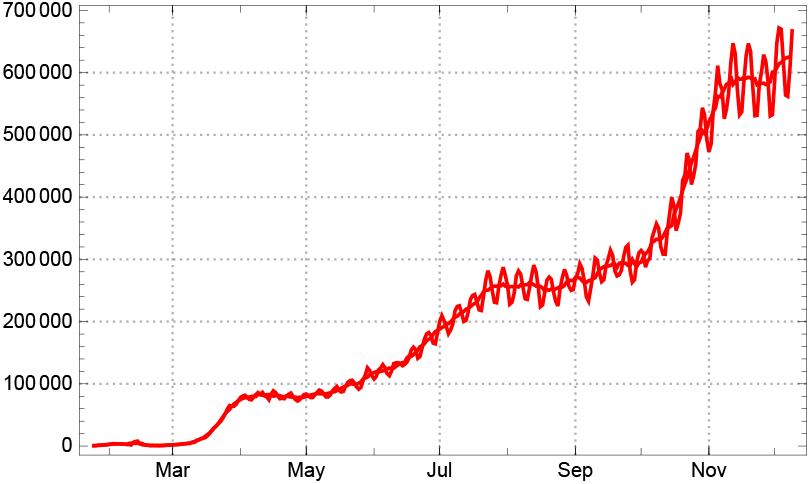
Daily new reported cases *Â*_*new*_ (*k*) for the World and 7-day sliding averages *Â*_*new*,7_(*k*).

These steps of steeper increase of the daily new infections correspond to local peaks or elevated levels of the mean strength of infection and reproduction numbers. The first two peaks of the mean reproduction numbers with *ρ*_*peak*−1_ ≈ 2 in January (China) and *ρ*_*peak*−2_ ≈2.5 in March (Europe) are followed by much lower phases of elevated levels from early May to early July, respectively in October 2020 (fig. 60).

**Figure 60.**
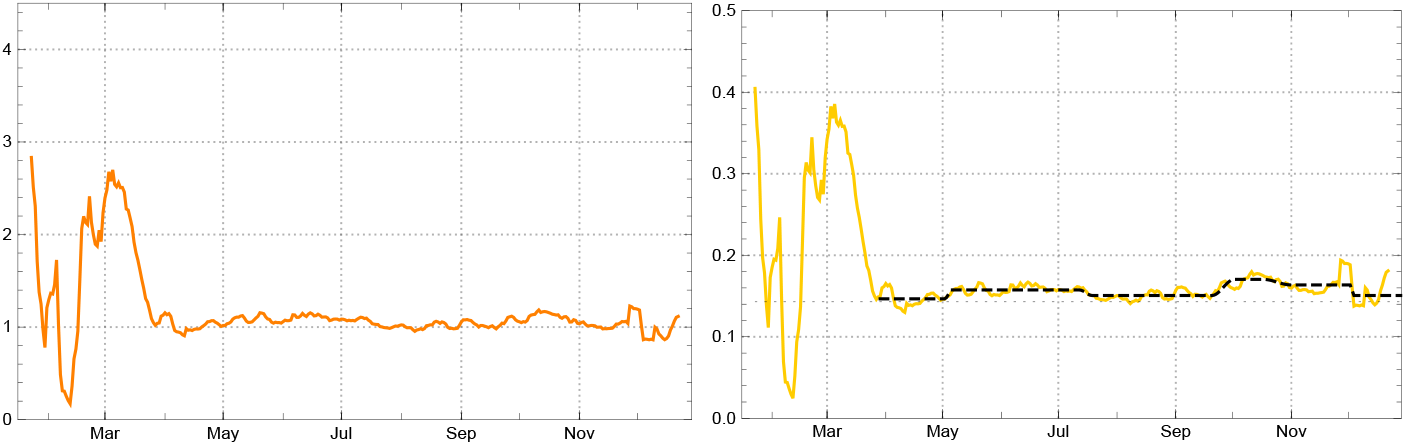
Bottom, left: Empirical reproduction rates 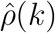 for the World (orange). Right: Daily strength of infection 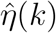 for the World (yellow) with model parameters *η*_*j*_ in the main intervals *J*_*j*_ (black dashed).

We let the model start at *t*_0_ = 01/25, 2020, the fourth day of the JHU day count, and use the following time separators for the main (constancy) intervals *t*_1_ = 03/29, *t*_2_ = 05/06, *t*_3_ = 07/19, *t*_4_ = 10/02, *t*_5_ = 11/10, *t*_6_ = 11*/*26, end of data *t*_*eod*_ = 12/11, 2020. In the count adapted to the *t*_0_ chosen here the main intervals are *J*_0_ = [1, 64], *J*_1_ = [65, 99], *J*_2_ = [103, 170], *J*_3_ = [177, 240]; *J*_4_ = [252, 276], *J*_5_ = [291, 303], *J*_6_ = [307, 322],]. The parameters *η*_*j*_ and the corresponding mean reproduction numbers in these intervals are give by the following table.

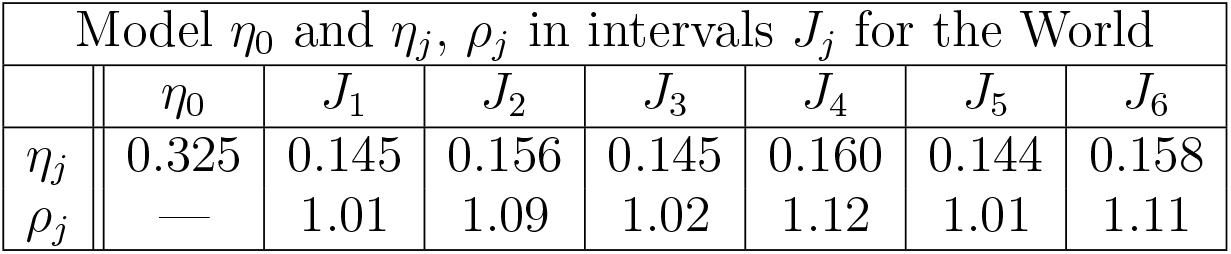

| Model $\eta_0$ and $\eta_j$, $\rho_j$ in intervals $J_j$ for the World | | | | | | | |
| --- | --- | --- | --- | --- | --- | --- | --- |
| | $\eta_0$ | $J_1$ | $J_2$ | $J_3$ | $J_4$ | $J_5$ | $J_6$ |
| $\eta_j$ | 0.325 | 0.145 | 0.156 | 0.145 | 0.160 | 0.144 | 0.158 |
| $\rho_j$ | — | 1.01 | 1.09 | 1.02 | 1.12 | 1.01 | 1.11 |

Because of the lack of reliable reporting for recovering dates in several countries, among them some large ones like the USA, we cannot expect a balanced value for the sojourn in the state of actual disease, documented in the statistics. Fig. 62, left shows that the estimated values 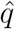 keeps close to 15 or even 20 until late March. Later on the weight of the countries with reliable documentation of recovering dates is large enough to keep the mean number of 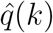 between 30 and 40, even with the rise of the pandemic after May 2020 (fig. 62, left). Accordingly the *q*-corrected number of actually infected *Â*_*q*_separate from the the ones given directly by the statistics *Â*(*k*) only in mid April. Since October 2020 they difference between the two is rising progressively (same figure, right).

**Figure 61.**
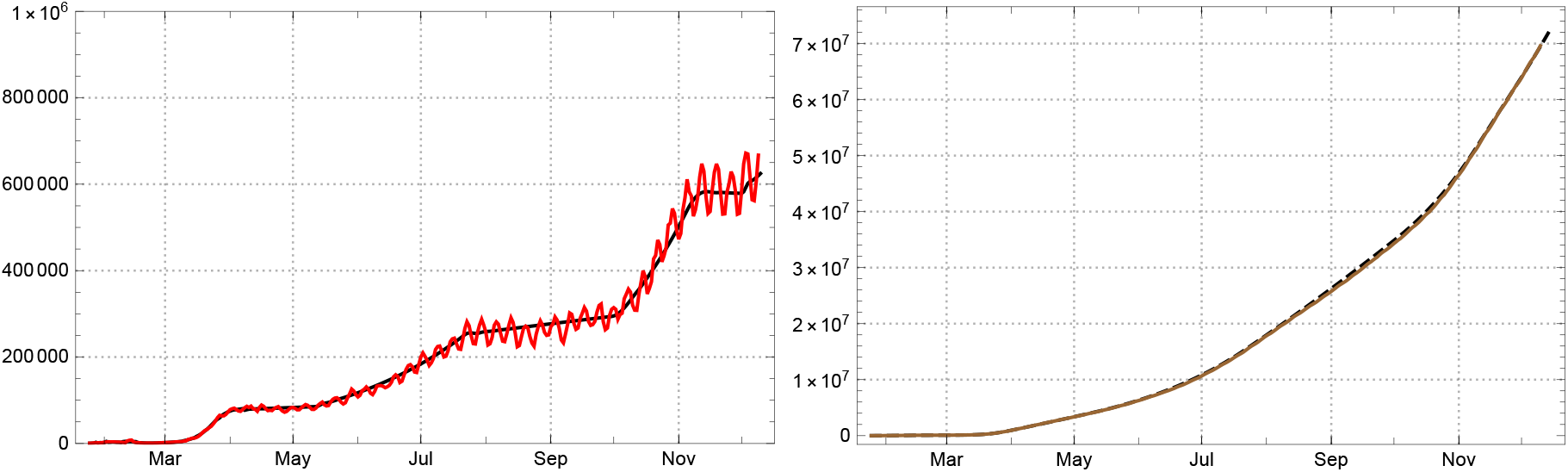
Left: 3-day averages of daily new reported infected for the the World (red); empirical *Â*_*new*_ solid red, model *A*_*new*_ black dashed. Right: Total number of reported infected (brown); empirical *Â*_*tot*_ solid, model *A*_*tot*_ dashed.

**Figure 62.**
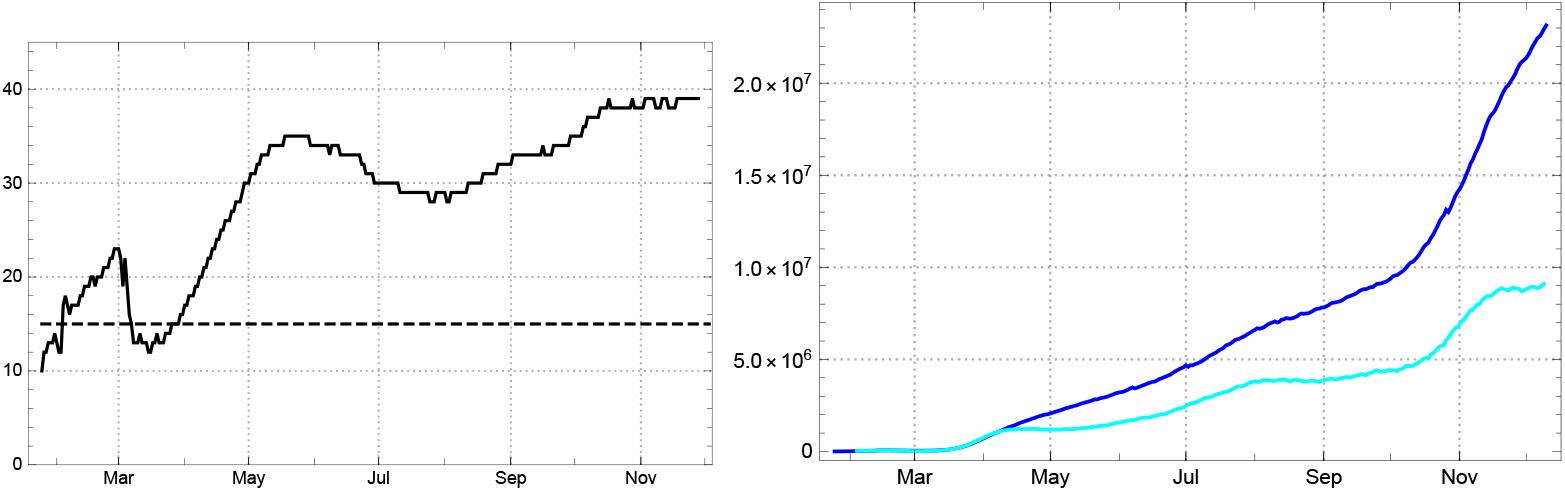
Left: Daily values of the mean time of statistically actual infection 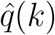 for the World. Right: Comparison of reported infected *Â*(dark blue) and *q*-corrected number (*q* = 15) of recorded actual infected *Â*_*q*_ (bright blue) from the JHU data in the World.

Like in the case of those countries which have an unreliable documentation of the actual state of infected (e.g. US, Sweden, …) we can here reconstruct the statistically given number *Â*(*k*) by the model value *A*(*k*) by using time variable durations 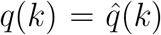. This is being displayed in the graph of the 3 curves of the World (fig. 63).

**Figure 63.**
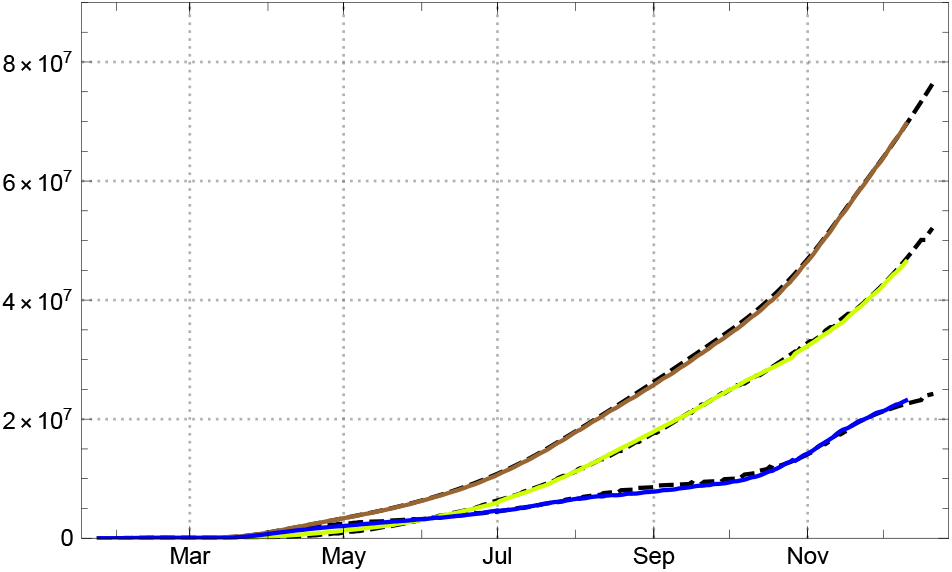
Empirical data (solid coloured lines) and model values (black dashed) for the World: numbers of totally infected *Â*_*tot*_ (brown), redrawn 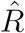 (bright green), and numbers of those which are statistically displayed as actually infected *Â*(blue).

## 4. Discussion

The data evaluation in sec. 3 shows clearly that the SEPAR_*d*_ model works well for countries or territories with widely differing conditions and courses of the epidemic. For the “tautological” application of the model with daily changing coefficients of infection *η*(*k*) this is self-evident, while it is not so for the use of a restricted number of constancy intervals. The examples studied in this paper show that in this mode of application the model is well-behaved, able to characterize the mean motion of an epidemic and to analyse its central dynamic. In the country studies we have shown that this is the case not only for the number of acknowledged daily new reported, our *A*_*new*_(*k*) but also for data which, in the standard SIR approach, are not easily interpretable like the number of actual infected persons, *A*(*k*)or the *q*-normalized number *A*_*q*_(*k*).

What is the *SEPAR*_*d*_ model good for? It is clear that it cannot predict the future. The main reason for this is that nobody knows how the contact rates are changing in the future. It allows – though – a prediction under assumptions. In the different countries we carried this out with different scenarios.

The main value of the model is as a tool for analysing the development, and to learn from such an analysis. We will discuss three such topics:

– the role of constancy intervals
– the role of the dark sector
– the influence of the time between infection and quarantine

### The role of constancy intervals

The empirical values of the infection strength 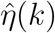 are calculated from data on reported new infected and are therefore subject to irregularities in data taking and reporting. The most drastic consequences of this are the obvious weekly fluctuations. Different methods can be applied to smooth these weekly fluctuation, sliding 7-day averages (used here), stochastic estimate used by the RKI (see appendix), band filter etc. Independent of the applied method there remain effects (e.g. non-weekly reporting delays) which distort the calculated numbers away from being correct empirical values for the intended quantities (e.g. *η*(*k*) = *γκ*(*k*)). Even if they were, one would encounter day to day fluctuations resulting from the variation of intensities of contacts and of the strengths of infectiousness involved, which one is not really interested in if one wants to gain insight into the dynamics of the epidemic. For this one needs to distil a cross-sectional picture of the process. In our approach this is achieved by constructing *constancy intervals* (main intervals) *J*_*j*_ and model strengths of infection *η*_*j*_, read off from the data, and to apply the infection recursion (5).

### The role of the dark sector

With increasing numbers of herd immunized, the influence of the dark sector on the ratio *s*(*k*) of susceptibles in the total population gains increasing weight, in particular for countries in which a high dark ratio *δ* may be expected. In most of the European countries studied here we find the ratio of recorded infected at the order of magnitude of 1 % all over the year 2020. With the non-reported ones added it can easily rise to the order of magnitude 10 % and start to have visible effects. If our estimated values of the dark factor *δ* are not utterly wrong, our model calculation shows that in nearly all countries of the study, Germany being the only exception, the development of the epidemic is already noticeably influenced by the dark sector at the beginning of the year 2021. The latter contributes essentially to turning the tide of the reported new infected, if one assumes constant contact ratios *κ*(*k*) and mean infection strength *γ* of the virus. Of course the appearance of new mutants may change *γ*, and counteract the decrease of the numbers of infected predicted by the model. This seems to be the main problem for the early months of 2021.

This becomes particularly succinct by a comparing the Swiss situation with Germany at the end of the year 2020 (fig. 64). In both countries containment measures were taken after a rise of the reproduction rate to 1.4 to 1.5 in late September /early October, although with different degrees of resoluteness and results (figs. 10, 19).

**Figure 64.**
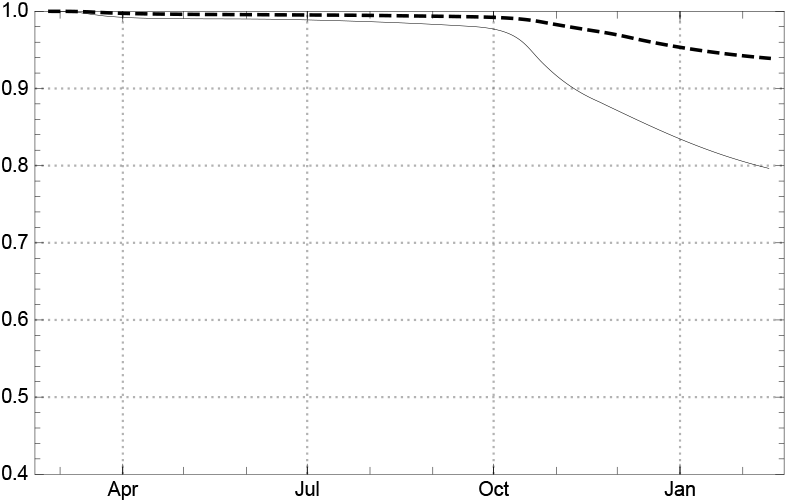
Ratio of susceptibles *s*(*k*) = *S*(*k*)*/N* for Germany (dashed) and Switzerland (solid line) at the end of the year 2020, assuming a dark ratio *δ* = 2 for Switzerland and *δ* = 1 for Germany.

The weight of the dark sector is much stronger for the non-European countries of our study. In the case of the USA and Brazil it has started to suppress the effective reproduction number below the critical value 1, according to our model assumptions on the dark factor. But even if one would set it dowm to *δ* = 1 or 2 the effect would already occur, although a bit later and weaker. That this is not yet reflected in the numbers of newly infected may have different reasons; one of it would, of course be, that the model can no longer be trusted in this region. Others have been mentioned in the country section. And finally it could be that persons infected some months ago need not necessarily be immune against a second attack. If virologists come to this conclusion, the whole model structure would need a revision. At the moment it is too early to envisage such a drastic step.

### The influence of the time between infectivity and quarantine

A central input into the *SEPAR*_*d*_ model is the assumption that there is a rather short period of length *p*_*c*_, where people, who later are positively tested, are infectious. This is closely related to the fact that people with positive test results are sent to quarantine or hospital. One can wonder what would happen, if the time between infectivity and quarantine or hospital is changed.

It is a bit confusing, but there are two answers to this question. To explain the difference we recall the role of *p*_*c*_ in our model. We usually derive the *η* parameters from the data (eq. 15). For the reproduction number (17) in the simplified *SEPAR*_*d*_-model with *p*_*c*_ = *p*_*d*_ = *p* and a constant coefficient *η* this means:

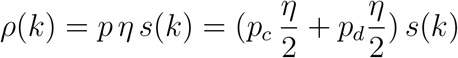

Here we assume that *p*_*c*_ is given. This number is only a rough estimate and may be chosen slightly differently. So, for each choice of the estimated number *p*_*c*_ one gets model curves and one might ask, how much these model curves differ, in particular how much the reproduction rates would differ. The answer is: not very much. The reason is that *p*_*c*_ enters implicitly also in the formula (14) for *η*, since the denominator is a sum over *p*_*c*_ values of the daily newly infected. If we assume that this number is constant (which often is approximately the case) then in the denominator we would have the factor *p*_*c*_ and in the formula for *ρ* it cancels out. Thus in this situation the reconstruction of *ρ* from *p*_*c*_ and *η* is independent of the choice of *p*_*c*_. If the values of the newly infected changes more drastically this is not the case and one has to use the general formula (16), but the difference is not dramatic. So the first answer to the question is: A different estimation for *p*_*c*_ does not have a noticeable influence for the model curves.

For understanding the second very different answer we have to recall that *η* may be interpreted as the product of the contact rate *κ* (as measured in the model) and the strength of the infection *γ*. If we assume that *γ* is constant, the change of *p*_*c*_ discussed above amounts to a change of the model-*κ*, which does not express a changing contact behaviour. This means that our measure for the contact rate is related to our choice of *p*_*c*_.

Now we come to the second answer. Here we assume that the contact rate remains the same, the contact behaviour of the society is not changed. But suppose that by some new regulations the value of *p*_*c*_ is changed. Then, as expected, if the contact behaviour is unchanged the reproduction number changes proportionally and so the curves are different. This second answer is what we are interested in here. Let us assume that one finds means by which the time until the people go to quarantine or hospital is reduced. Then less contacts take place and so the curves are flattened. This fact is well known, e.g., [2, appendix].^12^ But how much?

For answering this question we have taken the model description for Germany, lowering the value of *p*_*c*_ from 7 to 6 days from a certain moment on. Here we have to discuss an important point. One can only influence the time until quarantine or hospital for those who are registered, while the infected people who end up in the dark sector behave as before. At this moment we have to give up our assumption that *p*_*c*_ = *p*_*d*_. So, from a certain moment on we assume that *p*_*d*_ is still 7 but *p*_*c*_ is 6.

We have carried this out in two different scenarios for the expected numbers of daily new recorded infected *A*_*new*_(*k*) and the numbers of actual infected *A*(*k*). In the first one we compare the past development in Germany during the year 2020 with a fictitious reduction of *p*_*c*_ from 7 to 6 during May 2020, keeping *p*_*d*_ = 7 fixed (fig. 65). In a second one we take a look into the future, perpetuating the contact rate of the last constancy interval, i.e., assuming that the contact behaviour of the population is unchanged for a while and assume the same fictitious reduction as above in the second half of January 2021 (fig. 66). This doesn’t mean that we make a prediction of the future, our only aim here is to demonstrate what would happen if we could lower *p*_*c*_ from 7 to 6. The lowering of the numbers of infected, newly recorded and actual ones, are very impressive.

**Figure 65.**
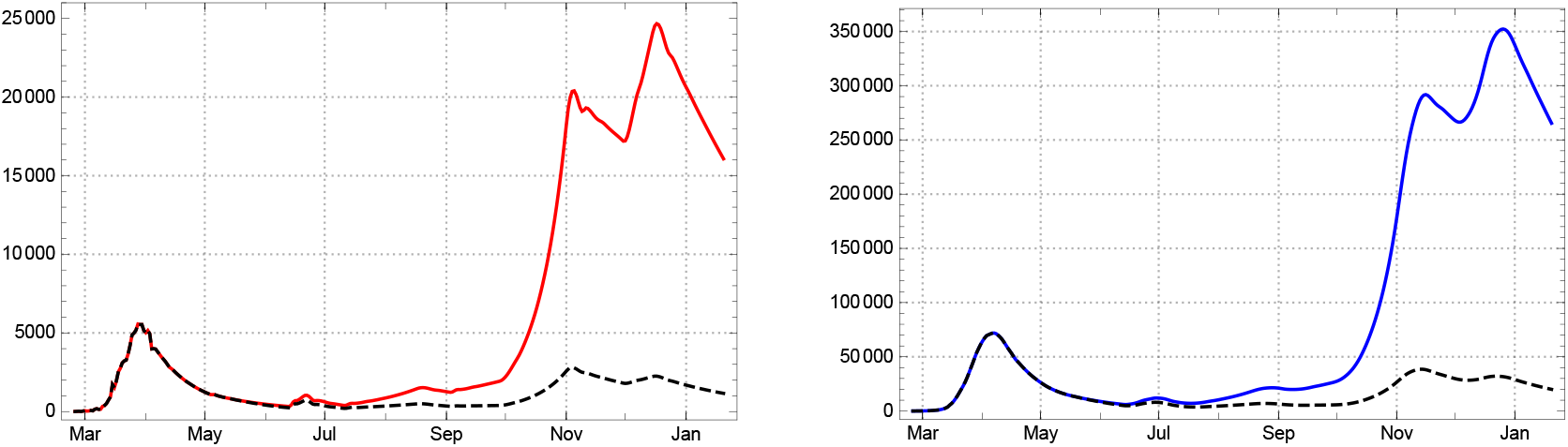
Model calculations for reported new infected *A*_*new*_(*k*) (left) and reported actual infected *A*(*k*) (right) for Germany. Solid lines with parameter values given in sec. 3(*p*_*c*_ = 7 all over the year 2020). Dashed *p*_*c*_ = 7 from March to May, *p*_*c*_ = 6 from August onward, smooth transition in June.

**Figure 66.**
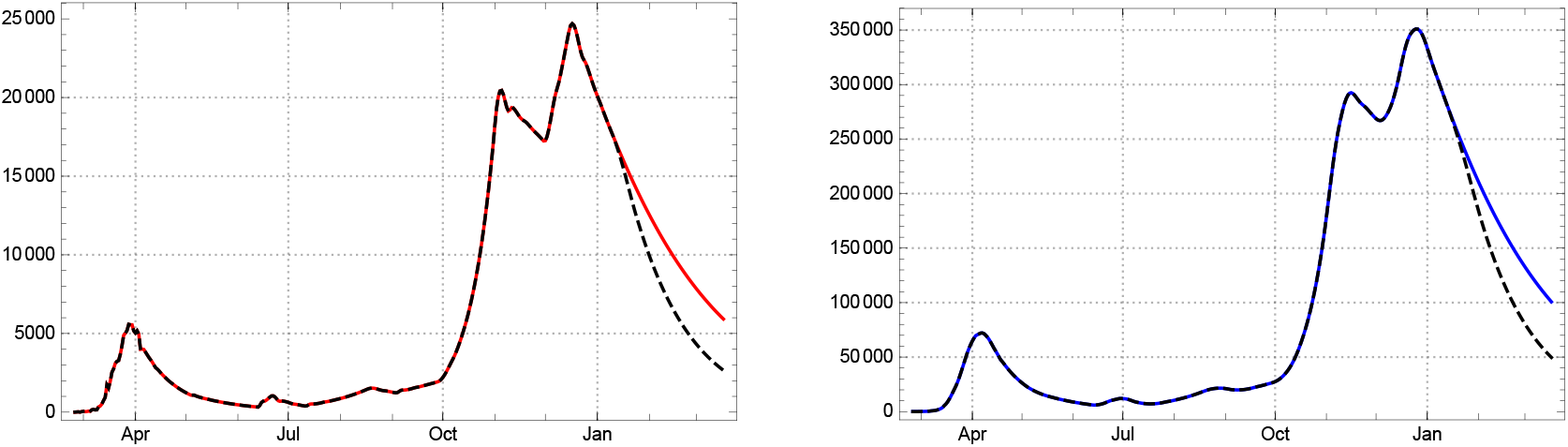
Model calculations for reported new infected *A*_*new*_(*k*) (left) and reported actual infected *A*(*k*) (right) for Germany (30 days prediction on the basis of data available 14 Jan 2021). Solid lines with parameter values given in the sec. 3, in particuilar *p*_*c*_ = 7. Dashed *p*_*c*_ = 7 from March 2020 to 15 Jan 2021, *p*_*c*_ = 6 from February 2021 onward, smooth transition in between.

In the past none of the regulations imposed by the German federal authorities made an attempt to reduce the time until people got to quarantine or hospital aside from raising the number of tests. Our considerations suggest to make a serious attempt in this direction. It has the big advantage that it does not require additional restrictions of the majority of the population and can be expected to be very effective at the same time.

## Appendix

### Comparison with RKI reproduction numbers

The estimates of the reproduction numbers for Germany by the *Robert Koch Institut* (RKI), Berlin, are based on an approach using the generation time as crucial delay time. The *generation time τ*_*g*_ of an epidemic is defined as the mean time interval between a primary infection and the secondary infections induced by the first one; similarly the length *τ*_*s*_ of the *serial interval* as the mean time between the onset of symptoms of a primary infected and the symptom onset of secondary cases. There are various methods to determine time dependent effective reproduction numbers on the basis of stochastic models for infections using both intervals. In our simplified approach with constant *e* and *p* these numbers correspond to 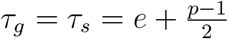.

The RKI calculation uses a method of its own for a stochastic estimation of the numbers of newly infected, called *E*(*t*), from the raw data of newly reported cases, described in [1]. The calculation of the reproduction numbers works with these *E*(*t*) and assumes constant generation time and serial intervals of equal lengths *τ*_*g*_ = *τ*_*s*_ = 4 [17]. ^13^ Two versions of reproduction numbers are being used, a day-sharp and therefore “sensitive” one 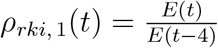, and a weekly averaged one,

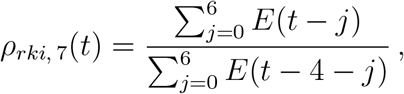

which we refer to in the following simply as *ρ*_*rki*_(*t*).

The paper remarks that the RKI reproduction numbers (“*R*-values”) *ρ*_*rki*_(*u*) indexed by the date *u* of calculation refer to a period of infection which, after taking the incubation period *ι* between 4 and 6 days into account, lies between *u* − 16, …, *u* – 8 (with central day *u* − 12 in the interval). We reformulate this redating by setting

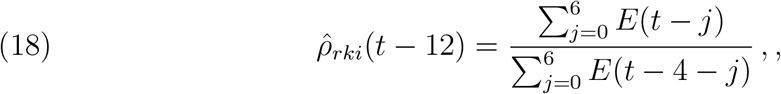

For a comparison with the SEPAR reproduction numbers we write (17) as

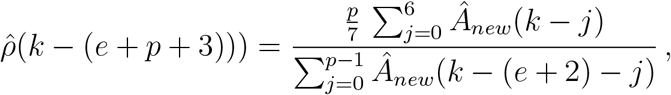

which for *e* = 2, *p* = 7 boils down to

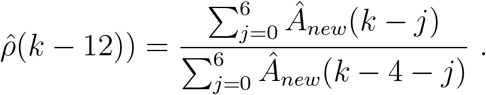

This is very close to (18). The main differences lie in the usage of different raw data bases (RKI versus JHU) and the adjustment of the raw data (stochastic redistribution *E*(*t*) versus sliding 7-day averages *Â*_*new*,7_). This may explain the differences in the level of low or high plateaus shown in fig. 67 (with 1 day additional time shift).

**Figure 67.**
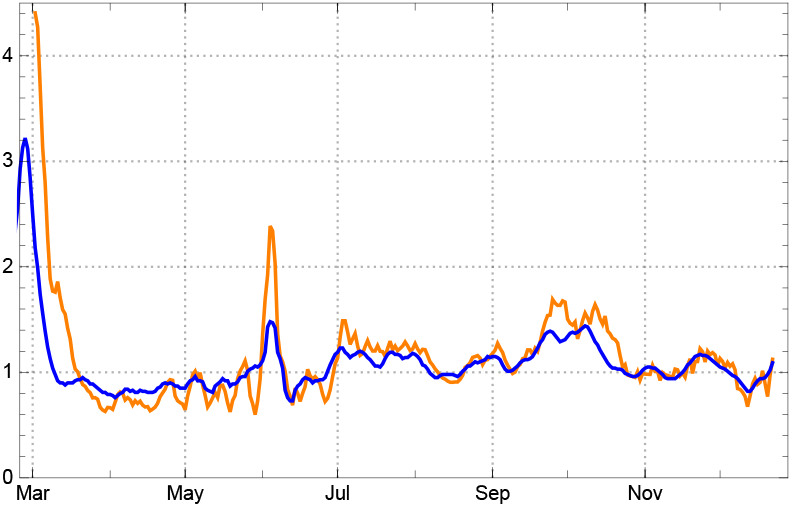
Empirical reproduction numbers 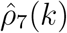 of the SEPAR_*q*_ model for Germany (orange) and reproduction numbers *ρ*_*rki*_(*k* − 13) (7-day averages) of the RKI (blue).

In this sense, our model supports the claim of the RKI that their reproduction numbers can be used as indicators of “a trend analysis of the epidemic curve” [17, p.1].

## Data Availability

The model works with data publicly available under https://data.humdata.org/dataset/novel-coronavirus-2019-ncov-cases

## Acknowledgements

We thank Odo Diekmann for discussing our thoughts as non-experts at an early stage of this work; he helped us to understand compartment models better. Moroever, we appreciate the exchange with Stephan Luckhaus, and thank Robert Schaback, Robert Feßler, Jan Mohring, and Matthias Ehrhardt for hints and discussions. Calculations and graphics were made with Mathematica 12.

## Footnotes

1 For Germany see [8, 18], for Switzerland [11] announcing a forth-coming study of *Corona-Immunitas*, for USA [15] and for India a report in ANI https://www.aninews.in/news/national/general-news/second-sero-survey-finds-2419-pc-of-punjab-population-infected-by-covid-1920201211181032/ retrieved 12/21 2020.

2 https://data.humdata.org/dataset/novel-coronavirus-2019-ncov-cases

3 E.g. in [19, p. 182] … A similar identification underlies the numbers for the active case in the *Worldometer* https://www.worldometers.info/coronavirus/country/.

4 This is done by the *Robert Koch Institut* for the German case.

5 Here we use the smoothing function described in [10]. Also an elementary optimization procedure for determining the main intervals is described in this paper.

6 Short report in [8]. This is consistent with the result in [18].

7 Such a hypothesis for the dark sector could be explained only by a drastic and irresponsible change of contact behaviour of the Swedish population or an increased infectivity of the virus. Neither of these explanations is supported by available empirical evidence.

8 The same holds for the UK.

9 A serological investigation of over 4000 inhabitants found 24 % infected (from which over 90 % were asymptomatic). With about 140 *k* reported infected in a population of roughly 31 *M* this amounts to a dark factor *δ* ≈50 (source ANI retrieved 12/21 2020 https://www.aninews.in/news/national/general-news/second-sero-survey-finds-2419-pc-of-punjab-population-infected-by-covid-1920201211181032/).

10 In 12/2020 it was already twice as much.

11 Only a permanently increasing amount of under-reporting could emulate a fake picture of a non-existing downswing of the epidemic for several months. We exclude such a hypothesis.

12 We thank S. Anderl for the hint.

13 For *e* = 2 this would correspond to *p* = 5, while for *τ*_*g*_ = *τ*_*s*_ = 5 we arrive at our *p* = 7.

